# First Phase 1b, single-center, age de-escalation trial of the *P. falciparum* blood-stage malaria vaccine candidate RH5.1/Matrix-M^™^: a delayed boost regimen induces high levels of functional antibodies in 5-17 month old Tanzanian infants

**DOI:** 10.1101/2024.03.25.24304862

**Authors:** Sarah E. Silk, Wilmina F. Kalinga, Jo Salkeld, Ivanny M. Mtaka, Saumu Ahmed, Florence Milando, Ababacar Diouf, Caroline K. Bundi, Neema Balige, Omar Hassan, Catherine G. Mkindi, Stella Rwezaula, Thabit Athumani, Sarah Mswata, Nasoro S. Lilolime, Beatus Simon, Hania Msami, Mohamed Mohamed, Damiano M. David, Latipha Mohammed, Gloria Nyaulingo, Bakari Mwalimu, Omary Juma, Tunu G. Mwamlima, Ibrahim A. Sasamalo, Rose P. Mkumbange, Janeth J. Kamage, Jordan R. Barrett, Lloyd D. W. King, Mimi M. Hou, David Pulido, Cecilia Carnrot, Alison M. Lawrie, Rachel E. Cowan, Fay L. Nugent, Rachel Roberts, Jee-Sun Cho, Carole A. Long, Carolyn M. Nielsen, Kazutoyo Miura, Simon J. Draper, Ally I. Olotu, Angela M. Minassian

## Abstract

**Background:** RH5.1 is a soluble protein vaccine candidate for blood-stage *Plasmodium falciparum* malaria, previously trialed in healthy UK adults in combination with AS01_B_ adjuvant. Here, RH5.1 was formulated with Matrix-M™ adjuvant to assess safety and immunogenicity in a malaria-endemic adult and pediatric population for the first time.

**Methods:** We conducted a Phase 1b, single-center, dose-escalation, age de-escalation, first-in-human trial of RH5.1/Matrix-M™ in Bagamoyo, Tanzania. Healthy adults (18-45 years) and infants (5-17 months) were recruited to receive the RH5.1/Matrix-M™ vaccine candidate in a variety of dosing regimens, including monthly dosing (0-1-2 month) or delayed booster dosing (0-1-6 month) using a 10 µg dose of RH5.1, or delayed fractional booster dosing (0-1-6 month) with the first two doses of RH5.1 at 50 µg and the third dose at 10 µg. All RH5.1 protein doses were formulated with 50 µg Matrix-M™ adjuvant. Primary outcomes for vaccine safety included solicited and unsolicited adverse events after each vaccination, along with any serious adverse events (SAEs) during the study period. Secondary outcome measures for immunogenicity included the concentration and avidity of anti-RH5.1 serum IgG antibodies by ELISA and their percentage growth inhibition activity (GIA) in vitro against P. falciparum parasites using purified IgG. All participants receiving at least one dose of vaccine were included in the primary analyses.

**Findings:** Between 25^th^ January 2021 and 15^th^ April 2021 a total of 60 adults and infants were enrolled; 57 of these completed the vaccination series, and 55 completed 22 months of follow-up post-third vaccination. Vaccinations were well-tolerated across both age groups. There were five SAEs involving four infant participants during the trial, none of which were deemed related to vaccination. RH5-specific T cell and serum antibody responses were induced by vaccination. The anti-RH5 serum IgG responses were significantly higher in the 5-17 month old infant groups as compared to adults. Serum antibody responses contracted over time post-third vaccination, but a similar hierarchy of responses across the age groups was maintained after 22 months follow-up (674 days post-third vaccination). Vaccine-induced anti-RH5 antibodies showed *in vitro* GIA with comparable functional quality across all age groups and dosing regimens. The highest anti-RH5 serum IgG responses were observed post-third vaccination in the 5-17 month old infants vaccinated with the 0-1-6 month delayed booster regimen using the 10 µg dose of RH5.1 (median 723 µg/mL; range: 450-1436 µg/mL), resulting in 100 % (11/11 infants) showing >60 % GIA following dilution of total IgG to 2.5 mg/mL (median 88 %; range: 73-97 %).

**Interpretation:** The RH5.1/Matrix-M™ vaccine candidate shows an acceptable safety and reactogenicity profile and highly promising antibody immunogenicity in 5-17 month old infants residing in a malaria-endemic area. The 0-1-6 month delayed booster regimen in 5-17 month old infants induced the highest levels of functional GIA reported to-date following human vaccination, with all infants achieving a level of GIA previously associated with protective outcome against blood-stage *P. falciparum* challenge in non-human primates. These data support onward efficacy assessment of this vaccine candidate against clinical malaria in young African infants.

**Funding:** The European and Developing Countries Clinical Trials Partnership (EDCTP).

**Trial Registration:** ClinicalTrials.gov: NCT04318002.

## Introduction

Malaria is a leading global cause of disease and death despite decades of interventions. In 2022 there were 249 million cases and 608,000 deaths, with over 75% of deaths occurring in children under the age of five in sub-Saharan Africa ^1^. Despite a welcome reduction in the global burden of disease between 2000 and 2015, this progress has since stalled and even reversed ^1^. Malaria prevention services were significantly disrupted by the COVID-19 pandemic, and drug and insecticide resistance, along with conflict and climate change, together pose significant ongoing threats; highly effective vaccines thus remain urgently needed.

Encouragingly, two malaria vaccines targeting the sporozoite stage of *Plasmodium falciparum*, RTS,S/AS01 (Mosquirix™) and R21/Matrix-M™, have now been recommended by the World Health Organization (WHO); both vaccines have shown efficacy against clinical malaria in young African infants in Phase 3 clinical trials ^2,3^. However, higher efficacy vaccines with improved durability remain needed, ideally without necessitating four doses. Moreover, efficacy against blood-stage infection with pre-erythrocytic vaccines requires creation and maintenance of sterile immunity, a high hurdle. Thus, addition of a complementary blood-stage vaccine strategy is attractive; such vaccines aim to prevent invasion of erythrocytes by merozoites, leading to control and clearance of blood-stage parasitemia, prevention of morbidity and reduced transmission ^4^. Additionally, a multi-stage malaria vaccine, consisting of partially effective anti-sporozoite and anti-merozoite components, provides a potential route to achieving a highly effective intervention; whereby the blood-stage component would eliminate any parasites that escaped the first (pre-erythrocytic) line of defense.

*P. falciparum* reticulocyte-binding protein homologue 5 (RH5) is the most advanced blood-stage vaccine candidate antigen. RH5 forms part of an invasion complex ^5,6^ and is both highly conserved and essential for invasion, where it mediates parasite binding to basigin on the erythrocyte surface ^7^. *In vivo* protection induced by RH5 vaccination was first demonstrated in an *Aotus* monkey model using a stringent heterologous *P. falciparum* challenge ^8^. Importantly, this study demonstrated that *in vivo* protection correlated with functional antibody activity, as measured by the *in vitro* growth inhibition activity (GIA) assay, and it enabled protective threshold levels of GIA and anti-RH5 serum IgG antibody to be defined. GIA was subsequently validated as a mechanistic immune correlate in *Aotus* by passive transfer of anti-RH5 monoclonal antibody ^9^.

The first RH5-based vaccine candidate to enter clinical trials was a viral-vectored formulation, consisting of the recombinant replication-deficient chimpanzee adenovirus serotype 63 (ChAd63) and the attenuated orthopoxvirus modified vaccinia virus Ankara (MVA). These were delivered to healthy UK adults in an 8 week prime-boost regimen (VAC057; ClinicalTrials.gov: NCT02181088). The viruses were safe and well-tolerated and elicited a functional antibody response with cross-strain *in vitro* GIA ^10^, although the level of GIA was below the *Aotus*-defined threshold of protection. However, given that another malaria vaccine candidate based on the same ChAd63 prime-MVA boost platform ^11^ had previously been shown to elicit 10-fold higher antibody levels in African infants (the target population for *P. falciparum* malaria vaccines) as compared with UK adults, testing of the ChAd63-MVA RH5 candidate in a malaria-endemic setting was warranted. A Phase 1b age de-escalation, dose escalation trial was therefore subsequently undertaken in Tanzania (VAC070; NCT03435874). This showed that the same viral-vectored RH5 vaccine candidate induced 10-fold higher anti-RH5 IgG responses in Tanzanian 6-11 month old infants as compared with UK adults, thus achieving the highest levels of functional GIA reported following human malaria vaccination at the time ^12^. In an attempt to optimize antibody quantity further, a full-length RH5 soluble protein vaccine, called RH5.1, was subsequently developed ^13,14^ and initially trialled in UK adults in a Phase 1/2a trial with GlaxoSmithKline’s (GSK’s) AS01_B_ adjuvant (VAC063; NCT02927145). This induced 10-fold higher antibody levels as compared to the viral-vectored RH5 in UK adults ^15^. Anti-RH5 serum IgG responses in UK adults were also more durable when the third dose was delivered at 6 months and at one-fifth of the initial dose (a so-called “delayed fractionation” (DFx) regimen), as compared to responses seen after three full (identical) doses given one month apart ^15^.

Here we report a Phase 1b clinical trial in Tanzanian adults and infants [VAC080; NCT04318002] that was therefore designed to assess the safety and immunogenicity of the RH5.1 soluble protein vaccine candidate, now formulated with Matrix-M^™^ adjuvant from Novavax. Following first-in-human safety assessment of this new formulation in adults, three different dosing regimens were assessed in the target infant population, 5-17 months of age (to align with delivery of R21/Matrix-M™ and RTS,S/AS01). This trial was designed to assess whether the levels of anti-RH5 serum IgG and functional GIA associated with a protective outcome in the *Aotus* monkey – *P. falciparum* challenge model could also be achieved in vaccinated African infants, and thereby inform decision-making on the progression of this RH5-based vaccine candidate to Phase 2b field efficacy testing.

## Results

### Study recruitment and vaccinations

The VAC080 Phase 1b trial was conducted in the Bagamoyo district of Tanzania. Malaria incidence in the district was estimated to range from 539 cases per 1000-person years in higher transmission areas to 76 cases per 1000-person years in lower transmission areas in 2020 ^16^. The protocol recruitment plan was to recruit two groups of healthy adults (aged 18-45 years) and three groups of 5-17 month old infants from Bagamoyo town, an area of relatively low malaria transmission within the district. The study was also designed to include a fourth infant group with relatively higher malaria pre-exposure, to assess for potential impact of prior malaria exposure on the immunogenicity of the RH5.1 /Matrix-M™ vaccine candidate. 185 participants were screened and 60 of these (N=12 adults and N=48 infants) were enrolled into the VAC080 Phase 1b trial (**Figure 1**). Prior to study start, an anti-parasite serological assay was established to provide a relative assessment of previous malaria exposure in the 5-17 month old infant age group. Serum IgG responses to parasite lysate of infants living in both urban (Dunda and Magomeni, N=90) and rural (Yombo and Fukayosi, N=15) wards within Bagamoyo district were used to define relative “low” versus “high” malaria pre-exposure thresholds relevant to this study population (**Figure S1A**). Within the infant cohort, 156 were tested for anti-parasite lysate responses during screening (**Figure S1B**), with N=36 recruited from those defined as relatively “low” prior malaria exposure and N=12 recruited from those defined as relatively “high” prior malaria exposure (**Figure S1C**).

**Figure 1:**
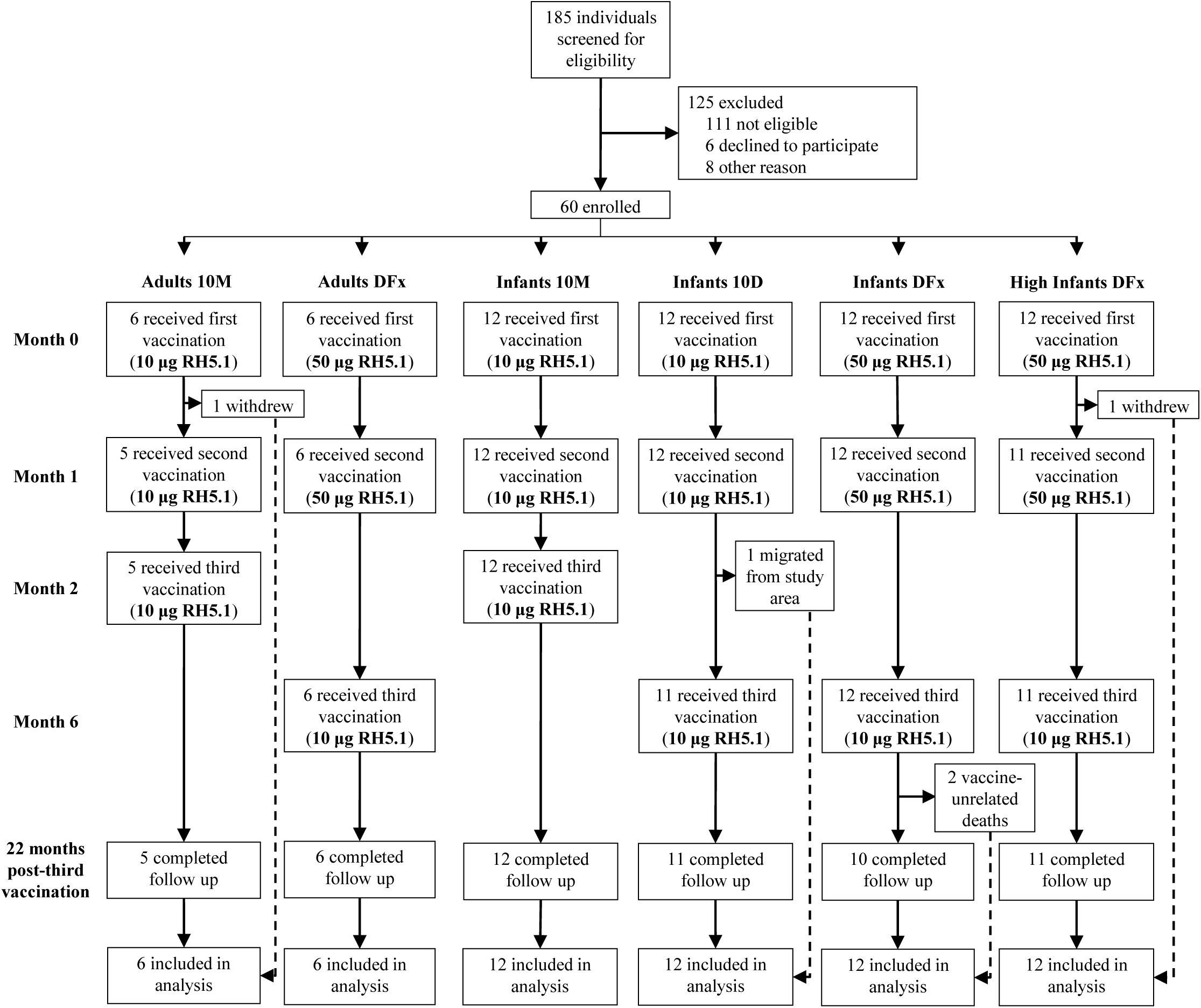
Trial profile. “Adults 10M” (adults aged 18-45 years) received 10 µg RH5.1 with 50 µg Matrix-M™ at 0, 1 and 2 months. “Adults Delayed-Fractional [DFx]” (adults aged 18-45 years) received 50 µg RH5.1 with 50 µg Matrix-M™ at 0 and 1 months and 10 µg RH5.1 with 50 µg Matrix-M™ at 6 months. “Infants 10M” (infants aged 5-17 months) received 10 µg RH5.1 with 50 µg Matrix-M™ at 0, 1 and 2 months. “Infants 10D” (infants aged 5-17 months) received 10 µg RH5.1 with 50 µg Matrix-M™ at 0, 1 and 6 months. “Infants DFx” (infants aged 5-17 months) received 50 µg RH5.1 with 50 µg Matrix-M™ at 0 and 1 months and 10 µg RH5.1 with 50 µg Matrix-M™ at 6 months. “High Infants DFx” (infants aged 5-17 months with higher prior malaria exposure based on anti-parasite lysate IgG at screening) received 50 µg RH5.1 with 50 µg Matrix-M™ at 0 and 1 months and 10 µg RH5.1 with 50 µg Matrix-M™ at 6 months. The main reason for participant withdrawal was consent withdrawal which was not due to adverse experience. Two participants died during the course of the study unrelated to study vaccination. All participants who received at least a single dose of RH5.1/Matrix-M™ were analyzed as part of the safety cohort. All participants who received at least a single dose of RH5.1/Matrix-M™ for whom data concerning immunogenicity endpoint measures were available were analyzed as part of the immunogenicity cohort. Two participants in the “High Infants DFx” group were recruited in error at below the “high” exposure threshold and are included in the immunogenicity analysis as indicated in subsequent figures.

Vaccinations began on 25th January 2021 and all 22 month post-third vaccination follow-up visits were completed by 27th July 2023. Group allocation was open label and non-randomized and enrolment proceeded in a dose escalation and age de-escalation fashion. Independent safety reviews were conducted by the Data Safety and Monitoring Board (DSMB) between every age de-escalation and dose-escalation step in the protocol. All vaccinations were administered via the intramuscular route in the deltoid and, in every case, the indicated dose of RH5.1 protein was formulated prior to injection with 50 µg Matrix-M™ adjuvant. Recruitment began with six adults in group “Adults 10M” who received 10 µg RH5.1 at 0, 1 and 2 months (monthly regimen), followed by six adults in group “Adults DFx” who received 50 µg RH5.1 at 0 and 1 months and 10 µg RH5.1 at 6 months (delayed-fractional regimen; DFx). Following a safety review, the four groups of twelve 5-17 month old infants were enrolled. Those in group “Infants 10M” (monthly regimen) and group “Infants 10D” (10 µg RH5.1 at 0, 1 and 6 months; delayed regimen) were recruited in parallel first, followed by group “Infants DFx”, who received the DFx regimen. Finally the group “High Infants DFx” was recruited as the relatively higher malaria pre-exposure cohort, and they also received the DFx regimen. Two participants in the “High Infants DFx” cohort were recruited with anti-parasite lysate IgG slightly below the serological threshold (**Figure S1C**); following a review they continued to receive the remaining vaccinations and study visits as scheduled.

The median age of adult participants was 23.5 years (range 20-30 years) and the median age of infant participants was 12 months (range 5-17 months), with comparable ranges across the groups (**Figure S2**). Similar numbers of males and females were enrolled across the infant age groups, however, although there were no sex-based exclusion criteria, no adult female participants were screened or recruited. There were three withdrawals during the vaccination period. Two participants withdrew after the first study vaccination, one in “Adults 10M” and one in “High Infants DFx”, and one participant in “Infants 10D” withdrew after the second vaccination. One withdrawal was due to the participant moving out of the study area and none were related to adverse experiences with study procedures. All other participants received the study vaccinations as scheduled. Two infant participants died during the post-vaccination follow-up period for reasons unrelated to the study or the study vaccinations as detailed below. All other participants completed follow-up out to 22 months post-final vaccination.

## Outcomes and estimation

### Reactogenicity and safety

There were no safety concerns during the course of the trial. The majority of local solicited adverse events (AEs) in both adults and infants were graded as mild to moderate in severity (**Figure 2**). Severe local solicited AEs were reported in one adult and three infant participants. The most common local solicited AE across all groups was swelling, reported after 18 % of vaccine doses (32/175 doses); this mainly occurred after the second and third doses and was mild (11 %; 19/175 doses), moderate (7 %; 12/175) or severe (1 %; 1/175) in graded severity. A slightly higher proportion of participants receiving the higher 50 µg dose of RH5.1 (those in groups “Adults DFx”, “Infants DFx” and “High Infants DFx”) experienced local solicited AEs of moderate to severe grade (14 %; 12/88 doses) compared to the lower dose groups (10 %; 9/87). Severe swelling and induration occurred in one adult participant from the lower dose group (“Adults 10M”) after the final vaccine administration; this resolved spontaneously within 7 days. All other local AEs in adults resolved by 72 hours post-vaccine administration. In the two infant DFx regimen groups, three infants developed severe and one developed moderate induration after receiving their final delayed dose. The majority of local solicited AEs in infants resolved within 72 hours post-vaccination, and all resolved by day 8.

**Figure 2:**
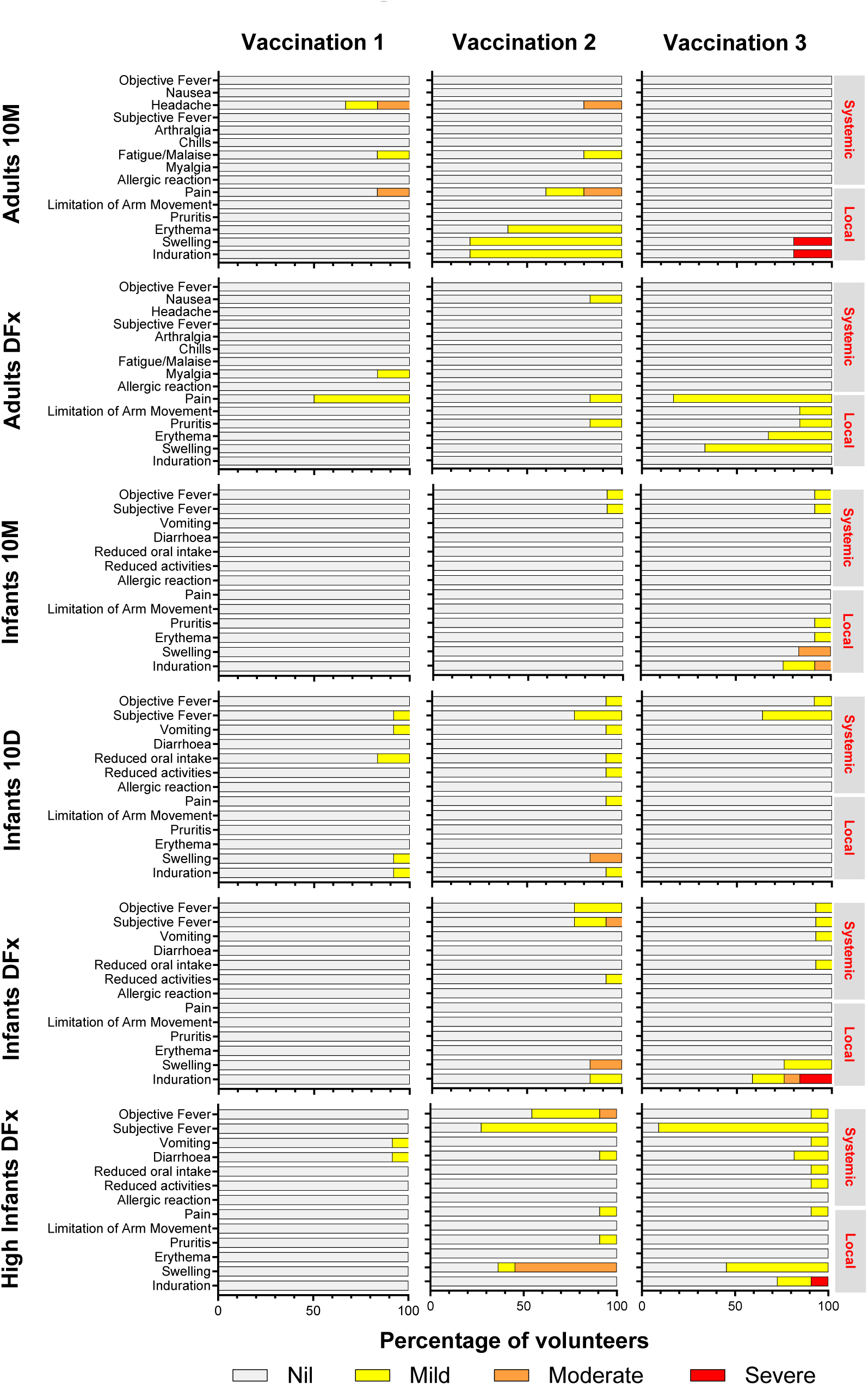
Solicited adverse events (AEs) following vaccination. Participants (or their carers) were asked about the presence of systemic and local AEs on the day of each study vaccination and for the following 7 days. The maximum severity reported by each participant for each AE after each study vaccination is shown. 10M = monthly vaccination regimen; DFx = delayed-fractional regimen; 10D = delayed regimen.

The majority of systemic solicited AEs were mild in severity, and there were no severe systemic solicited AEs reported (**Figure 2**). The most frequent systemic solicited AEs were mild subjective fever and mild objective fever, reported after 18 % (31/175) and 7 % (12/175) of vaccinations, respectively, occurring only in infants. One case of moderate severity subjective fever and one case of moderate severity objective fever were reported in the higher dose infant groups after receiving the second study vaccination. Vomiting, diarrhea, reduced oral intake and reduced activities were also experienced by infants only and were all of mild severity. All systemic AEs reported by adult participants were mild in severity, except for one participant who reported moderate grade headache on day one following both the first and second study vaccinations. In adults, most systemic solicited AEs resolved within 48 hours after vaccination and in infants most systemic solicited AEs resolved within 72 hours of vaccine administration. All systemic solicited AEs resolved by day 8.

Unsolicited AEs deemed at least possibly related to study interventions were all of mild or moderate severity (**Figure S3**). There was only one unsolicited AE in the adult groups – one participant reported an episode of mild dizziness six days following the third study vaccination which resolved spontaneously on the same day. There were five different unsolicited AEs in infants; all resolved within 8 days with the exception of one participant who was diagnosed with a helminth infection which was deemed probably related to vaccination due to the temporal association with study vaccination and lasted for 53 days. All unsolicited AEs had resolved by the end of the study period.

There were five serious adverse events (SAEs) involving four infant participants during the trial, none of which were deemed related to vaccination (**Figure S4**). Two of the five SAEs had a fatal outcome: one infant with stage IV neuroblastoma and one infant with acute respiratory failure secondary to herbal intoxication. The independent trial DSMB was informed of both cases and determined there was no relationship to study vaccinations. There were no AEs of special interest (AESIs) or suspected unexpected serious adverse reactions (SUSARs) during the course of the trial.

The most frequent laboratory abnormality identified in the 28-day post-vaccination period in adults was lymphopenia, occurring in 17 % (1/6) of “Adults 10M” participants and 33 % (2/6) of “Adults DFx” participants (**Figure S5**). All laboratory AEs occurring during this period in adults were mild in graded severity and resolved by the end of the study period, apart from one participant who had mild lymphopenia when they withdrew following their day 14 visit (post-first vaccination). The most frequent laboratory abnormality identified in the 28-day post-vaccination period in infants was anemia, occurring in 58 % of participants (7/12) in the “Infants 10M”, “Infants 10D” and “Infants DFx” groups, and in 42 % (5/12) in the “High Infants DFx” group. The maximum severity of anemia was moderate grade and in all cases had resolved by the final study visit (maximum duration 154 days). The majority of other laboratory AEs occurring during this period in infants were mild to moderate and all had resolved by the final study visit.

There were three severe grade laboratory AEs in infants in the 28 days following vaccination (**Figure S5**). One participant in the “High Infants DFx” group developed severe neutropenia at 7 days post-second vaccination which was temporally associated with a diagnosis of eczema with superimposed bacterial infection. This had improved to mild by day 14 and resolved completely by the third vaccination. One participant in the “Infants 10D” group developed severe neutropenia at 7 days post-second vaccination which resolved by day 14 and was temporally associated with a diagnosis of pneumonia. One participant in the “High Infants DFx” group developed severe eosinophilia at 7 days post-third vaccination which persisted at moderate grade until 28 days post-vaccination and had resolved by 84 days. There were no symptoms suggestive of allergy and stool microscopy did not show any evidence of helminth infection. This infant was well throughout this time period with no concerns apart from an isolated episode of fever on day 22.

All participants tested negative for malaria by microscopy at screening and were subsequently tested by microscopy at the second and third vaccination visits and monthly thereafter. Two participants in the “High Infants DFx” group (with higher prior malaria exposure) tested positive during study visits in the follow-up period (at 4 and 12 months post-third vaccination); these participants were clinically well and received anti-malarial treatment in the community. In addition, one participant in the “Infants DFx” group tested positive for malaria outside of the study during chemotherapy for neuroblastoma (see above section on SAEs).

### Immunogenicity

The magnitude of the serum anti-RH5 IgG antibody response was measured over time by ELISA against full-length RH5 (RH5.1) recombinant protein. Almost all participants showed background level responses at baseline (data not shown); consistent with numerous other immuno-epidemiological data that suggest RH5 is not a dominant target of naturally-acquired malaria immunity ^10,17,18^. Fourteen days following two vaccinations (denoted “V2+14”) of either 10 µg or 50 µg RH5.1 formulated with Matrix-M™ adjuvant, responses were comparable across the two doses within the adult or infant cohorts (**Figure 3A,B**). However, the magnitude of the antibody response showed a clear age-dependence, with significantly higher responses observed in the 5-17 month old infants (**Figure S6A**); here the adult cohorts showed median responses ranging from 38-52 µg/mL anti-RH5.1 IgG compared to the infant cohorts with ∼4-5 fold-higher median responses ranging from 200-230 µg/mL.

**Figure 3.**
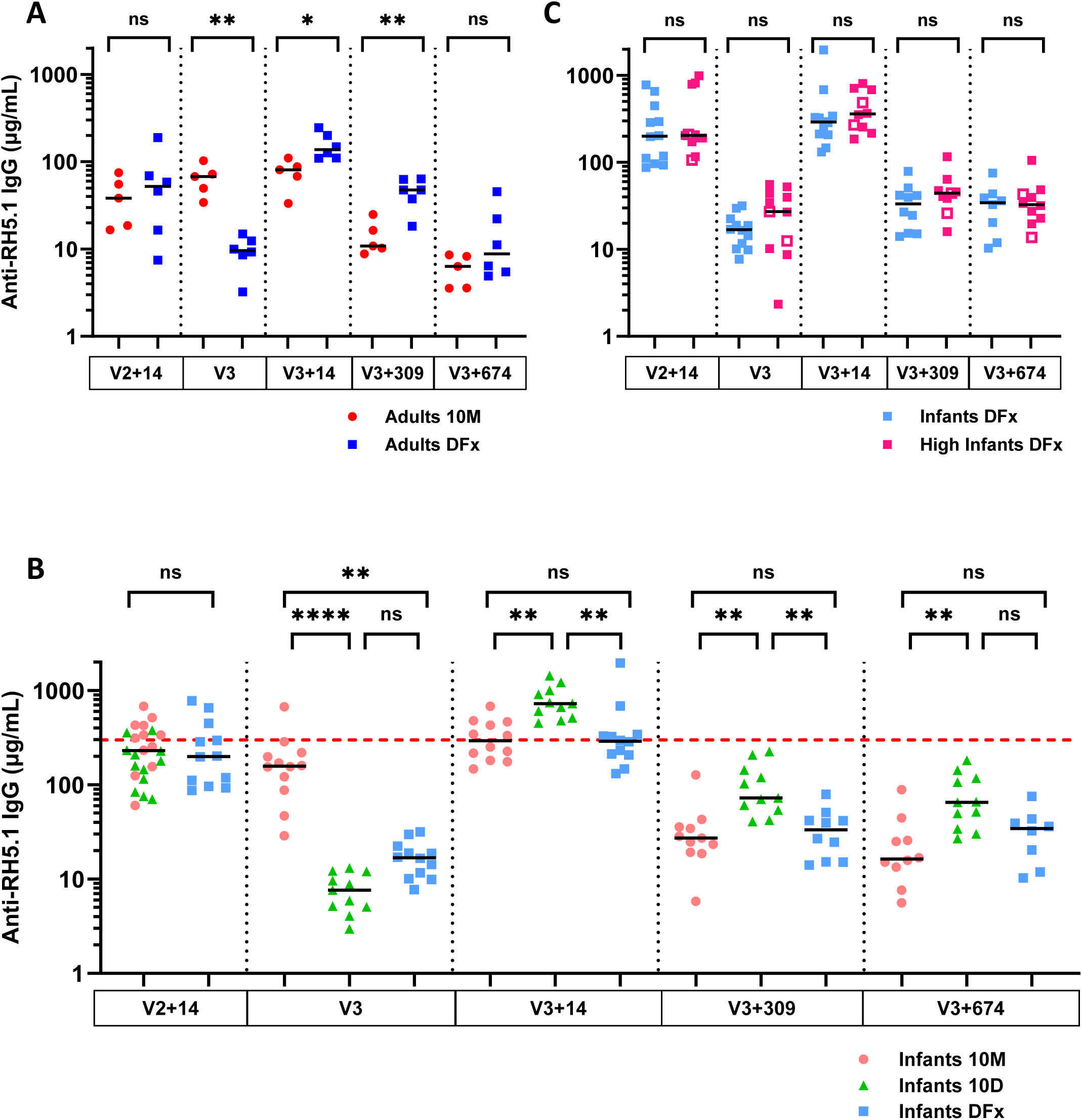
Serum antibody response to vaccination. Median and individual anti-RH5.1 serum total IgG responses as measured by ELISA are shown at 14 days post-second vaccination (V2+14), day of third vaccination (V3), and 14, 309 and 674 days post-third vaccination (V3+14 / 309 / 674) for (**A**) “Adults 10M” (N=5) and “Adults DFx” (N=6), and (**B**) “Infants 10M” (N=10-12), “Infants 10D” (N=11-12) and “Infants DFx” (N=8-12). “Infants 10M” and “Infants 10D” data were pooled at V2+14 due to identical immunization and dosing regimen at this time-point. The red dotted line indicates the protective threshold associated with protection against *P. falciparum* blood-stage challenge in RH5-vaccinated *Aotus* monkeys (∼300 µg/mL) ^8^. (**C**) ELISA data shown for the “Infants DFx” (N=8-12) and “High Infants DFx” (N=10-11) groups. Open symbols indicate N=2 participants who did not meet high pre-exposure threshold. Group kinetic responses are shown in **Figure S6C**. Analyses used Mann-Whitney test between dosing regimens within age groups only, \**P* < 0.05, \*\**P* < 0.01; and Kruskal-Wallis with Dunn’s multiple comparison test for V3, V3+14, V3+309 and V3+674 in (B) \**P* < 0.05, \*\**P* < 0.01, \*\*\**P* < 0.001, \*\*\*\**P* < 0.0001.

Responses then declined over time and were significantly lower prior to the third vaccination (V3) in all participants receiving the delayed or delayed-fractional boost at month 6, as compared to those receiving a final boost at month 2 (**Figure 3A,B**). Subsequently, 14 days following the third dose (V3+14; day 70 for monthly cohorts and day 196 for delayed and delayed-fractional cohorts), significantly higher responses (∼2-3-fold) were maintained in the 5-17 month old infants, as compared to adults, regardless of vaccination with either the monthly or DFx regimen (**Figure S6B**). Here, within the adult groups, responses were moderately but significantly higher at V3+14 in those receiving the DFx as compared to 10M regimen, with median responses of 138 µg/mL and 81 µg/mL, respectively (**Figure 3A**). In contrast, responses at V3+14 in the 5-17 month old infants receiving these same two regimens were comparable, with median responses at ∼290 µg/mL (**Figure 3B**). There was also no difference in the serum anti-RH5 IgG response at any time-point between the two infant cohorts who received the same DFx regimen, but who differed in baseline malaria pre-exposure status (**Figure 3C**). However, in contrast, the infants receiving the 10D regimen showed significantly higher responses, reaching a median of 723 µg/mL, with all infants in this group showing anti-RH5 serum IgG responses at the V3+14 time-point above the level associated with protection against *P. falciparum* blood-stage challenge in *Aotus* monkeys ^8^ (**Figure 3B**).

To further explore immune responses following the third vaccination, we also assessed the avidity of the anti-RH5 serum IgG response and the induction of RH5-specific IFN-γ T cell responses. Here, delayed and DFx boosting, as opposed to monthly boosting, were both associated with significantly more avid anti-RH5 IgG responses in adults and infants (**Figure S7**). In contrast, the anti-RH5 IFN-γ T cell response, as assessed by *ex-vivo* ELISpot assay ^15^, was comparable across all groups, regardless of age or vaccination regimen (**Figure S8**).

Anti-RH5 serum IgG antibody responses subsequently declined over time (**Figure S6C**), but a similar pattern of immunogenicity to the peak response was maintained at ∼1 year post-third vaccination (V3+309), although no significant differences were observed at ∼2 years (V3+674) between the monthly and DFx regimens within the adult and infant cohorts (**Figure 3A,B**). The highest responses were maintained at V3+674 in the Infants 10D cohort, with a median of 65 µg/mL; these were significantly higher than those in the Infants 10M cohort with median of 16 µg/mL (**Figure 3B**).

Serum samples were next analyzed for *in vitro* functional anti-parasitic activity at the GIA Reference Center (NIAID/NIH). Purified total IgG was normalized to a starting concentration of 10 mg/mL and tested against *P. falciparum* (3D7 clone) parasites using the standardized single-cycle GIA assay. Samples at baseline (pre-vaccination) were negligible (<20 % GIA), with a single exception of one adult who had 58 % GIA (**Figure 4A**), however large increases in overall GIA were observed post-RH5.1/Matrix-M™ vaccination (**Figure 4B**). Here, at the V3+14 time-point (14 days post-dose 3) and using 10 mg/mL total IgG, median levels ranged from 83-87 % and 95-98 % GIA across the adult and infant groups, respectively. All total IgG samples with >40 % GIA at 10 mg/mL were subsequently titrated using a 2-fold dilution series in the assay (**Figure S9A**). The overall levels of GIA mirrored the previous serology, with infants overall showing higher levels of GIA as compared to adults and as measured using 2.5 mg/mL total purified IgG. Moreover, the infants vaccinated with the 10D regimen showed the highest levels of GIA, significantly outperforming the 10M and DFx regimens tested in the other infant groups (**Figure 4C**). There were also no differences seen in the levels of GIA between the two infant cohorts who differed in baseline malaria pre-exposure status and received the same DFx regimen (**Figure 4D**).

**Figure 4.**
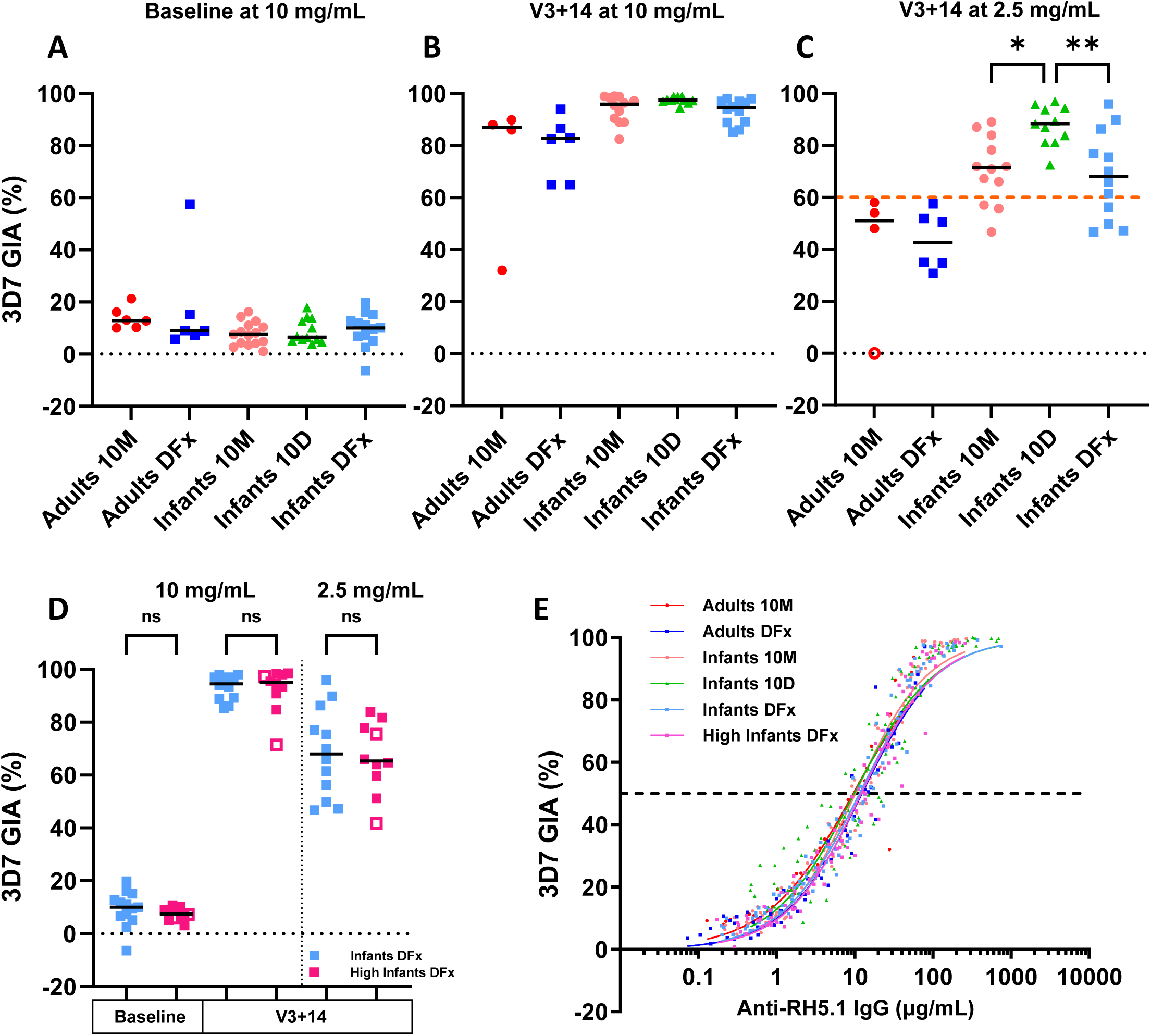
Functional GIA induced by RH5 vaccination. *In vitro* GIA of total IgG purified from serum was assessed against 3D7 clone *P. falciparum* parasites. Median and individual values are shown for each group at (**A**) Baseline (day 0) at 10 mg/mL total IgG; (**B**) 14 days post-third vaccination (V3+14) at 10 mg/mL total IgG; and (**C**) V3+14 at 2.5 mg/mL total IgG (“Adults 10M” N=4-6; “Adults DFx” N=6; “Infants 10M” N=12; “Infants 10D” N=11; “Infants DFx” N=12). Open circle datapoint in (C) is arbitrarily set at 0 % GIA and for the sample not titrated in the assay. Red dashed line in (C) indicates threshold of 60 % GIA associated with protective outcome in RH5-vaccinated and *P. falciparum* challenged *Aotus* monkeys ^8^. Analysis between infant dosing regimens by Kruskal-Wallis with Dunn’s multiple comparison test, \**P* < 0.05, \*\**P* < 0.01. (**D**) GIA of samples at baseline and V3+14 tested at 10 mg/mL and 2.5 mg/mL total IgG from the “Infants DFx” (N=12) and “High Infants DFx” (N=10-12) cohorts. Open symbols indicate N=2 participants who did not meet the high pre-exposure threshold. Analysis used Mann-Whitney test to compare between the two groups; ns = not significant. (**E**) V3+14 samples were titrated in the GIA assay using a 2-fold dilution series starting at 10 mg/mL. Relationship between GIA data from the dilution series and the concentration of anti-RH5.1-specific IgG used in the assay as measured by ELISA in the total IgG is shown. Non-linear regression curves (constrained to >0 % and <100 % GIA) are shown for each group. The EC_50_ (concentration of anti-RH5.1 polyclonal IgG that gives 50 % GIA) was calculated (dashed line represents 50% GIA) using data points pooled across all groups (r^2^ = 0.95, N=487).

The GIA showed a strong relationship to the concentration of RH5.1-specific IgG present in the total IgG used in the assay and as measured by ELISA (**Figure 4E**). The concentration of RH5.1-specific polyclonal IgG required to give 50 % GIA (EC_50_) was highly comparable across all the groups (regardless of age, vaccine regimen and malaria pre-exposure status at baseline). Analysis across all groups combined showed a GIA EC_50_ = 14.3 μg/mL (95% CI: 13.4-15.2 μg/mL). This functional “quality” readout of the RH5.1/Matrix-M™ vaccine candidate, i.e. the concentration of antigen-specific IgG required to achieve 50 % GIA, was consistent with our previously reported data following vaccination of healthy volunteers in the UK or Tanzania with viral-vectored RH5 or RH5.1/AS01_B_ ^10,12,15^ (**Figure S9B**).

We previously reported an association between the *in vitro* GIA assay and *in vivo* protection outcome following RH5 vaccination and *P. falciparum* blood-stage challenge in *Aotus* monkeys, with protected animals showing >60 % GIA at 2.5 mg/mL purified IgG ^8,9^. A similar association was observed in a controlled human malaria infection (CHMI) trial in healthy UK adults vaccinated with RH5.1/AS01_B_, although this threshold level of GIA was not reached in that study ^15^. Notably, in the VAC080 trial, all four groups of vaccinated infants showed median levels of GIA at 2.5 mg/mL total IgG over the level of 60 % GIA (**Figure S10A**). The Infants 10D group showed the highest median level of GIA at 88 % with all participants (11/11) above the 60 % GIA target threshold at day 14 following the third vaccination (**Figure 4C**). This level of GIA is the highest level of functional GIA achieved following human vaccination to date (**Figure S10B**).

## Discussion

Here we report a dose-escalation, age de-escalation, first-in-human clinical trial of the RH5.1/Matrix-M™ vaccine candidate, assessed in a malaria-endemic population. These data in healthy Tanzanian adults and 5-17 month old infants show that this adjuvanted protein subunit vaccine formulation has a favorable safety and reactogenicity profile, highly comparable to data from Phase 1b malaria vaccine trials using the R21 immunogen formulated with the same Matrix-M™ adjuvant ^19,20^. Independent review concluded none of the five SAEs (involving four infant participants) that occurred during the trial, including two with fatal outcomes, were related to vaccination (see **Supplementary Information**). These occurred against a similar backdrop child mortality rate (aged 1-5 years) of 26.5/1000 and infant mortality rate (aged 0-1 years) of 53.0/1000 in Bagamoyo District as last recorded in 2012 ^21^. Including this trial, safety data have now been reported for 193 healthy volunteers (109 adults, 18 children and 66 infants) vaccinated with RH5-based vaccines ^10,12,15^, all showing similar safety and tolerability profiles, whilst Matrix-M™ adjuvant is now a component in vaccines for malaria and Covid-19 that are licensed for human use ^3,22^. On-going Phase 1/2 trials, including a Phase 2b efficacy trial in >200 infants 5-17 months of age (VAC091; NCT05790889), continue to monitor the safety of RH5.1/Matrix-M™ vaccination.

The RH5.1/Matrix-M™ vaccine candidate was immunogenic for anti-RH5 IFN-γ T cell responses and serum IgG antibody across all age groups and dosing regimens. The levels of peripheral T cell response observed by ELISPOT assay were not dissimilar to those previously reported in UK adults immunized with the RH5.1/AS01_B_ formulation using the same dosing regimens ^15^. While the contribution of IFN-γ-secreting T cells to RH5-mediated malaria immunity in humans remains unclear, it is likely that RH5-specific CD4^+^ T cells are needed to help B cell responses. Indeed, previous work in UK adults has shown RH5.1/AS01_B_ can induce RH5-specific circulating T follicular helper (Tfh) cell responses and that these correlate with both RH5-specific memory B cells and serum antibody ^23^. Work remains on-going to investigate B and Tfh cell response phenotypes in the Tanzanian infant population vaccinated here with RH5.1/Matrix-M™.

Priming of Tanzanian adults with two doses of 10 or 50 µg RH5.1 formulated with 50 µg Matrix-M™ led to similar anti-RH5 serum IgG responses two weeks after the second dose. Notably, however, following the third dose, the 0-1-6 month DFx regimen induced significantly more avid and higher levels of anti-RH5 serum IgG, as compared to the 0-1-2 monthly regimen; consistent with our previous observations in UK adults vaccinated with the same dosing regimens of RH5.1 with AS01_B_ adjuvant ^15^. These serum IgG responses were better maintained over time in the DFx regimen vaccinees for at least ∼1 year following the third dose, although responses declined by ∼2 years. In line with this, we have also reported enhanced circulating RH5-specific memory B cell responses in adults vaccinated with the DFx (as opposed to monthly) dosing regimen, along with a higher frequency of RH5-specific (and putatively long-lived) plasma cells in the bone marrow that correlated strongly with the serum anti-RH5 IgG response ^24,25^.

Interestingly, these observations in adult vaccinees were only partly replicated in the 5-17 month old infants. Responses were again similar two weeks after the second vaccination, regardless of RH5.1 protein priming dose, however, following the third vaccination the monthly and DFx regimens performed comparably, whereas significantly higher anti-RH5 serum IgG responses were induced in those infants receiving a delayed, but not fractionated dose, of RH5.1 protein at 6 months. The underlying reasons for this improvement with the delayed, as opposed to DFx, regimen remain to be determined, although circulating levels of vaccine-specific antibody may affect vaccine responsiveness ^26^ and this cohort primed twice with 10 µg RH5.1 with Matrix-M™ showed lower anti-RH5 serum IgG responses at the time of third vaccination (month 6) as compared to those in the DFx regimen group who were primed twice with a higher 50 µg RH5.1 dose. In line with our observations here, trials of RTS,S/AS01 showed improved outcome measures with a DFx regimen, as opposed to 0-1-2 monthly dosing, in healthy US adults ^27^, however no improvement in field efficacy when tested in 5-17 month old infants ^28^. Notably, these DFx regimens tested with RTS,S/AS01 fractionated the dose of adjuvant and antigen, unlike our trials where a fractionated dose of antigen is delivered in a full dose of adjuvant. Nevertheless, our data suggest a delayed boosting, as opposed to DFx, regimen may be more optimal in this younger age group and this warrants further investigation.

Regardless of vaccination regimen, peak anti-RH5 serum IgG responses in the 5-17 month old infants were significantly higher than those induced in adults – a phenomenon now consistently observed with numerous other malaria subunit vaccine candidates including viral-vectored and recombinant antigen-in-adjuvant formulations such as R21/Matrix-M™ ^11,12,19,29,30^. These data bode well for future assessment of multi-stage malaria vaccine strategies, whereby a blood-stage RH5 component could be trialed in combination with the licensed pre-erythrocytic vaccines. The very high levels of anti-RH5 serum IgG induced in the 5-17 month old infants translated into the highest levels of functional GIA observed in human vaccinees to-date. Indeed, overall GIA largely mirrored the serological responses, and all infants vaccinated with the 0-1-6 month delayed boost regimen (who averaged ∼700 µg/mL RH5-specific IgG at the peak of the response) exceeded the threshold level of *in vitro* GIA that our work has previously associated with a protective outcome in *Aotus* monkeys challenged with blood-stage *P. falciparum* ^8,9^. Importantly, this GIA threshold was also reached in 75% of infants vaccinated with the 0-1-2 month regimen, suggesting RH5 vaccination could be aligned with the current delivery schedule for the primary series of RTS,S/AS01 and/or R21/Matrix-M™. The functional quality of serum antibody induced by RH5.1/Matrix-M™ (i.e. GIA per µg anti-RH5 IgG) was also highly similar across all age groups and vaccination regimens tested here, and also highly similar to that reported in all previous clinical trials of vaccine candidates based on the full-length RH5 molecule ^10,12,15^. Consistent with previous observations using RH5.1/AS01_B_ in UK adults ^15,25^, all the delayed booster regimens tested here in adults and infants also showed improvement in the avidity of anti-RH5 serum IgG, however, this did not impact the functional quality of these IgG in the assay of GIA. These observations have now been explained by analysis of anti-RH5 human mAbs that showed delayed boosting leads to improvement in the anti-RH5 antibody dissociation rates, but not in the antibody association rates which are strongly correlated with GIA potency ^31^.

Previous work on historical blood-stage vaccine candidates targeting the AMA1 antigen suggested that naturally-occurring anti-malarial antibody responses could interfere with vaccine-induced IgG ^32^, whilst other studies have suggested such antibodies may interact additively or synergistically with anti-RH5 IgG in terms of GIA ^33^. We therefore also assessed the DFx regimen in 5-17 month old infants who had experienced relatively higher levels of malaria exposure prior to study recruitment. Our data indicated no apparent impact of these anti-malarial responses on RH5.1/Matrix-M™ immunogenicity or functional GIA. Whether much higher levels of existing anti-malarial natural immunity at the time of RH5 vaccine priming may enhance or interfere with vaccine outcomes now remains to be determined in larger field trials at sites of higher malaria endemicity.

The main limitations of this trial were the relatively small numbers of participants recruited from an area of low to moderate malaria transmission, and that in the adult groups only males volunteered to participate. Nonetheless, the very high levels of GIA achieved in this trial indicate that a 0-1-6 month delayed boost regimen with RH5.1/Matrix-M™ may protect the 5-17 month old target population against blood-stage *P. falciparum* malaria. In light of this, the RH5.1/Matrix-M™ vaccine candidate has since progressed to a Phase 2b field efficacy trial in Burkina Faso (VAC091; NCT05790889) to assess efficacy against clinical malaria following monthly or delayed boost vaccination.

## Methods

### Study Design

The VAC080 trial was an open-label, age de-escalation, dose-escalation study, conducted according to the principles of the current revision of the Declaration of Helsinki 2013 and in full conformity with the ICH guidelines for Good Clinical Practice (GCP). It was approved by the Oxford Tropical Research Ethics Committee in the UK (reference 9-20), and the following authorities in Tanzania: the Ifakara Health Institute Institutional Review Board (reference 49-2020), the National Institute for Medical Research, the National Health Research Ethics Sub-Committee and the Tanzania Medicines and Medical Devices Authority (reference TMDA0020/CTR/0006/01). The VAC080 trial was registered on ClinicalTrials.gov (NCT04318002). The study was conducted at Ifakara Health Institute Clinical Trial Facility, Bagamoyo Research and Training Centre, Tanzania.

We report here the safety, reactogenicity and immunogenicity profile of vaccination with the RH5.1 protein ^14^ given with 50 µg Matrix-M™ adjuvant (Novavax) in monthly, delayed and delayed-fractional regimens up until 674 days (22 months) post-final study vaccination. The Consolidated Standards of Reporting Trials (CONSORT) guidelines were followed.

### Participants

Healthy adults (18-45 years) and infants (5-17 months) residing in Bagamoyo District, Tanzania, with a negative malaria blood film at screening, were eligible for inclusion in the study. Enrolment was into six groups (two adult and four infant groups) according to age, timing of enrolment and malaria pre-exposure status (infants only). A full list of inclusion and exclusion criteria are listed in the study protocol which is included within the Supplementary Appendix.

Each participant (or guardian) signed or thumb-printed an informed consent form at the in-person screening visit and consent was verified before each vaccination. The independent DSMB periodically reviewed the study progress and safety data according to a safety review schedule, including prior to each age de-escalation or dose escalation step.

## Procedures

### Safety Analysis

Following each vaccination, each participant was visited at home on days 1 (infants only), 3, 4, 5 and 6 by a community health worker for assessment and any solicited and unsolicited AEs were recorded. At days 1 (adults only), 2, 7, 14 and 28 post-vaccination, participants were seen at the clinical research facility. In addition, participants attended the facility for follow-up on days 84, 112, 309 and 674 post-third vaccination. Solicited AEs were collected at visits up to 7 days post-vaccination and unsolicited AEs were collected at visits up to 28 days post-vaccination. SAEs were collected at all visits. Observations (heart rate, temperature and blood pressure measurements) were taken at the clinic visits from the day of vaccination until day 84 post-third vaccination. Blood tests for exploratory immunology were taken at all visits except those occurring 2 days after each vaccination and 7 days after the first vaccination. Blood samples for safety (full blood count, alanine aminotransferase and creatinine) were carried out at screening and at all clinic visits except those occurring on days 1-2 post-vaccination and on 28 days post-second vaccination for the delayed regimen groups.

Solicited AEs were defined as being at least possibly related to vaccination. The likely causality and grading of unsolicited AEs and SAEs were assessed as described in the protocol. Unsolicited AEs were classified according to MedDRA (version 26.0).

### Outcomes

Primary outcome measures for vaccine safety included solicited and unsolicited AEs after each vaccination, along with any SAEs during the study period. The primary outcome analysis was conducted on the safety analysis population which included participants who received at least the first dose of vaccine in the study. Solicited AEs were collected for seven days after each study vaccination and the maximum severity reported for each solicited systemic and local AE by each participant after each vaccination was summarized by group. Unsolicited AEs were collected for 28 days after each study vaccination and those deemed at least possibly related to study vaccination are reported. SAEs were collected for the entire study period and all SAEs, regardless of causality, are reported.

The secondary outcome measures for immunogenicity were the concentration and avidity of anti-RH5.1 serum IgG antibodies by ELISA and their percentage GIA *in vitro* using purified IgG, as well as cellular immunogenicity to RH5.1 as measured by ELISPOT and/or (reported elsewhere) flow cytometry.

Exploratory outcome measures included the assessment of the frequency of RH5-specific plasma cells from bone marrow aspirates at baseline and 4 weeks post-third vaccinations, reported elsewhere ^24^.

### Vaccines

The design, production and preclinical testing of RH5.1 has been reported previously in detail ^13,14^. Briefly, RH5.1 is a soluble protein immunogen based on the full-length *P. falciparum* reticulocyte-binding protein homologue 5 (RH5). The protein is produced by secretion from *Drosophila melanogaster* Schneider-2 cells. RH5.1 was developed at the University of Oxford and an initial batch produced to Good Manufacturing Practice (GMP) by the Clinical Biomanufacturing Facility in Oxford.

Matrix-M™ adjuvant was manufactured by Novavax AB (Uppsala, Sweden). It is a potent, saponin-based adjuvant and comprises of partially-purified extracts of the bark of the *Quillaja saponaria* Molina tree, phosphatidylcholine and cholesterol. Matrix-M™ has demonstrated a favorable safety profile, as well as enhancement of cellular and humoral immune responses to a range of vaccine candidates and is licensed for human use in other malaria and Covid-19 vaccines ^3,22^.

### Peripheral Blood Mononuclear Cell (PBMC), Plasma and Serum Preparation

Blood samples were collected into lithium heparin-treated vacutainer blood collection systems (Becton Dickinson, UK). PBMC were isolated and used within 6 h in fresh assays as previously described ^34^. Excess cells were frozen in fetal calf serum (FCS) containing 10 % dimethyl sulfoxide and stored in liquid nitrogen. Plasma samples were stored at –80 °C. For serum preparation, using untreated vacutainers, blood samples were stored at room temperature and then the clotted blood was centrifuged for 5 min (1000 *xg*). Serum was stored at –80 °C.

### Anti-parasite Lysate ELISA

Serum antibody levels to parasite lysate were assessed by standardized ELISA methodology previously described ^12,16,35^. Schizont extract from *P. falciparum* (3D7 clone) parasites produced by the GIA Reference Laboratory, NIAID, NIH, was adsorbed overnight at 4 °C to 96 well NUNC-Immuno Maxisorp plates (Thermo Fisher Scientific) at an equivalent of 5×10^2^ parasites per µL. Test sera were diluted in 1 % milk in Dulbecco’s phosphate buffered saline (DPBS) and added in triplicate to plates following blocking with 5 % milk in DPBS (Sigma). A reference standard and internal control samples from a pool of N=34 “high” malaria pre-exposed serum samples, from the Bagamoyo Clinical Trial Facility-Ifakara Health Institute (BCTF-IHI) Biobank, plus blank wells, were included. Use of these samples received ethical approval from the Ifakara Health Institute Institutional Review Board (IHI/IRB/No: 49-2020). Bound antibodies were detected using goat anti-human IgG conjugated to alkaline phosphatase (Sigma), developed using 4-nitrophenyl phosphate disodium salt hexahydrate (Sigma) and absorbance at 405 nm (OD_405_) was determined on a BioTek Elx808 reader with Gen5 software. Antibody units were assigned using the reciprocal dilution of the standard giving an optical density of 1.0 at OD_405_. The standard curve and Gen5 software v3.04 (Agilent) were then used to convert the OD_405_ of test samples to arbitrary units (AU). “Low” and “high” malaria pre-exposure thresholds were first defined at <25^th^ percentile (20 AU) and >75% percentile (85 AU) of all N=105 population test samples. However, due to a large number of screening failures making recruitment difficult these were redefined as <75% percentile (71 AU) from “urban” wards, Magomeni and Dunda, and >60% percentile (140 AU) from “rural” wards, Fukayosi and Yombo, within Bagamoyo district. These criteria maintained a clear difference in anti-malarial antibody levels between the two cohorts.

### RH5 ELISA

Anti-RH5 total IgG ELISAs were performed against full-length RH5 protein (RH5.1) using standardized methodology as previously described ^10,12,15^. The reciprocal of the test sample dilution giving an OD_405_ of 1.0 in the standardized assay was used to assign an ELISA unit value of the standard. A standard curve and Gen5 ELISA software v3.04 (BioTek, UK) were used to convert the OD_405_ of individual test samples into AU. These responses in AU are reported in µg/mL following generation of a conversion factor by calibration-free concentration analysis as reported previously ^10^.

### Avidity ELISA

Anti-RH5.1 IgG antibody avidity was assessed by sodium thiocyanate (NaSCN)-displacement ELISA using previously described methodology ^10,15^. The concentration of NaSCN required to reduce the OD_405_ to 50% of that without NaSCN was used as a measure of avidity (IC_50_).

### Assay of Growth Inhibition Activity (GIA)

Standardized assays were performed by the GIA Reference Center, NIH, USA, using previously described methodology ^36^. Each sample was tested in three independent replication assays using three different batches of red blood cells (RBC), and the median of these three results was used to generate the final dataset as per previously reported RH5 vaccine trials and methodology ^12,15,37^. For each assay, in brief, protein G purified IgG samples were incubated with RBC infected with synchronized *P. falciparum* 3D7 clone parasites in a final volume of 40 µL for 40 h at 37 °C, and the final parasitemia in each well was quantified by biochemical determination of parasite lactate dehydrogenase. All purified IgG samples were tested initially at 10 mg/mL final test well concentration. For samples with >40 % GIA at 10 mg/mL, a dilution series was tested and used to determine the concentration that gave 50 % GIA (EC_50_).

### IFN-ɣ ELISPOT

Fresh PBMC were used in all assays using a previously described protocol ^10,12,15^. Spots were counted using an ELISPOT counter (Autoimmun Diagnostika (AID), Germany). Results are expressed as IFN-ɣ spot-forming units (SFU) per million PBMC with unstimulated control well responses subtracted from peptide-stimulated well measurements.

## Statistical Analysis

Data were analyzed using GraphPad Prism version 10.1 for Windows (GraphPad Software Inc., California, USA). All tests used were two-tailed and are described in the text. A value of *P* < 0.05 was considered significant.

## Author Contributions

Conceived and performed the experiments: SES, WFK, JS, IMM, FM, AD, CKB, NB, OH, CGM, SR, SM, NSL, BS, HM, MM, DMD, LM, GN, BM, OJ, TGM, IAS, RPM, JJK, JRB, LDWK, MMH, DP, CAL, CMN, KM, SJD, AIO, AMM.

Analyzed the data: SES, WFK, JS, TA, KM, SJD, AIO, AMM.

Project Management: SA, CC, AML, REC, FLN, RR, J-SC.

Wrote the paper: SES, JS, SJD, AMM.

## Data Availability

Requests for materials or data produced in the present study should be addressed to the corresponding author.

## Acknowledgments

The authors are grateful to Julie Furze, Jing Jin, Daniela Tomescu, Rebecca Ashfield, Kate Skinner, Susanne Hodgson, Cheryl Turner, Jack Quaddy, Subarna Sriskantharajah and Hilary Vitkus for assistance and Sally Pelling-Deeves for arranging contracts (University of Oxford); Richard Tarrant, Helena Parracho, Robert Smith and Catherine Green (Clinical Biomanufacturing Facility, University of Oxford); Jenny Reimer (Novavax); the independent DSMB for their insightful safety oversight (Brian Angus, Karim Manji, Roma Chilengi, Kwaku Poke Asante, William Macharia and Greg Fegan); and all the study participants.

This work was funded in part by the European and Developing Countries Clinical Trials Partnership (EDCTP) Multi-Stage Malaria Vaccine Consortium (MMVC) [RIA2016V-1649-MMVC]; an African Research Leader Award to AIO from the UK Medical Research Council (MRC) [MR/P020593/1], this award was jointly funded by the UK MRC and the UK Department for International Development (DFID) under the MRC/DFID Concordat agreement and is also part of the EDCTP2 programme supported by the European Union; the UK MRC [MR/K025554/1]; the National Institute for Health and Care Research (NIHR) Oxford Biomedical Research Centre (BRC), the views expressed are those of the authors and not necessarily those of the NHS, the NIHR, or the Department of Health; the GIA work was supported by the Division of Intramural Research, National Institute of Allergy and Infectious Diseases (NIAID) and by an Interagency Agreement (AID-GH-T-15-00001) between the United States Agency for International Development (USAID) Malaria Vaccine Development Program (MVDP) and NIAID. The findings and conclusions are those of the authors and do not necessarily represent the official position of USAID. CMN held a Wellcome Trust Sir Henry Wellcome Postdoctoral Fellowship [209200/Z/17/Z]. SJD is a Jenner Investigator.

## Data and Materials Availability

Requests for materials should be addressed to the corresponding authors.

## References

1. World Health O. World malaria report 2023. Geneva: World Health Organization; 2023.

2. Rts SCTP. Efficacy and safety of RTS,S/AS01 malaria vaccine with or without a booster dose in infants and children in Africa: final results of a phase 3, individually randomised, controlled trial. Lancet 2015; 386(9988): 31–45.

3. Datoo MS, Dicko A, Tinto H, et al. Safety and efficacy of malaria vaccine candidate R21/Matrix-M in African children: a multicentre, double-blind, randomised, phase 3 trial. Lancet 2024; 403(10426): 533–44.

4. Draper SJ, Sack BK, King CR, et al. Malaria Vaccines: Recent Advances and New Horizons. Cell Host Microbe 2018; 24(1): 43–56.

5. Scally SW, Triglia T, Evelyn C, et al. PCRCR complex is essential for invasion of human erythrocytes by Plasmodium falciparum. Nature microbiology 2022; 7(12): 2039–53.

6. Ragotte RJ, Higgins MK, Draper SJ. The RH5-CyRPA-Ripr Complex as a Malaria Vaccine Target. Trends Parasitol 2020; 36(6): 545–59.

7. Crosnier C, Bustamante LY, Bartholdson SJ, et al. Basigin is a receptor essential for erythrocyte invasion by Plasmodium falciparum. Nature 2011; 480(7378): 534–7.

8. Douglas AD, Baldeviano GC, Lucas CM, et al. A PfRH5-Based Vaccine Is Efficacious against Heterologous Strain Blood-Stage Plasmodium falciparum Infection in Aotus Monkeys. Cell Host Microbe 2015; 17(1): 130–9.

9. Douglas AD, Baldeviano GC, Jin J, et al. A defined mechanistic correlate of protection against Plasmodium falciparum malaria in non-human primates. Nat Commun 2019; 10(1): 1953.

10. Payne RO, Silk SE, Elias SC, et al. Human vaccination against RH5 induces neutralizing antimalarial antibodies that inhibit RH5 invasion complex interactions. JCI Insight 2017; 2(21): 96381.

11. Bliss CM, Drammeh A, Bowyer G, et al. Viral Vector Malaria Vaccines Induce High-Level T Cell and Antibody Responses in West African Children and Infants. Mol Ther 2017; 25(2): 547–59.

12. Silk SE, Kalinga WF, Mtaka IM, et al. Superior antibody immunogenicity of a viral-vectored RH5 blood-stage malaria vaccine in Tanzanian infants as compared to adults. Med 2023; 4(10): 668–86.

13. Hjerrild KA, Jin J, Wright KE, et al. Production of full-length soluble Plasmodium falciparum RH5 protein vaccine using a Drosophila melanogaster Schneider 2 stable cell line system. Sci Rep 2016; 6: 30357.

14. Jin J, Tarrant RD, Bolam EJ, et al. Production, quality control, stability, and potency of cGMP-produced Plasmodium falciparum RH5.1 protein vaccine expressed in Drosophila S2 cells. NPJ Vaccines 2018; 3: 32.

15. Minassian AM, Silk SE, Barrett JR, et al. Reduced blood-stage malaria growth and immune correlates in humans following RH5 vaccination. Med 2021; 2(6): 701–19.

16. Mwamlima TG, Mwakasungula SM, Mkindi CG, et al. Understanding the role of serological and clinical data on assessing the dynamic of malaria transmission: a case study of Bagamoyo district, Tanzania. Pan Afr Med J 2022; 43: 60.

17. Douglas AD, Williams AR, Illingworth JJ, et al. The blood-stage malaria antigen PfRH5 is susceptible to vaccine-inducible cross-strain neutralizing antibody. Nat Commun 2011; 2: 601.

18. Osier FH, Mackinnon MJ, Crosnier C, et al. New antigens for a multicomponent blood-stage malaria vaccine. Sci Transl Med 2014; 6(247): 247ra102.

19. Sang S, Datoo M, Otieno E, et al. Safety and immunogenicity of varied doses of R21/Matrix-M? vaccine at three years follow-up: A phase 1b age de-escalation, dose-escalation trial in adults, children, and infants in Kilifi-Kenya [version 1; peer review: awaiting peer review]. Wellcome open research 2023; 8(450).

20. Venkatraman N, Tiono AB, Bowyer G, et al. Phase I assessments of first-in-human administration of a novel malaria anti-sporozoite vaccine candidate, R21 in matrix-M adjuvant, in UK and Burkinabe volunteers. medRxiv 2019: 19009282.

21. https://www.nbs.go.tz/nbs/takwimu/census2012/Mortality_and_Health_Monograph.pdf.

22. Heath PT, Galiza EP, Baxter DN, et al. Safety and Efficacy of NVX-CoV2373 Covid-19 Vaccine. N Engl J Med 2021; 385(13): 1172–83.

23. Nielsen CM, Ogbe A, Pedroza-Pacheco I, et al. Protein/AS01B vaccination elicits stronger, more Th2-skewed antigen-specific human T follicular helper cell responses than heterologous viral vectors. Cell Rep Med 2021; 2(3): 100207.

24. Barrett JR, Silk SE, Mkindi CG, et al. Analyses of human vaccine-specific circulating and bone marrow-resident B cell populations reveal benefit of delayed vaccine booster dosing with blood-stage malaria antigens. Front Immunol 2023; 14: 1193079.

25. Nielsen CM, Barrett JR, Davis C, et al. Delayed boosting improves human antigen-specific Ig and B cell responses to the RH5.1/AS01B malaria vaccine. JCI Insight 2023; 8(2).

26. McNamara HA, Idris AH, Sutton HJ, et al. Antibody Feedback Limits the Expansion of B Cell Responses to Malaria Vaccination but Drives Diversification of the Humoral Response. Cell Host Microbe 2020; 28(4): 572–85 e7.

27. Regules JA, Cicatelli SB, Bennett JW, et al. Fractional Third and Fourth Dose of RTS,S/AS01 Malaria Candidate Vaccine: A Phase 2a Controlled Human Malaria Parasite Infection and Immunogenicity Study. J Infect Dis 2016; 214(5): 762–71.

28. Samuels AM, Ansong D, Kariuki SK, et al. Efficacy of RTS,S/AS01(E) malaria vaccine administered according to different full, fractional, and delayed third or early fourth dose regimens in children aged 5-17 months in Ghana and Kenya: an open-label, phase 2b, randomised controlled trial. Lancet Infect Dis 2022; 22(9): 1329–42.

29. Rts SCTP. Efficacy and safety of the RTS,S/AS01 malaria vaccine during 18 months after vaccination: a phase 3 randomized, controlled trial in children and young infants at 11 African sites. PLoS Med 2014; 11(7): e1001685.

30. Leroux-Roels G, Leroux-Roels I, Clement F, et al. Evaluation of the immune response to RTS,S/AS01 and RTS,S/AS02 adjuvanted vaccines: randomized, double-blind study in malaria-naive adults. Hum Vaccin Immunother 2014; 10(8): 2211–9.

31. Barrett JR, Pipini D, Wright ND, et al. Analysis of the Diverse Antigenic Landscape of the Malaria Invasion Protein RH5 Identifies a Potent Vaccine-Induced Human Public Antibody Clonotype. bioRxiv 2023: 2023.10.04.560576.

32. Miura K, Perera S, Brockley S, et al. Non-apical membrane antigen 1 (AMA1) IgGs from Malian children interfere with functional activity of AMA1 IgGs as judged by growth inhibition assay. PLoS One 2011; 6(6): e20947.

33. Willcox AC, Huber AS, Diouf A, et al. Antibodies from malaria-exposed Malians generally interact additively or synergistically with human vaccine-induced RH5 antibodies. Cell Rep Med 2021; 2(7): 100326.

34. Sheehy SH, Duncan CJ, Elias SC, et al. Phase Ia Clinical Evaluation of the Safety and Immunogenicity of the Plasmodium falciparum Blood-Stage Antigen AMA1 in ChAd63 and MVA Vaccine Vectors. PLoS One 2012; 7(2): e31208.

35. Miura K, Orcutt AC, Muratova OV, Miller LH, Saul A, Long CA. Development and characterization of a standardized ELISA including a reference serum on each plate to detect antibodies induced by experimental malaria vaccines. Vaccine 2008; 26(2): 193–200.

36. Malkin EM, Diemert DJ, McArthur JH, et al. Phase 1 clinical trial of apical membrane antigen 1: an asexual blood-stage vaccine for Plasmodium falciparum malaria. Infect Immun 2005; 73(6): 3677–85.

37. Miura K, Diouf A, Fay MP, et al. Assessment of precision in growth inhibition assay (GIA) using human anti-PfRH5 antibodies. Malar J 2023; 22(1): 159.

