## Supplemental Information for "First Phase 1b, single-center, age de-escalation trial of the *P. falciparum* blood-stage malaria vaccine candidate RH5.1/Matrix-M^™^: a delayed boost regimen induces high levels of functional antibodies in 5-17 month old Tanzanian infants"

#### Supplementary Figures

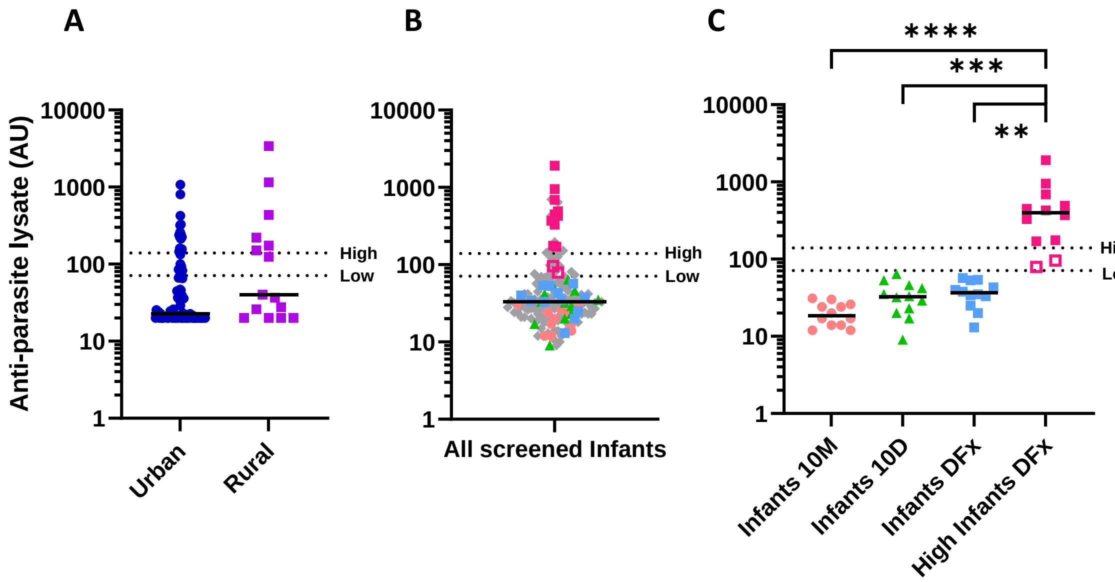

**Figure S1. Malaria pre-exposure assessment at recruitment.**

Median and individual anti-parasite lysate serum total IgG responses, as measured by ELISA in arbitrary units (AU) and using samples from the BCTF-IHI Biobank, are shown for (**A**) 5-17month old infants from low malaria incidence “urban” (N=90) and high malaria incidence “rural” (N=15) wards within Bagamoyo district. (**B**) All 5-17 month old infants in Bagamoyo district screened for the VAC080 study (N=156), with participants subsequently recruited into groups indicated by respective colours from panel (**C**). (**C**) All four infant cohorts in the VAC080 study (“Infants 10M” N=12; “Infants 10D” N=12; “Infants DFx” N=12; and “High Infants DFx” N=12. Open symbols indicate N=2 volunteers in “High Infants DFx” who did not meet high pre-exposure threshold. Upper dotted line indicates high malaria pre-exposure threshold (140 AU) for recruitment into “High Infants DFx”. Lower dotted line indicates low malaria pre-exposure threshold (71 AU) for recruitment into “Infants 10M”, “Infants 10D” and “Infants DFx”.

| Study group | | Adults 10M | Adults DFx | Infants 10M | Infants 10D | Infants DFx | High Infants DFx |
|---|---|---|---|---|---|---|---|
| **No. of participants** | | 6 | 6 | 12 | 12 | 12 | 12 |
| **Sex** | Female | 0 (0%) | 0 (0%) | 8 (75%) | 4 (25%) | 4 (25%) | 8 (75%) |
| | Male | 6 (100%) | 6 (100%) | 4 (25%) | 8 (75%) | 8 (75%) | 4 (25%) |
| **Age at screening** | Median | 25.5 yrs | 21 yrs | 12 mths | 14.5 mths | 8.5 mths | 11.5 mths |
| | Range | 23-29 yrs | 20-30 yrs | 6-15 mths | 5-17 mths | 5-15 mths | 5-15 mths |
| **Literate** | Yes | 6 (100%) | 6 (100%) | N/A | N/A | N/A | N/A |
| | No | 0 (0%) | 0 (0%) | | | | |
| **Education** | Primary | 3 (50%) | 1 (17%) | N/A | N/A | N/A | N/A |
| | Secondary | 3 (50%) | 5 (83%) | | | | |
| | Tertiary | 0 (0%) | 0 (0%) | | | | |

**Figure S2: Demographic information for study participants.**

Numbers are number of participants (%) unless otherwise indicated. 10M = monthly vaccination regimen; DFx = delayed-fractional regimen; 10D = delayed regimen; N/A = not applicable; yrs = years; mths = months.

| Age group | MedDRA System Organ Class | MedDRA Higher Level Term | MedDRA Preferred Term | Group | Onset (days post-vaccination) | Duration (days) | Maximum severity |
|---|---|---|---|---|---|---|---|
| Adults | Nervous system disorders | Neurological signs and symptoms NEC | Dizziness | Adults 10M | V3+6 | 0 | Mild |
| Infants | Gastrointestinal disorders | Gastrointestinal infections, site unspecified | Helminthic infection | Infants 10D | V2+14 | 53 | Mild |
| | | | Diarrhoea infectious | Infants 10M | V1+9 | 2 | Mild |
| | General disorders and administration site conditions | Febrile disorders | Pyrexia | Infants 10D | V2+1 | 2 | Mild |
| | Respiratory, thoracic and mediastinal disorders | Upper respiratory tract infections NEC | Upper respiratory tract infection | Infants DFx | V1+2 | 8 | Mild |
| | Skin and subcutaneous tissue disorders | Dermatitis and eczema | Eczema | High Infants DFx | V2+4 | 3 | Moderate |

**Figure S3: Unsolicited adverse events (AEs).**

Unsolicited AEs were collected for 28 days following each study vaccination. Unsolicited AEs deemed at least possibly (possibly, probably or definitely) related to study interventions in all participants are shown, with MedDRA coding and maximum severity reported. 10M = monthly vaccination regimen; DFx = delayed-fractional regimen; 10D = delayed regimen; VX+Y = Y number of days post study vaccination X.

| SAE number | Participant group | Participant age at onset (months) | Participant sex | SAE description | SAE category | SAE onset (days post V3) | SAE duration (days) | SAE relationship to vaccine | SAE outcome |
|---|---|---|---|---|---|---|---|---|---|
| 1 | Infants DFx | 16 | Male | Stage IV neuroblastoma | Resulted in death | 91 | 142 | Not related | Death |
| 2 | Infants 10D | 25 | Male | Febrile convulsion | Hospitalization | 119 | 2 | Not related | Resolved |
| 3 | Infants DFx | 18 | Female | Acute respiratory failure | Resulted in death | 126 | 0 | Not related | Death |
| 4 | Infants 10D | 29 | Male | Severe pneumonia | Hospitalization | 237 | 73 | Not related | Resolved |
| 5 | Infants 10D | 33 | Male | Febrile convulsion | Hospitalization | 356 | 19 | Not related | Resolved |

**Figure S4: Serious adverse events (SAEs).**

All SAEs occurring during the VAC080 study period are shown. No SAEs were deemed related to the study vaccinations. 10M = monthly vaccination regimen; DFx = delayed-fractional regimen; 10D = delayed regimen.

SAE 1 occurred in a 16 month old child who was diagnosed with neuroblastoma. The child was initially found to be newly anemic just prior to their third study vaccination. This was 4 months after receiving their second study vaccination. They were commenced on iron supplements and, as their hemoglobin remained stable and they were well, the decision was made to administer the third vaccination a week later. Two months later, they became unwell with a febrile illness and were found to have worsening anemia and lymphadenopathy. They were referred for further investigations at the district and then tertiary hospital. Investigations confirmed a diagnosis of stage IV neuroblastoma and the child was transferred to the oncology ward. The child received a blood transfusion to correct their anemia and chemotherapy was commenced shortly thereafter. Three days after commencing chemotherapy, the child developed fevers and was diagnosed with severe malaria. They were initially treated with intravenous (IV) artesunate and later switched to IV artemether-lumefantrine. The child recovered from this episode of malaria; however, despite commencing chemotherapy, the child later died. This SAE was deemed not related to vaccination by both the local site and the Sponsor. The

DSMB was also informed.

SAE 2 occurred in a 2 year old child who developed a febrile convulsion secondary to pneumonia and required hospitalization for two days. After discharge the child completed a five day course of oral antibiotics. The child developed further fever and cough two days after completing antibiotics, so was taken to the nearest health facility where they were treated with further IV antibiotics. The child's condition improved after recommencing IV antibiotics and the child was subsequently discharged after completion of treatment. This SAE occurred three months after receiving their final study vaccination and was deemed not related to vaccination.

SAE 3 occurred in an 18 month old child who died after developing acute respiratory failure following local herb ingestion. The participant experienced sudden greenish vomiting and two episodes of passing greenish loose stool a few hours prior to death and during resuscitation greenish aspirates were noted. Herbal intoxication was presumed as the underlying cause of death due to the clinical presentation, however, an autopsy could not be performed in order to confirm the cause of death due to the child's parents' cultural beliefs. There was no temporal relation between this event and the study vaccinations; the child received their final study vaccination four months previously. This SAE was deemed not related to vaccination by the local site, the Sponsor and the DSMB.

SAE 4 occurred in a 2 year old child who was hospitalized with suspected pneumonia after presenting acutely unwell with fever, difficulty breathing, vomiting and diarrhea. The child was treated with IV antibiotics and oxygen therapy after which their condition improved. After four days of inpatient treatment, the child was discharged home to complete a seven day course of oral antibiotics. The child was followed up as an outpatient for three months due to persisting hematological abnormalities (leukocytosis and neutrophilia), however, these findings eventually normalized without additional treatment. As the child received their final study vaccination three months prior to the start of these events, this SAE was deemed not related to vaccination.

SAE 5 occurred in the same child as SAE 2 eight months later. The child was admitted to hospital with dry cough, fever, vomiting and loose stools. Upon arrival to hospital, the child experienced several febrile convulsions. The child was diagnosed with febrile convulsions secondary to sepsis and treated with IV antibiotics. The child was discharged from hospital two days later with oral antibiotics following improvement in their cough and resolution of their other presenting symptoms. This second SAE occurred eleven months after the participant received their final study vaccination and was deemed not related to vaccination.

**A**

| Laboratory abnormality | Adults 10M | | | | Adults DFx | | | |
|---|---|---|---|---|---|---|---|---|
| | No. of participants | Grade | | | No. of participants | Grade | | |
| | | 1 | 2 | 3 | | 1 | 2 | 3 |
| Leukopenia | 1 (17%) | 1 (17%) | 0 (0%) | 0 (0%) | 1 (17%) | 1 (17%) | 0 (0%) | 0 (0%) |
| Thrombocytopenia | 1 (17%) | 1 (17%) | 0 (0%) | 0 (0%) | 0 (0%) | 0 (0%) | 0 (0%) | 0 (0%) |
| Neutropenia | 1 (17%) | 1 (17%) | 0 (0%) | 0 (0%) | 1 (17%) | 1 (17%) | 0 (0%) | 0 (0%) |
| Lymphopenia | 1 (17%) | 1 (17%) | 0 (0%) | 0 (0%) | 2 (33%) | 2 (33%) | 0 (0%) | 0 (0%) |
| Eosinophilia | 1 (17%) | 1 (17%) | 0 (0%) | 0 (0%) | 0 (0%) | 0 (0%) | 0 (0%) | 0 (0%) |
| Elevated alanine aminotransaminase | 2 (33%) | 2 (33%) | 0 (0%) | 0 (0%) | 2 (33%) | 2 (33%) | 0 (0%) | 0 (0%) |

**B**

| Laboratory abnormality | Infants 10M | | | | Infants 10D | | | |
|---|---|---|---|---|---|---|---|---|
| | No. of participants | Grade | | | No. of participants | Grade | | |
| | | 1 | 2 | 3 | | 1 | 2 | 3 |
| Anemia | 7 (58%) | 5 (42%) | 2 (17%) | 0 (0%) | 7 (58%) | 5 (42%) | 2 (17%) | 0 (0%) |
| Leukocytosis | 5 (42%) | 5 (42%) | 0 (0%) | 0 (0%) | 4 (33%) | 4 (33%) | 0 (0%) | 0 (0%) |
| Leukopenia | 0 (0%) | 0 (0%) | 0 (0%) | 0 (0%) | 0 (0%) | 0 (0%) | 0 (0%) | 0 (0%) |
| Thrombocytopenia | 0 (0%) | 0 (0%) | 0 (0%) | 0 (0%) | 0 (0%) | 0 (0%) | 0 (0%) | 0 (0%) |
| Neutropenia | 4 (33%) | 4 (33%) | 0 (0%) | 0 (0%) | 5 (42%) | 2 (17%) | 1 (8%) | 1 (8%) |
| Eosinophilia | 0 (0%) | 0 (0%) | 0 (0%) | 0 (0%) | 1 (8%) | 1 (8%) | 0 (0%) | 0 (0%) |
| Elevated creatinine | 0 (0%) | 0 (0%) | 0 (0%) | 0 (0%) | 1 (8%) | 0 (0%) | 1 (8%) | 0 (0%) |
| Elevated alanine aminotransaminase | 1 (8%) | 1 (8%) | 0 (0%) | 0 (0%) | 1 (8%) | 1 (8%) | 0 (0%) | 0 (0%) |

| Laboratory abnormality | Infants DFx | | | | High Infants DFx | | | |
|---|---|---|---|---|---|---|---|---|
| | No. of participants | Grade | | | No. of participants | Grade | | |
| | | 1 | 2 | 3 | | 1 | 2 | 3 |
| Anemia | 7 (58%) | 3 (25%) | 4 (33%) | 0 (0%) | 5 (42%) | 2 (17%) | 3 (25%) | 0 (0%) |
| Leukocytosis | 3 (25%) | 3 (25%) | 0 (0%) | 0 (0%) | 4 (33%) | 4 (33%) | 0 (0%) | 0 (0%) |
| Leukopenia | 0 (0%) | 0 (0%) | 0 (0%) | 0 (0%) | 1 (8%) | 1 (8%) | 0 (0%) | 0 (0%) |
| Thrombocytopenia | 1 (8%) | 0 (0%) | 1 (8%) | 0 (0%) | 0 (0%) | 0 (0%) | 0 (0%) | 0 (0%) |
| Neutropenia | 3 (25%) | 3 (25%) | 0 (0%) | 0 (0%) | 0 (0%) | 2 (17%) | 0 (0%) | 1 (8%) |
| Eosinophilia | 1 (8%) | 1 (8%) | 0 (0%) | 0 (0%) | 5 (42%) | 2 (17%) | 1 (8%) | 1 (8%) |
| Elevated creatinine | 3 (25%) | 3 (25%) | 0 (0%) | 0 (0%) | 0 (0%) | 0 (0%) | 0 (0%) | 0 (0%) |
| Elevated alanine aminotransaminase | 2 (17%) | 2 (17%) | 0 (0%) | 0 (0%) | 0 (0%) | 0 (0%) | 0 (0%) | 0 (0%) |

#### Figure S5: Laboratory adverse events (AEs).

All participants reporting mild (grade 1) or higher severity laboratory abnormalities at least possibly related to study vaccination are shown. (**A**) Adults (aged 18-45 years; N=6 for each group). (**B**) Infants (aged 5-17 months; N=12 for each group). 10M = monthly vaccination regimen; DFx = delayed-fractional regimen; 10D = delayed regimen; Grade 1 = mild; Grade 2 = moderate; Grade 3 = severe.

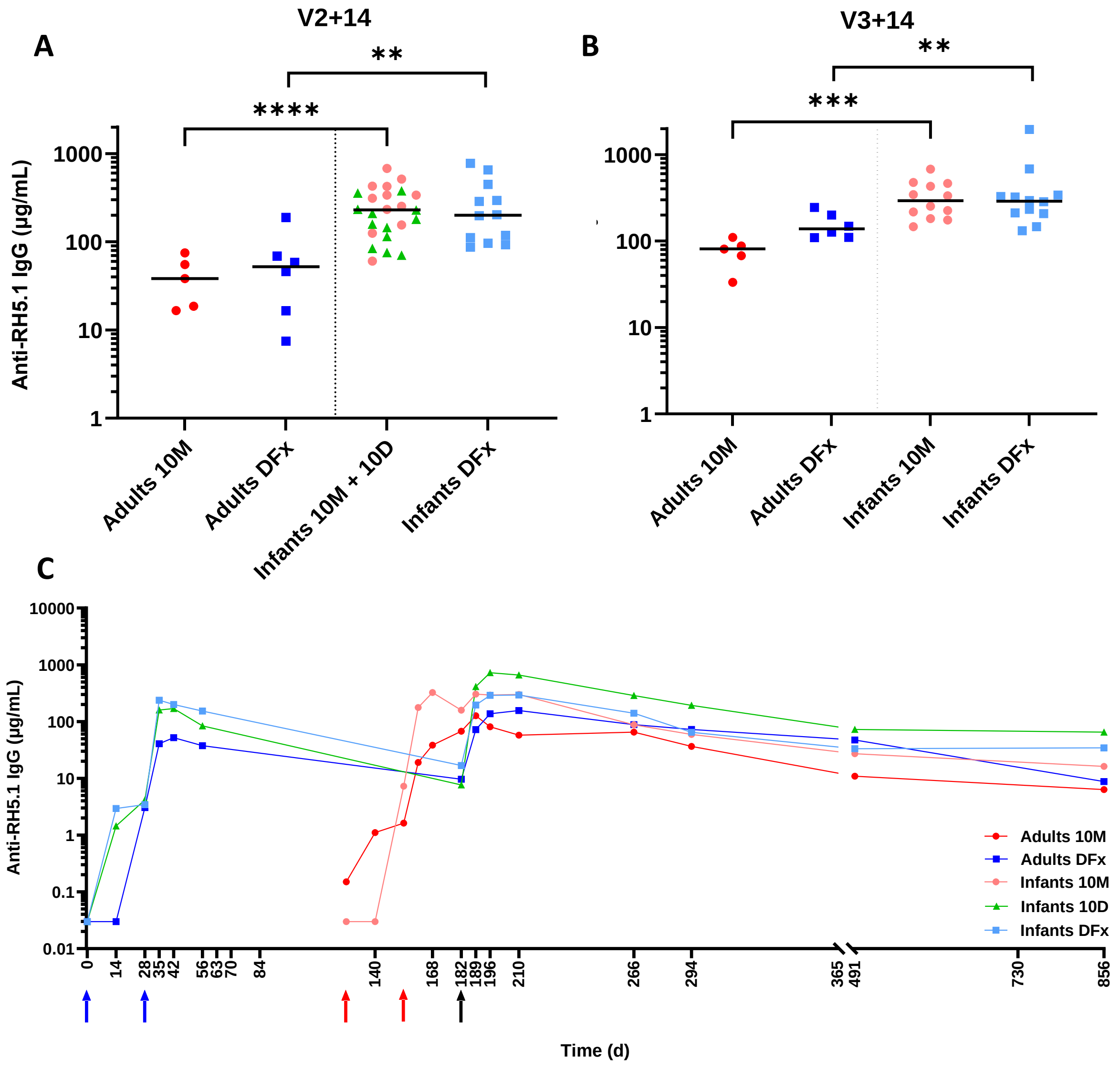

**Figure S6: Serum antibody response and kinetic following vaccination.**

(**A**) Median and individual anti-RH5.1 serum total IgG responses as measured by ELISA are shown at 14 days post-second vaccination (V2+14) for "Adults 10M" (N=5), "Adults DFx" (N=6), "Infants 10M+10D" (combined due to identical immunization and dosing regimen at this time-point; N=24) and "Infants DFx" (N=12); and at (**B**) 14 days post-third vaccination (V3+14), with same groups as in (A) except "Infants 10M" (N=12). Analyses used Mann-Whitney test to compare between the same dosing regimens across age groups, ***P* < 0.01, ****P* < 0.001, *****P*< 0.0001. (**C**) Median anti-RH5.1 serum total IgG responses as measured by ELISA are shown over time for all groups in the

VAC080 trial, except “High Infants DFx”. “Adults 10M” (N=5-6); “Infants 10M” (N=12-15); “Adults DFx” (N=6); “Infants DFx” (N=11-12); and “Infants 10D” (N=11-12). Data for all groups are aligned by third vaccination timepoint: delayed regimens (“Adult DFx”, “Infant DFx” and “Infant 10D”) vaccinations 1 and 2 are indicated by blue arrows (day 0 for dose 1 and day 28 for dose 2); monthly (“Adult 10M”, “Infant 10M”) vaccinations 1 and 2 are indicated by red arrows (day 0 for dose 1 aligned to day 126 and day 28 for dose 2 aligned to day 154); third vaccination for all groups indicated by black arrow at day 182 (representing day 56 for monthly groups).

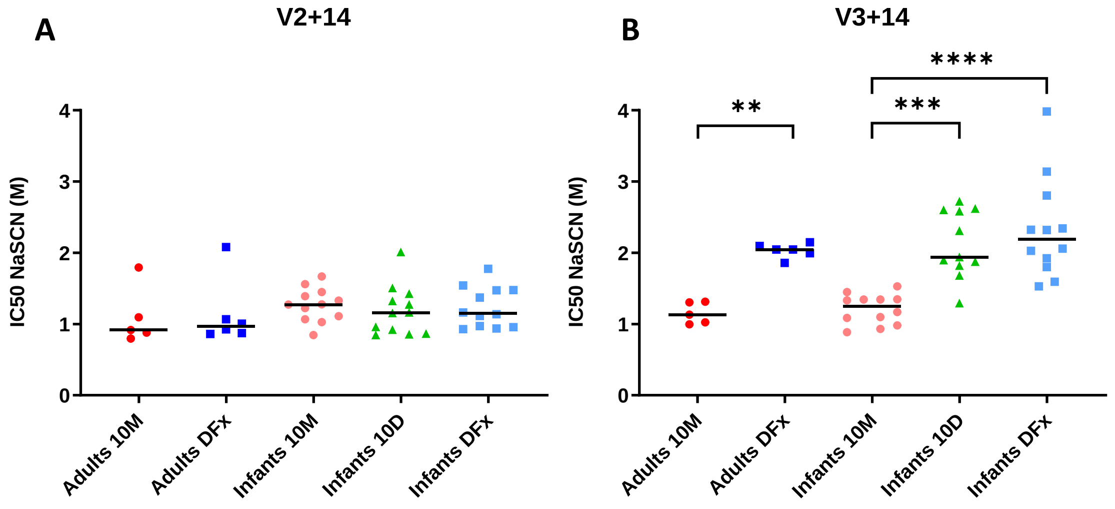

**Figure S7: Avidity of serum antibody responses.**

Avidity of serum anti-RH5.1 total IgG responses was assessed by NaSCN displacement ELISA at (**A**) 14 days post-second vaccination (V2+14 / day 42) and (**B**) 14 days post-third vaccination (V3+14 / day 70 for monthly regimens / day 196 for delayed regimens. "Adults 10M" (N=5); "Infants 10M" (N=12); "Adults DFx" (N=6); "Infants DFx" (N=12); and "Infants 10D" (N=11-12). Avidity is reported as the molar concentration of NaSCN required to reduce the starting optical density (OD) in the ELISA by 50 % ($IC_{50}$). Analyses used Mann-Whitney test between the two adult groups, $^{**}P < 0.01$; and Kruskal-Wallis with Dunn's multiple comparison test between the three infant groups, $^{**}P < 0.01$, $^{***}P < 0.001$, $^{****}P < 0.0001$.

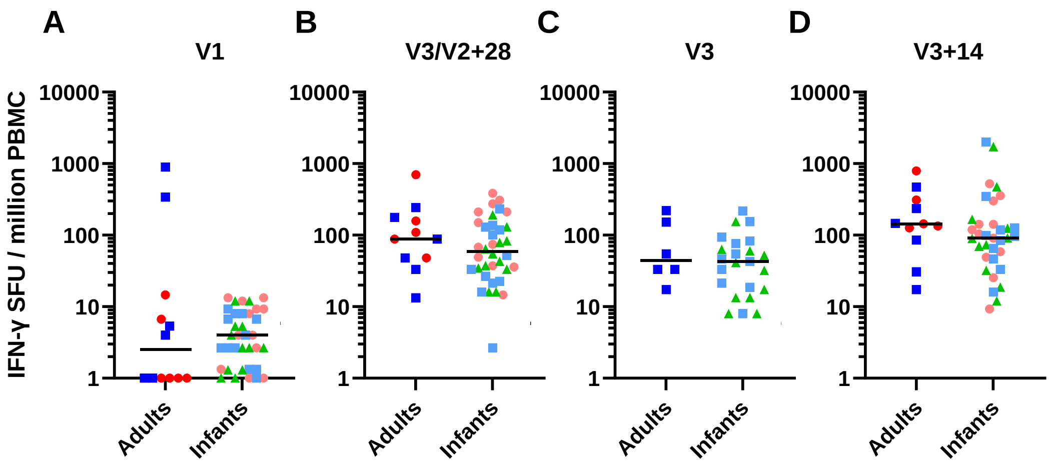

**Figure S8: *Ex vivo* IFN-γ T cell response to RH5.1 vaccination.**

Median and individual *ex vivo* IFN-γ enzyme-linked immunospot (ELISPOT) responses to RH5 per million peripheral blood mononuclear cells (PBMC) are shown for all groups in the VAC080 trial (except "High Infants DFx"). Responses are shown at: (**A**) baseline, day of first vaccination (V1), Adults (N=12) and Infants (N=36); (**B**) 28 days post-second vaccination (V2+28 = day 56 and day of third vaccination (V3) for monthly regimen groups), Adults (N=11) and Infants (N=36); (**C**) V3 for delayed regimen groups (day 182), Adults (N=6) and Infants (N=23); and (**D**) 14 days post-third vaccination (V3+14 = day 70 for monthly regimens and day 196 for delayed regimens, Adults (N=11) and Infants (N=35). Individual groups within each age cohort are color-coded as per **Figure 3**. Negative responses at V1 are plotted at 1 IFN-γ SFU / million PBMC. Analyses using Mann-Whitney test to compare between adult and infant responses at each timepoint showed no significant differences.

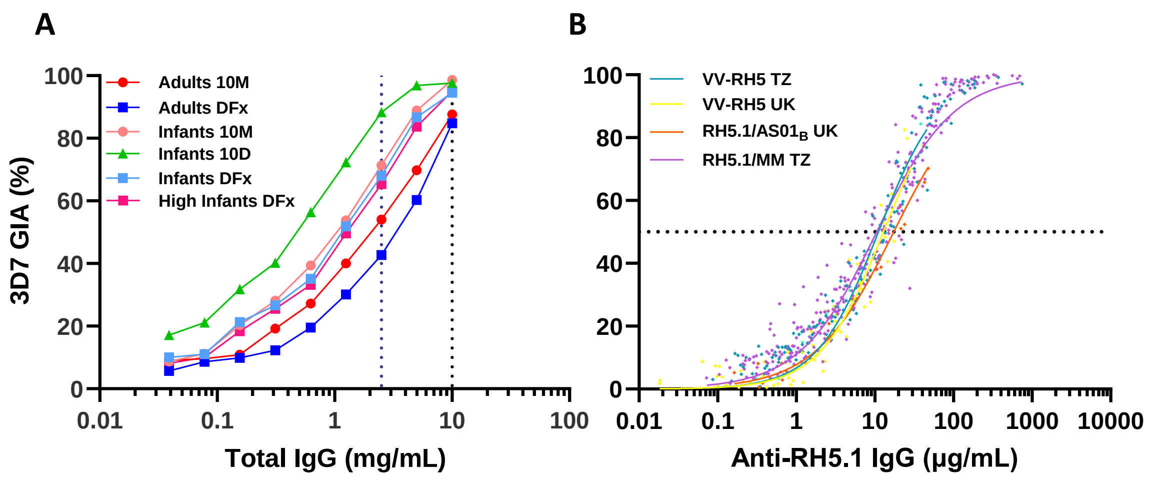

**Figure S9: Functional GIA induced by RH5 vaccination.**

*In vitro* GIA of total IgG purified from serum was assessed against 3D7 clone *P. falciparum* parasites. (**A**) V3+14 samples (Day 70 for monthly regimens: "Adults 10M" N=3; "Infants 10M" N=12; and Day 196 for delayed regimens: "Adults DFx" N=6; "Infants 10D" N=11; "Infants DFx" N=12; and "High Infants DFx" N=10) were titrated in the GIA assay using a 2-fold dilution series and starting at 10 mg/mL total IgG. Group medians are plotted. Vertical dashed lines indicate GIA at 10 mg/mL and 2.5 mg/mL total IgG as shown in **Figure 4**. (**B**) Relationship between GIA and concentration of anti-RH5.1-specific IgG used in the assay as measured by ELISA in the total purified IgG is shown for samples from the VAC080 trial using RH5.1/Matrix-M™ (RH5.1/MM) in healthy Tanzanian (TZ) adults and infants (all GIA datapoints plotted from N=54 participants). Previously published GIA assay data from three other clinical trials (reported in [1]) are shown for comparison using viral vectored RH5 (VV-RH5) vaccines in healthy UK adults (data from N=16 participants) or healthy Tanzanian (TZ) adults and infants (data from N=19 participants), or the RH5.1/AS01$_B$ protein-in-adjuvant formulation in healthy UK adults (data from N=8 participants) [1-3]. Non-linear regression curves (constrained to >0 % and <100 % GIA) are shown for each group. Dotted line indicates 50 % GIA.

**A**

| | Median GIA (%) | N = | Proportion >60% GIA (%) |
|---|---|---|---|
| **Infants 10M** | 71.4 | 12 | 75 |
| **Infants 10D** | 88.3 | 11 | 100 |
| **Infants DFx** | 68.0 | 12 | 67 |
| **High Infants DFx** | 65.4 | 10 | 70 |

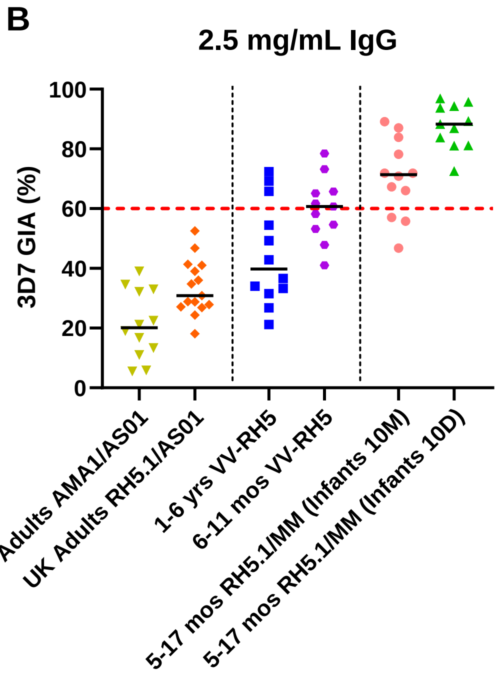

**Figure S10: Comparison of functional GIA across vaccines, regimens and populations.** (**A**) Summary table of GIA data at the V3+14 timepoint from the four infant cohorts in the VAC080 trial as tested using 2.5 mg/mL purified total IgG. The % proportion of infants in each group measuring over a target threshold of 60 % GIA is indicated. This level of GIA was previously associated with protective outcome in RH5-vaccinated and *P. falciparum* challenged *Aotus* monkeys [4,5]. (**B**) Comparison of *in vitro* GIA against 3D7 clone *P. falciparum* parasites, across different blood-stage malaria vaccines in different populations, tested at the sub-physiological concentration of 2.5 mg/mL total IgG purified from serum 1-2 weeks following the final immunization. Data reproduced from this and previous clinical studies. The AMA1/AS01 and RH5.1/AS01 vaccines were previously tested in healthy UK adults including efficacy assessment by blood-stage *P. falciparum* CHMI [3,6]. The viral vectored RH5 (VV-RH5) vaccine was previously tested in healthy Tanzanian children 1-6 years and 6-11 months of age [1]. The RH5.1/Matrix-M™ vaccine (RH5.1/MM) was tested in healthy Tanzanian infants 5-17 months of age in the study reported here. Individual and median responses are shown. Tanzanian infants (5-17 months of age) immunized with RH5.1/MM in the "Infants 10D" regimen show the highest vaccine-induced GIA in humans to date, all over the target threshold of 60 % GIA.

#### Supplementary References

1. Silk SE, Kalinga WF, Mtaka IM, et al. Superior antibody immunogenicity of a viral-vectored RH5 blood-stage malaria vaccine in Tanzanian infants as compared to adults. *Med* 2023; **4**(10): 668-86.
2. Payne RO, Silk SE, Elias SC, et al. Human vaccination against RH5 induces neutralizing antimalarial antibodies that inhibit RH5 invasion complex interactions. *JCI Insight* 2017; **2**(21): 96381.
3. Minassian AM, Silk SE, Barrett JR, et al. Reduced blood-stage malaria growth and immune correlates in humans following RH5 vaccination. *Med* 2021; **2**(6): 701-19.
4. Douglas AD, Baldeviano GC, Lucas CM, et al. A PfRH5-Based Vaccine Is Efficacious against Heterologous Strain Blood-Stage Plasmodium falciparum Infection in Aotus Monkeys. *Cell Host Microbe* 2015; **17**(1): 130-9.
5. Douglas AD, Baldeviano GC, Jin J, et al. A defined mechanistic correlate of protection against Plasmodium falciparum malaria in non-human primates. *Nat Commun* 2019; **10**(1): 1953.
6. Payne RO, Milne KH, Elias SC, et al. Demonstration of the Blood-Stage Controlled Human Malaria Infection Model to Assess Efficacy of the Plasmodium falciparum AMA1 Vaccine FMP2.1/AS01. *J Infect Dis* 2016; **213**(11): 1743-51.

#### Supplementary Appendix – VAC080 Trial Protocol

**Trial Title:** A Phase Ib clinical trial to assess the safety and immunogenicity of the blood-stage *Plasmodium falciparum* malaria vaccine candidate RH5.1/Matrix-M in healthy adults and infants in Tanzania.

**Short title:** Safety and Immunogenicity of RH5.1/Matrix-M in adults and infants living in Tanzania.

| | |
|---|---|
| Study reference: | VAC080 |
| Protocol Version: | 3.0 |
| Date: | 14th April 2021 |
| OXTREC Number | 9-20 |
| Sponsor: | University of Oxford |
| Funding body | EDCTP |
| Authors | Ally Olotu, Angela M Minassian, Simon J Draper |

#### Statement of Compliance

I have read this protocol, and I agree to abide by all provisions set forth herein. I agree to comply with the principles of the International Conference on Harmonization Tripartite Guideline on Good Clinical Practice (GCP).

#### Confidentiality Statement

This document contains confidential information that must not be disclosed to anyone other than the trial Sponsor, the Investigator Team, and members of the Ethics Committees and Regulatory Authorities. This information cannot be used for any purpose other than the evaluation or conduct of the clinical investigation without the prior written consent of the principal investigator.

#### Signature Page

| Role | Name | Signature | Date |
|---|---|---|---|
| Principal Investigator | Dr Ally Olotu | 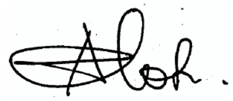 | 10/05/2021 |
| UK Senior Laboratory Investigator | Prof Simon J Draper | 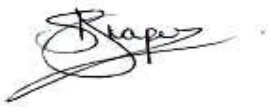 | 07/05/2021 |
| Chief Investigator | Dr Angela M Minassian | 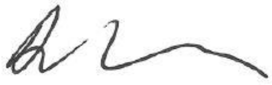 | 10/05/2021 |

#### Confidentiality Statement

This document contains confidential information that must not be disclosed to anyone other than the Sponsor, the Investigator Team, the Regulatory Authority and members of the Independent Ethics Committee. This information cannot be used for any purpose other than the evaluation or conduct of the clinical investigation without the prior written consent of Dr. Angela Minassian or Professor Simon Draper.

#### Conflict of Interest

**1.** "According to the Declaration of Helsinki, 2008, I have read this protocol, and declare the following conflict of interest"

Details: I have a family member who is an inventor on patents for RH5-based vaccines.

| | | |
|---|---|---|
| | | 10/05/2021 |
| **Chief Investigator** | **Investigator Signature** | **Date** |

**Dr Angela M. Minassian**

**2.** "According to the Declaration of Helsinki, 2008, I have read this protocol, and declare the following conflict of interest"

Details: I am a named inventor on patents relating to RH5-based vaccines.

| | | |
|---|---|---|
| | | 07/05/2021 |
| **Principal Investigator** | **Investigator Signature** | **Date** |

**Professor Simon J. Draper**

**3.** "According to the Declaration of Helsinki, 2008, I have read this protocol, and declare no conflict of interest"

Details:

| | | |
|---|---|---|
| | | 10/05/2021 |
| **Principal Investigator** | **Investigator Signature** | **Date** |

**Dr Ally Olotu**

#### 2. Figures

Figure 10: (A): qPCR data for the VAC063A Phase IIa study; Group 5 vaccinees (n=14) and Group 6 (n=15). Median parasitemia is shown over time for each group. The lower limit of quantification is indicated by the dotted line at 20 p/mL. Time=days (d) post blood-stage CHMI. (B) Primary efficacy endpoint analysis of PMR, showing each individual, plus the mean. Both datasets are normally distributed (D'Agostino-Pearson test); * P=0.031 using two-tailed t-test with Welch's correction for non-equal variances (F-test; P=0.008). (C): Secondary efficacy endpoint analysis

of PMR, showing each individual, plus the median for the VAC063B Phase IIa study. Group 7 (n=9), Group 8 (n=8) and Group 9 (n=6). * P=0.022; Kruskal-Wallis test with Dunn's multiple comparison test........................................................................................................33
Figure 5 : Study groups and schedule........................................................................43
Figure 6: Example of Vaccine labels ........................................................................77

#### 3. Tables

#### 4. KEY ROLES AND GENERAL INFORMATION

| | |
|---|---|
| **Trial Centre** | Ifakara Health Institute Clinical Trial Facility,<br>Bagamoyo Research and Training Centre<br>P.O. Box 74<br>Bagamoyo, Tanzania |
| **Principal Investigator (IHI):** | Ally Olotu MD DPhil<br>Ifakara Health Institute<br>P.O. Box 74<br>Bagamoyo, Tanzania<br>Tel: +255 718 927 104<br> |
| **UK Senior Laboratory Investigator (UOXF):** | Prof Simon J Draper<br>Jenner Institute<br>University of Oxford<br>Old Road Campus Research Building<br>Roosevelt Drive<br>Headington Oxford, OX3 7DQ<br>United Kingdom. |
| **Sub-investigators:** | *Ifakara Health Institute, Bagamoyo, Tanzania*<br>1. Ali Mtoro MD MMED<br>2. Caroline Bundi Bsc MSc.<br>3. Florence A Milando MD MPH<br>4. Omary Lweno MD MPH<br>5. Omary Juma MD MPH<br>6. Sarah Mswata BSc.<br>7. Wilmina Kalinga BSc.<br>8. Catherine Mkindi BSc. PhD. |
| **Collaborator** | Dr Stella Rwezaula<br>Haematology and Blood Transfusion Unit.<br>Muhimbili National Hospital<br>Tanzania<br> |
| **Chief Investigator UOXF** | Dr Angela M Minassian<br>Jenner Institute<br>Centre for Clinical Vaccinology & Tropical Medicine<br>University of Oxford<br>Churchill Hospital<br>Old Road<br>Oxford, OX3 7LJ |

| | |
|---|---|
| | United Kingdom |
| **Project Manager (Tanzania)** | *Ifakara Health Institute*<br>Saumu Ahmed MD, PGDip,<br>Tel: +255 784 358 670<br> |
| **Sponsor:** | University of Oxford<br>Named Contact: Dr Rebecca Bryant<br>Research Services<br>University Offices<br>Wellington Square<br>Oxford, OX1 2JD<br>United Kingdom<br>Phone number: +44 (0)1865 282585<br> |
| **Funder:** | The European and Developing Countries Clinical Trials Partnership (EDCTP) |
| **Local Safety Monitor** | Prof Karim Manji, M.D., M.Med., M.P.H.<br>Department of Pediatrics and Child Health<br>Muhimbili University of Health and Allied Sciences<br>Dar es Salaam, Tanzania<br> |
| **Clinical Trial Pharmacist** | Beatus Simon, BSc MPH<br>Ifakara Health Institute<br> |
| **Statistician** | Ummy Abdul, MSc<br>Ifakara Health Institute, Tanzania<br> |
| **Ethics Committees** | Ifakara Health Institute IRB<br>P.O. Box 78373<br>Dar es Salaam, Tanzania<br>Tel: +255 (0) 22 2774714,<br>Fax: + 255 (0) 22 2771714<br><br><br>National Health Research Ethics Sub-Committee (NatHREC)<br>National Institute for Medical Research<br>P.O. Box 9653<br>Dar es Salaam, Tanzania<br>Tel: +255 22 2121400 |

| | |
|---|---|
| | Fax: 255 22 2121360<br><br><br>Oxford Tropical Research Ethics Committee (OxTREC)<br>University of Oxford<br>Research Services<br>University Offices<br>Wellington Square<br>Oxford, OX1 2JD<br>United Kingdom<br>Tel: +44 (0)1865 282585<br> |
| **Regulatory Authority** | Tanzania Medicines and Medicinal Drugs Authority<br>P.O. Box: 77150<br>Dar es Salaam, Tanzania.<br>Tel: +255 22 2450512 / 2450751 / 2452108<br>Fax: +255 22 2450793<br> |
| **Laboratories** | 1. Bagamoyo Research and Training Centre Laboratory, Ifakara Health Institute, Bagamoyo Tanzania.<br>2. Jenner Institute laboratories, Centre for Clinical Vaccinology & Tropical Medicine, University of Oxford, Churchill Hospital, Old Road, Oxford, OX3 7LJ, United Kingdom.<br>3. NIH/NIAID Laboratory of Malaria and Vector Research, Malaria Immunology Section, GIA Reference Centre, 12735 Twinbrook Parkway, Twinbrook III, Room 3W-13, Rockville, MD 20852, USA.<br>4. KEMRI-Wellcome Trust Research Laboratory, CGMRC, PO Box 230-80108, Kilifi, Kenya.<br>5. Department of Biochemistry laboratories, University of Oxford, 3 South Parks Road, OX1 3QU |

#### 5. SYNOPSIS

<table>
<tr><td>Trial Title</td><td>A Phase Ib age de-escalation dose-escalation open label study of the safety and immunogenicity of RH5.1/Matrix-M, administered intramuscularly in healthy adults and infants in Tanzania.</td></tr>
<tr><td>Study reference</td><td>VAC080</td></tr>
<tr><td>Clinical Phase</td><td>Phase Ib</td></tr>
<tr><td>Trial Design</td><td>Age de-escalation dose-escalation open label randomised trial. The study will consist of 6 groups as shown below.
<table>
<tr><th>Group (Age)</th><th># volunteers</th><th>Malaria exposure status</th><th>Month 0</th><th>Month 1</th><th>Month 2</th><th>Month 6</th></tr>
<tr><td>Group 1A (18-45 yrs)</td><td>6</td><td>Low</td><td>10µg RH5.1 /50µg Matrix-M</td><td>10µg RH5.1 /50µg Matrix-M</td><td>10µg RH5.1 /50µg Matrix-M</td><td></td></tr>
<tr><td>Group 1B (18-45 yrs)</td><td>6</td><td>Low</td><td>50µg RH5.1 /50µg Matrix-M</td><td>50µg RH5.1 /50µg Matrix-M</td><td></td><td>10µg RH5.1 /50µg Matrix-M</td></tr>
<tr><td>Group 2A (5-17 months)</td><td>12</td><td>Low</td><td>10µg RH5.1 /50µg Matrix-M</td><td>10µg RH5.1 /50µg Matrix-M</td><td>10µg RH5.1 /50µg Matrix-M</td><td></td></tr>
<tr><td>Group 2B (5-17 months)</td><td>12</td><td>Low</td><td>10µg RH5.1 /50µg Matrix-M</td><td>10µg RH5.1 /50µg Matrix-M</td><td></td><td>10µg RH5.1 /50µg Matrix-M</td></tr>
<tr><td>Group 2C (5-17 months)</td><td>12</td><td>Low</td><td>50µg RH5.1 /50µg Matrix-M</td><td>50µg RH5.1 /50µg Matrix-M</td><td></td><td>10µg RH5.1 /50µg Matrix-M</td></tr>
<tr><td>Group 2D (5-17 months)</td><td>12</td><td>High</td><td>50µg RH5.1 /50µg Matrix-M</td><td>50µg RH5.1 /50µg Matrix-M</td><td></td><td>10µg RH5.1 /50µg Matrix-M</td></tr>
</table></td></tr>
<tr><td>Study population</td><td>Healthy adults (18-45 years) and infants (5-17 months) residing in Bagamoyo district, Tanzania. A total of 60 participants will be enrolled. Participants will be recruited from areas of low malaria transmission in Bagamoyo town and areas of high malaria transmission within Bagamoyo district.</td></tr>
<tr><td>Follow up duration</td><td>All participants will be followed for 2-2.5years after the first vaccination with RH5.1/Matrix-M vaccination. The duration of the entire study will be 2-2.5years per participant from the time of first vaccination.</td></tr>
<tr><td>Planned Trial Period</td><td>2-2.5 years after the start of recruitment</td></tr>
</table>

| | Objectives | Outcome Measures |
|---|---|---|
| Primary | To determine safety and tolerability of RH5.1/Matrix-M given intramuscularly as a 0,1, 2 or 0, 1, 6 month schedule in adults (18-45 years), and infants (5-17 months) residing in a malaria endemic country. | • Solicited symptoms after vaccination.<br>• Unsolicited symptoms after each vaccination.<br>• Serious adverse events during the study period. |
| Secondary | To evaluate the magnitude of humoral and cellular immune responses to RH5 in adults and infants residing in a malaria endemic country.<br>To evaluate the quality of humoral and cellular immune responses to RH5 in adults and infants residing in a malaria endemic country.<br>To evaluate the longevity of humoral and cellular immune responses to RH5 in adults and infants residing in a malaria endemic country. | • Anti-RH5 antibody titres by quantitative ELISA.<br>• Growth inhibition activity of IgG from vaccinees on a panel of *P. falciparum* parasites.<br>• Avidity of anti-RH5 antibodies by ELISA and/or other assays (to be defined).<br>• Cellular immune responses to RH5 by ELISpot assays and/or Flow cytometry and/or other assays (to be defined). |
| Exploratory | To determine the frequency of RH5 specific plasma cells in the bone marrow of adults vaccinated with RH5.1/Matrix-M.<br>To assess impact of malaria exposure status on vaccine responses. | • Frequency of RH5 specific plasma cells from bone marrow aspirates at baseline and 4 weeks after 3rd vaccinations.<br>• Statistical analysis of secondary outcome measures of G2D with other group |
| Investigational Medicinal Product(s) | • RH5.1<br>• Matrix-M | |
| Route of Administration | All vaccines will be given by intramuscular injection to the left deltoid area. | |

#### 6. ABBREVIATIONS

| | |
|---|---|
| ALT | Alanine Aminotransferase |
| ASC | Antibody Secreting Cells |
| BCG | Bacillus Calmette–Guérin |
| BDH | Bagamoyo District Hospital |
| BRTC | Bagamoyo Research and Training Centre |
| BMI | Body Mass Index |
| CBF | Clinical Biomanufacturing Facility |
| CHW | Community Health Worker |
| CMI | Cell Mediated Immunity |
| CRF | Case Report form |
| CRO | Clinical Research Organization |
| CSP | Circumsporozoite Protein |
| CTA | Clinical Trial Agreement |
| DNA | Deoxyribonucleic acid |
| DPT | Diphtheria, Pertussis and Tetanus. |
| EC | Ethic Committee |
| EDC | Electronic Data Capture |
| EPI | Expanded Program of Immunization |
| FVO | Falciparum Vietnam Oak-Knoll |
| GCP | Good Clinical Practice |
| GIA | Growth Inhibition Assay |
| GPI | Glycosylphosphatidylinositol |
| GSK | Glaxosmithkline |
| HBV | Hepatitis B Virus |
| HIV | Human Immunodeficient Virus |
| HRA | Health Research Authority |
| ICF | Informed Consent Form |
| ICS | Informed Consent Sheet |
| IDT | Impfstoffwerke DessauTornau |
| IFN | Interferon |
| IHI | Ifakara Health Institute |
| IPT | Intermittent Presumptive Treatment |
| IRB | Institutional Review Board |
| IRS | Indoor Residual Spraying |
| ISM | Independent Safety Monitor |
| IVD | Immunization and Vaccine Development |
| CTF | Clinical Trial Facility |
| MRC | Medical Research Council |

| | |
|---|---|
| MVA | Modified Vaccinia Ankara |
| NatHREC | National Health Research Ethics Sub-Committee (NatHREC) |
| OPV | Oral Polio Virus |
| OXTREC | Oxford Tropical Research Ethics Committee |
| PBMC | Peripheral blood mononuclear cells |
| PIS | Participant Information Sheet |
| RBC | Red Blood Cells |
| RSV | Respiratory Syncytial Virus |
| SAE | Serious Adverse Event |
| SAR | Serious Adverse Reaction |
| SDV | Source Data Verification |
| DSMB | Data Safety and Monitorig Committee |
| SOP | Standard Operating Procedures |
| SPR | Surface Plasmon Resonance |
| TMDA | Tanzania Medicine and Medical Devices Authority |
| TMF | Trial Master File |
| TSG | Oxford University Hospitals NHS Foundation Trust / University of Oxford Trials Safety Group |
| UOXF | University of Oxford |
| WHO | World Health Organization |

### 7. BACKGROUND AND RATIONALE

#### 7.1. Malaria epidemiology

Malaria still remains a disease of public health significance affecting millions across the globe. In 2015, it was estimated 212 million new cases of clinical malaria were diagnosed worldwide resulting in more than 430,000 malaria deaths [1]. This is likely to be an underestimation since the estimates are highly influenced by the surveillance methods used [2]. The highest malaria burden is in children below the age of five years among whom over 303,000 and 292,000 deaths were reported worldwide and in Africa respectively in 2015. In addition to the immediate morbidity and mortality risk, malaria may inflict long-term effects with negative consequences on health and quality of life. For example, children who suffer from severe malaria are at increased risk of epilepsy [3,4], chronic neurological and cognitive impairment [5]. Furthermore, malaria causes significant economic hardships and disproportionately affects the poor [6]. In endemic countries, it is estimated that more than 1% loss in gross domestic product is attributable to malaria [7,8].

According to the Tanzania HIV/AIDS and Malaria Indicator Surveys (THMIS) carried out in 2008 and 2012, malaria prevalence in children aged 6-59 months has declined by 50%, from 18.1% to 9.5%. However, there is significant variations across regions with prevalence varying from less than 1% to greater than 33%. Therefore, malaria still remains a disease of public health significance. The goal of National Malaria Strategic plan of 2014-2020 was to reduce the average country malaria prevalence from 10% in 2012 to 5% in 2016 and further down to less than 1% in 2020 [9].

Malaria control interventions such as insecticide treated bed-nets (ITNs), treatment with artemisinin based combination therapies (ACTs), intermittent presumptive treatment (IPT) and indoor residual spraying (IRS) have made a significant impact on the burden of malaria. There have been encouraging reports on a decline of malaria from several parts of Africa [10-13] but this has not been consistent everywhere with some areas reporting sustained or even an increase in the burden of malaria [14,15].

The progress made in controlling malaria is however being threatened by development of resistance in the parasite and in the vector. The above interventions also do not appear sufficient to eliminate malaria in high transmission settings in Africa [16].

#### 7.2. Malaria vaccines

Immunisation against communicable diseases is one of the most cost-effective public health interventions [17,18]. The global health success of eradicating smallpox was the result of a widespread deployment of smallpox vaccine [19]. Furthermore, significant inroads have been made in the fight against other major childhood infectious diseases such as polio, *Haemophilus influenzae* and measles through immunisation [20,21]. The success of vaccines is based not only on their ability to offer protection to the individual recipients, but also to the community by reducing the transmission through herd immunity [22,23]. Malaria vaccines are regarded as important component in the fight against malaria in Tanzania by the National Malaria Strategic Plan of 2014-2020 [9].

There have been many strategies utilized in the development of an effective malaria vaccine including development of peptides [24,25], recombinant proteins and fusion proteins [26-29] targeting specific stages of the *Plasmodium falciparum* (*Pf*) life cycle, non-replicating whole sporozoites [30], DNA vaccines [31] and vectored and prime-boost vaccines [32].

The most clinically advanced malaria vaccine candidate, RTS,S/AS01, has shown modest efficacy in children and children in Phase II and III field trials [33,34]. The efficacy wanes over time and there is potential for rebound in areas with high malaria transmission [35], providing strong rationale for the future inclusion of an effective blood-stage component to prevent clinical disease as pre-erythrocytic anti-infection immunity wanes.

Thrombospondin-related adhesion protein (TRAP) fused to a multi-epitope (ME) is another pre-erythrocytic vaccine candidate that has shown promising results and is currently being evaluated in the field [36,37]. The antigen is delivered using a viral vector prime-boost strategy. Early studies using fowlpox strain 9 or plasmid DNA as priming vector followed by MVA were unsuccessful in providing adequate protection in malaria-exposed individuals [38]. Chimpanzee adenovirus serotype 63 (ChAd63) as a priming vector has provided better immunogenicity and significant protection, as has been demonstrated in both malaria naïve and malaria exposed adults [39-41]. The trials to investigate the efficacy of ME-TRAP in children and infants are ongoing.

Whole sporozoite-based approaches provide short term homologous protection in malaria-naïve adults up to 14 months after the last dose [30,42]. The vaccine induces high levels of tissue resident $CD8^+$ T cell responses and moderate levels of antibodies to Pf Circumsporozoite protein (CSP), both of which correlate with protection. In malaria exposed individuals who receive comparable doses to malaria naïve adults, efficacy against malaria infection is moderate both by experimental and natural challenge [43]. Studies are still ongoing to further optimize the dose and schedule including evaluating the potential of non-attenuated whole sporozoite given under anti-malarial chemoprophylaxis in malaria exposed individuals. Transmission-blocking vaccines would be ideal for elimination, however, most of these vaccine candidates are still in early stages of development [44,45].

#### 7.3. Blood-stage malaria vaccines

Blood-stage malaria vaccines are essential for prevention of morbidity and mortality from malaria. The importance of blood-stage naturally acquired immune responses was demonstrated by experiments using passive transfer of immunoglobulins [46,47]. However the naturally acquired immunity develops slowly over time, does not completely prevent infection and wanes if exposure is removed [48]. In 2002 it was shown that blood-stage immunity can be induced experimentally through immunization with ultra-low doses of infected erythrocytes [49], although the effect of antimalarial used to clear parasitaemia in this study could not be completely ruled out as confounding factor.

Most of the research on blood-stage vaccines has focused on the antigens, which are critical in the host-parasite interactions and expressed on the merozoites. Although the safety and immunogenicity of many blood-stage candidate vaccines have been promising, they have shown no clinical protection [29,50] or only strain-specific protection [28] in the field. Substantial levels of polymorphism among the most widely studied blood-stage antigens and redundant erythrocyte invasion pathways present a major challenge for the development of blood-stage vaccines [51]. In addition rapid erythrocyte invasion means extremely high concentrations of functional antibody are required to neutralize the parasite to prevent erythrocyte invasion [52].

Thus while the development of blood-stage malaria vaccines is faced with challenges, they remain a critical element of the malaria vaccine portfolio. With the decline in malaria transmission and subsequent waning of naturally acquired immunity, populations living in a malaria endemic country will become increasingly at risk of a malaria epidemic once the interventions are interrupted [53], making a blood-stage malaria vaccine an essential component of any integrated malaria control programme. Furthermore, blood-stage malaria vaccines need to be part of the multi-stage vaccine that would protect against rebound malaria when the immunity to the pre-erythrocytic component of the vaccine wanes over time.

#### 7.4. *Plasmodium falciparum* reticulocyte-binding protein homolog 5

*Pf* reticulocyte-binding protein homolog 5 (PfRH5) is a new generation and highly promising blood-stage malaria vaccine antigen [54]. The PfRH5 is part of the family of a protein complex consisting of PfRH1, PfRH2a, PfRH2b, PfRH3 and PfRH4. PfRH proteins are located in the apical organelles of the merozoite and are released onto the surface during invasion of erythrocytes. In contrast to other protein members, PfRH5 is a much shorter protein (~60kDa for PfRH5 vs 200-375kDa for other PfRH proteins) and does not have a transmembrane and cytosolic region at the C-terminus [55].

PfRH5 forms a complex with another two antigens, namely PfRH5-interacting protein (PfRipr) [56] and the cysteine-rich protective antigen (PfCyRPA) [57] to facilitate interaction with the erythrocyte. It has recently been shown that the N-terminal region of RH5 (RH5Nt) binds glycosylphosphatidylinositol (GPI)-anchored merozoite protein P113, providing a platform for RH5 to attach to the merozoite surface [58]. It is hypothesized that CyRPA recruits Ripr to the complex resulting in the release of the RH5-CyRPA-Ripr complex. The RH5-CyRPA-Ripr complex is thought to play a role in the formation of a pore between the merozoite and the RBC that allows $Ca^{2+}$ ingress into the host cell prior to establishment of the tight junction.

PfRH5 binds basigin on the surface of erythrocytes forming a critical non-redundant interaction during erythrocyte invasion [59]. Immunization with full-length PfRH5 immunogens induces functional antibodies to PfRH5, unlike fragments of antigens produced from *E. coli* [55,60]. Because of the technical challenge of generating full-length PfRH5 protein (RH5_FL), the earliest promising results were achieved using viral vectored immunization [61], whereby antigen is expressed *in situ* from virally infected muscle cells [62]. Recently, production of full-length PfRH5 protein has been successful from numerous heterologous expression systems including mammalian HEK293 cells [63], *E. coli* [64,65], baculovirus-infected insect cells [66,67], wheatgerm [68],and *Drosophila* S2 stable cell lines [69].

PfRH5 has several advantages compared to other investigational blood-stage malaria vaccine candidates, namely PfMSP1 and PfAMA1. PfRH5 is highly conserved across parasite lines, thus leading to strain-transcending neutralizing antibodies post-vaccination [61,70]. Furthermore, the high degree of PfRH5 sequence conservation is associated with functional constraints linked to basigin binding. Substitution of minimal amino acids in PfRH5 results in loss of basigin binding, suggesting the antigen may not easily escape vaccine-induced immune pressure [71-73].

Antibodies to PfRH5 can also block invasion of erythrocytes with high potency, quantitatively requiring less antibody levels to achieve maximal blockage compared to other antigens [74]. In addition, antibodies to PfRH5 can cross-inhibit all field isolates and laboratory strains of *Pf* tested to date [59,61,70]. Naturally-acquired antibodies to PfRH5 are several fold lower compared to other blood-stage antigens such as PfMSP1 and PfAMA1 indicating that PfRH5 is not a dominant target of naturally acquired immunity [66,75]. However, antibodies to PfRH5 have been shown to inhibit parasite growth *in vitro* and predict protection from malaria in the field [66,76].

#### 7.5. RH5 vaccine clinical development

Given their previous success as antigen delivery platforms in vaccines against HIV, Ebola and Tuberculosis [77], viral vectors were used with the RH5 antigen during early RH5 candidate vaccine human clinical trials. This approach uses modified live viruses as vectors that carry antigens or antigen-encoding genes for the antigen of interest leading to the *in vivo* expression of these antigens when subjects are immunized. Their advantage over traditional vaccine delivery platforms was ability to stimulate a broad range of immune responses including both humoral and cell mediated immunity [62,78]. In the case of RH5, a recombinant replication-deficient adenovirus of simian serotype was used to prime the immune response, followed by a booster vaccination 8 weeks later with an attenuated poxvirus recombinant for the same antigen [62].

Viral-vectors were preferred platforms largely due to difficulty in producing recombinant antigens using heterologous expression platforms [78-80]. Being one of such antigens, RH5_FL was difficult to produce using conventional recombinant technology until a viral vector platform was utilized leading to promising data that led to a clinical trial in humans [61]. Despite encouraging results of RH5 administered in viral vectors, the serum antibody levels only reached peak median levels of ~9 µg/mL and 150 µg/mL PfRH5-specific IgG in malaria naïve adults[81] and malaria exposed infants in Tanzania (*Olotu A et al., under preparation*) respectively, suggesting substantial room for improvement in terms of quantitative vaccine immunogenicity. Indeed, previous malaria vaccine candidates delivered as recombinant antigen formulated in strong adjuvant (such as Alhydrogel + CPG 7909 or GlaxoSmithKline's (GSK) adjuvant system $AS01_B$) have achieved peak responses of ≥100 µg/mL antigen-specific serum IgG in humans[82-84].

Recent advances in technology have led to a development of protein expression and purification process that; i) allowed for production of full-length RH5 protein, and (ii) was scalable and compliant with current good manufacturing practice (cGMP). This expression uses cGMP-compliant platform called ExpreS2, based on a Drosophila melanogaster Schneider 2 (S2) stable cell line system [85]. Following expression from stable S2 cell lines, secreted protein is purified using affinity purification system that uses C-terminal tag known as "C-tag" [79]

##### 7.6. Experience with Matrix-M adjuvant used with other vaccines

The Matrix-M adjuvant is manufactured by Novavax AB (Uppsala, Sweden), a subsidiary of Novavax, Inc. The adjuvant contains purified saponin components derived from an extract of the bark of the Quillaja saponaria tree; a phospholipid, egg-derived phosphatidylcholine (PC); and semi-synthetic cholesterol of non-animal origin. The detailed information is contained in the safety Matrix M adjuvant safety data supplement that has been submitted with the protocol.

Matrix-M adjuvant, in 1 of 2 formulations (named Matrix-M1 or Matrix-M2), has been administered to 2820 individuals, of which over 2250 have received Matrix-M1, in a total of 20 clinical trials in the US, Europe, and Australia. Collectively, the clinical data with Matrix-M at doses up to 75 µg, formulated with inactivated influenza viruses, a virosomal avian influenza vaccine, recombinant HSV proteins, inactivated rabies virus, EBOV protein antigen, an influenza VLP, RSV F nanoparticle, or influenza nanoparticle, all indicate that vaccines containing the adjuvant have reversible acute reactogenicity but are generally well-tolerated and demonstrate an acceptably safety profile. Matrix-M-adjuvanted vaccine formulations have also demonstrated a clear immunogenicity benefit, with documented evidence of antigen dose sparing and in some cases broadening of antibody specificities and/or induction of otherwise unapparent T-cell responses.

Injection site pain is the most commonly reported local complaint, occurring more frequently in Matrix-M-adjuvanted vaccinees (50% of subjects) than in unadjuvanted vaccinees (26%) or placebo subjects (8%), with a very slight increase incidence with higher dose of the adjuvant (< 50 vs 50 µg of Matrix-M1 adjuvant). Reports of injection site redness, swelling, and bruising all occurred at lower incidences relative to pain, but were more frequent among adjuvanted vaccinees (7 to 13% of subjects), compared to unadjuvanted vaccinees (3 to 5% of subjects) or placebo subjects (1 to 2% of subjects). Among the systemic complaints, headache, muscle pain, fatigue, chills, joint pain, wheezing, and eyelid swelling all occurred at higher rates among adjuvanted vaccinees, when compared to placebo vaccinees at an excess of ≥ 1% of subjects. No apparent increase in incidence of the AEs was noted with higher dose of the adjuvant. While almost all solicited local and systemic complains across studies were mild to moderate in nature and resolved within the 7-day solicitation period, about 4% of all adjuvanted vaccinees experienced solicited AEs classified as severe, compared to about 2% of placebo and 3% of unadjuvanted vaccinees. The most common severe solicited AE among adjuvanted vaccinees (ie, excess of ≥ 1.0% of subjects when compared with placebo or unadjuvanted vaccinees) was muscle pain.

#### 7.7. Experience with Matrix-M used with Malaria vaccine antigens

Matrix-M has been administered with malaria antigens in healthy malaria naïve adults in the UK and malaria-exposed adults in Africa, at doses of 25µg and 50µg.

These trials include assessment of two pre-erythrocytic malaria candidate vaccines ChAd63/MVA ME-TRAP and the virus-like particle (VLP) R21, adjuvanted with Matrix-M1, within four Phase I and Phase I/II trials conducted in Oxford and Burkina Faso (ClinicalTrials.gov identifiers: NCT02572388, NCT02925403, NCT01669512, NCT02905019). Available safety data demonstrates that the Matrix-M adjuvant is well-tolerated and there have been no serious unexpected adverse reactions or adverse reactions of special interest reported to date. Furthermore the safety profile of Matrix-M is significantly more favorable compared with AS01 adjuvant which has been widely used with candidate malaria vaccines RTS,S.

The most reported local solicited AE was pain at injection site, and it was predominantly mild in severity. Other local solicited AE were redness, swelling, and warmth at the site of injection. Arthralgia, malaise, headache, nausea, fatigue and myalgia were the commonest systemic solicited AEs and they were mostly of mild and moderate severity.

There are currently ongoing studies in Kenya and Burkina Faso comparing 25µg and 50µg Matrix-M in adults, children and infants.

Data from the Kenyan study (enrolment began in April 2019, all vaccinations completed by December 2019) is still undergoing a final analysis for the infant group (group 3, aged 5-11 months, n=51) by dose of R21 and Matrix-M, but the majority of AEs were mild in nature, with most of the infant group experiencing no local adverse events, regardless of the dose of Matrix-M they received (**Figure 1**).

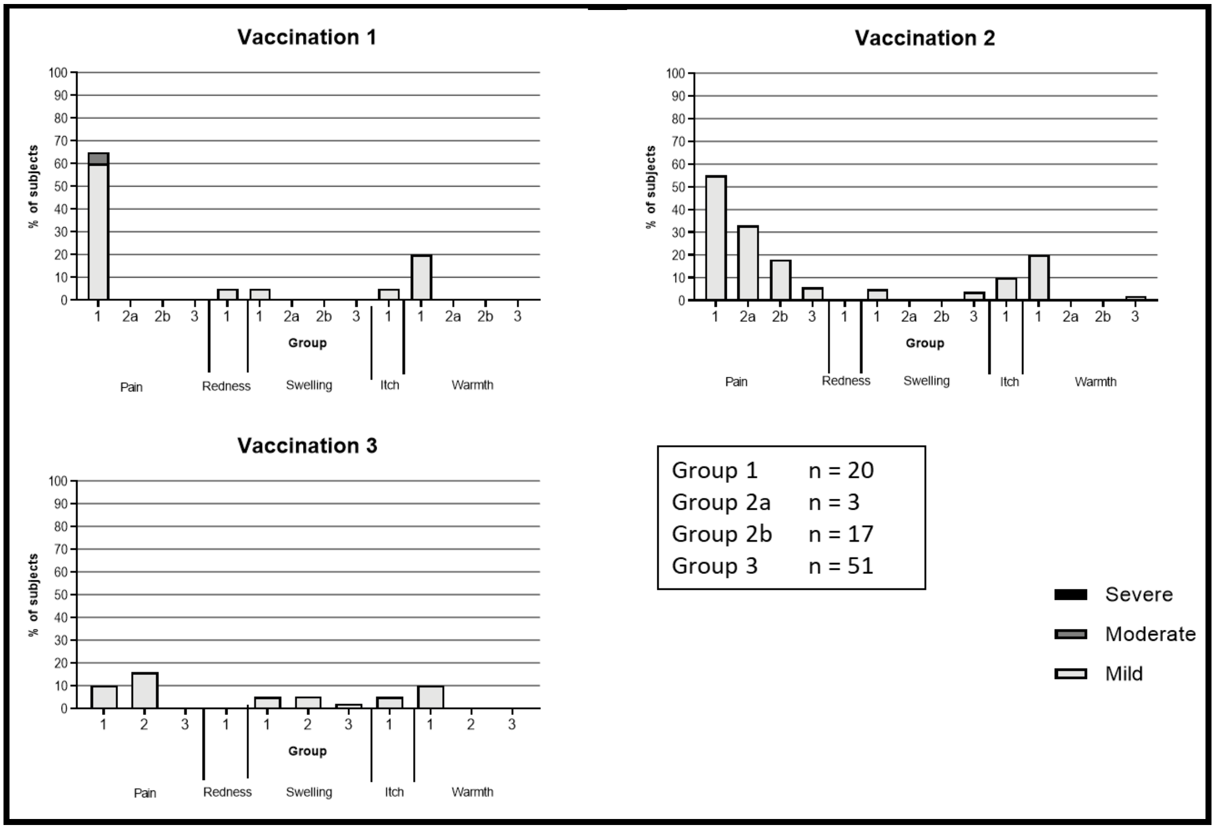

*Figure 1: Local adverse event profile of R21/Matrix-M – 3 doses given 4 weeks apart in different dose groups: Group 1 – Adults, 10µg R21/50µg MM; Group 2a – Children, 5µg R21/25µg MM; Group 2b – Children, 10µg R21/50µg MM; Group 3 – Infants (received either 5µg R21/25µg MM (n=18), 10µg R21/50µg MM (n=18) or 5µg R21/50µg MM (n=15). Combined safety analysis shown for Group 3.*

The most frequent noted systemic adverse event after the second or third vaccination was fever. After the second vaccination 7/21 children (Group 2) had fever and 16/51 (31%) infants (Group 3) had fever (1 severe). After the third vaccination 8/21 in group 2 had fever (1 severe) and 20/51 in group 3 (1 severe). Fevers were seen in all groups but severe fevers were in the higher adjuvant group (50 µg MM). Severe fevers were defined as >39.$^{0}$C **(Figure 2).**

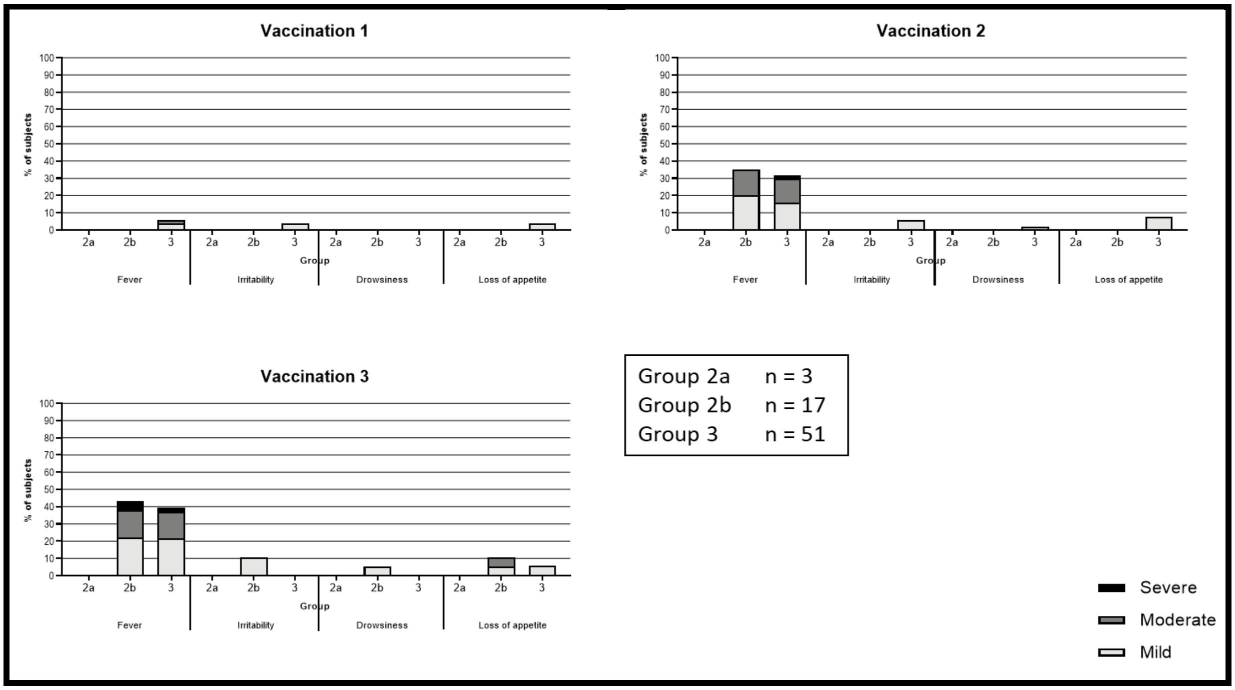

*Figure 2: Fig 2: Systemic adverse event profile of R21/Matrix-M – 3 doses given 4 weeks apart at different doses: Group 2a – Children, 5µg R21/25µg MM; Group 2b – Children, 10µg R21/50µg MM; Group 3 – Infants (received either 5µg R21/25µg MM (n=18), 10µg R21/50µg MM (n=18) or 5µg R21/50µg MM (n=15). Combined safety analysis shown for Group 3.*

In Burkina Faso, infants were vaccinated with either 5µg R21 with 25µg of Matrix-M (N=150) or 5µg R21with 50 µg of Matrix-M (N=150). The control group received rabies vaccine (N=150). There overall reactogenicity data in all groups was very satisfactory with only few mild adverse events reported (**Figure 3**). For systemic solicited adverse events, 35 and 16 participants out of 450 had Grade 1 and Grade 2 fever respectively after first vaccination, 39 and 27 participants out of 445 had Grade 1 and Grade 2 fever respectively after the second vaccination, and 33 and 23 participants out 442 had Grade 2 and Grade 3 fever respectively after the third vaccination **(Figure 4**).

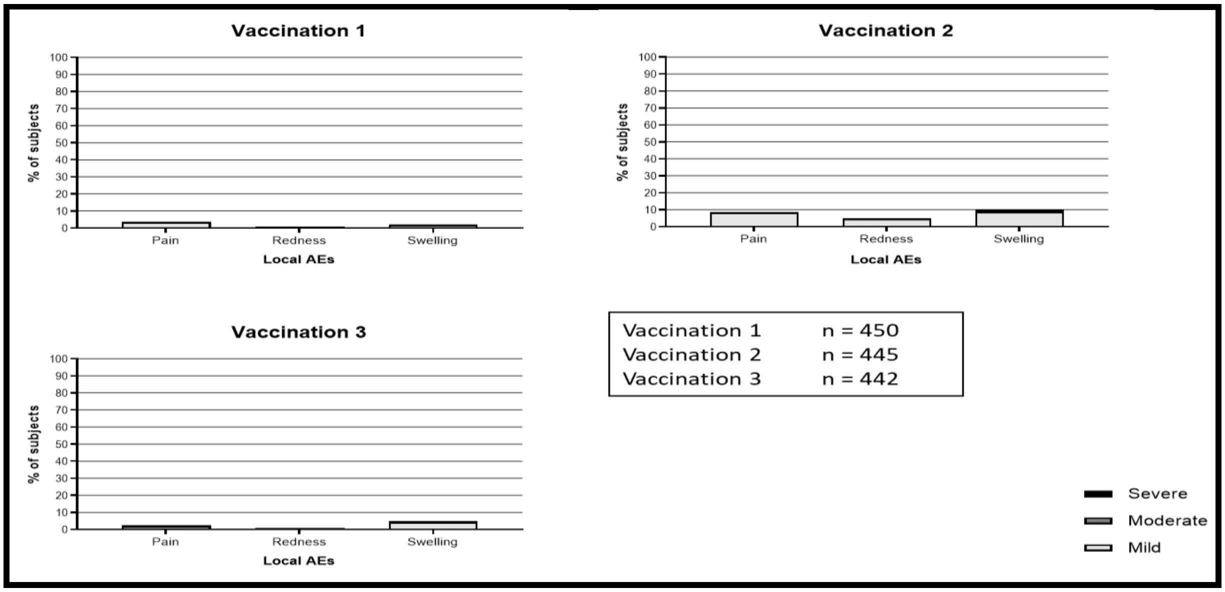

*Figure 3: Local adverse event profile of R21/Matrix-M – 3 doses given 4 weeks apart in different dose groups: Group 1 (N=150) Infants, 5µg R21/25µg MM; Group 2 (N=150) Infants, 5µg R21/50µg MM or Group 3 (N=150) infants Rabies vaccine. Combined safety analysis shown for all Groups.*

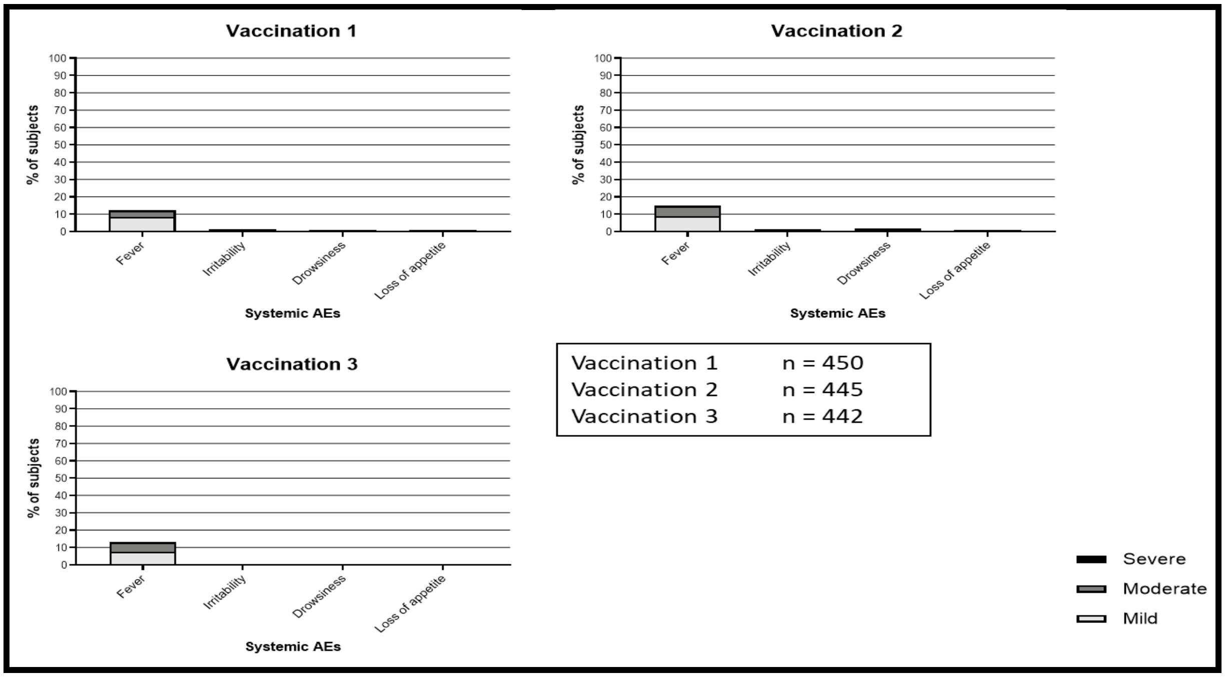

*Figure 4: Systemic solicited adverse event profile of R21/Matrix-M – 3 doses given 4 weeks apart in different dose groups: Group 1 (N=150) Infants, 5µg R21/25µg MM; Group 2 (N=150) Infants, 5µg R21/50µg MM or Group 3 (N=150) infants Rabies vaccine. Combined safety analysis shown for all Groups.*

**Figure 5** shows the antibodies to the NANP repeat of the CS protein, at baseline and at Day 84 (1 month post the 3rd vaccine). All samples were positive by Day 84 (all children were negative at baseline) and overall antibody levels exceeded those of the RTS,S/AS01 vaccine by about 3-fold. In particular, these data show the subgroup analysis for infants – those infants receiving the higher 50µg dose of MM (G3e) compared to those receiving the lower 25µg dose (G3a/b) (but the same 5µg dose of R21), showed higher mean antibody titres of 15122 ELISA units compared to 10466, respectively. This study therefore shows the higher dose of Matrix M to be more immunogenic in Kenyan infants.

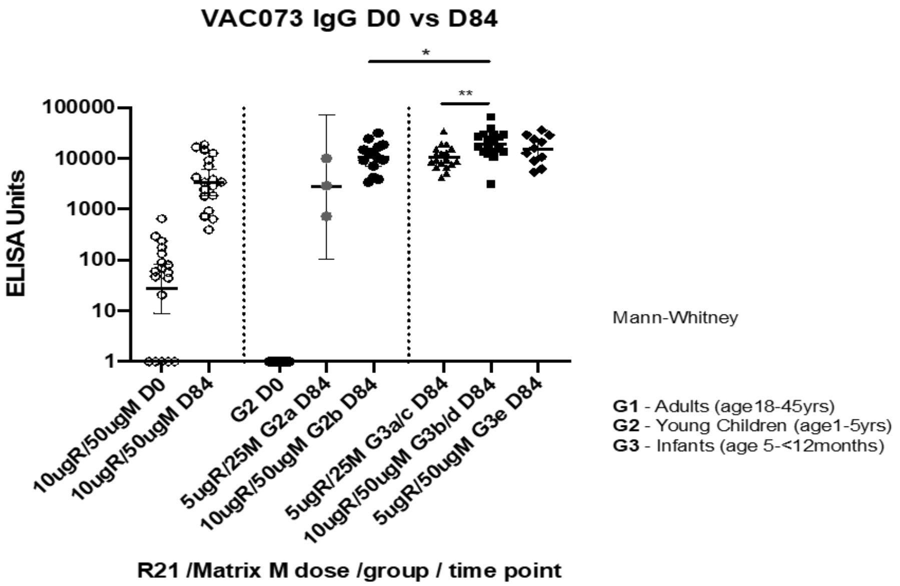

| | Adults | | 5ug R21/25ugMM | | 10ug R21/50ugMM | | 5ug R21/25ugMM | 10ug R21/50ugMM | 5ug R21/50ugMM |
|---|---|---|---|---|---|---|---|---|---|
| | D0 | D84 | D0 | D84 | D0 | D84 | D84 | D84 | D84 |
| Geo mean titre | 27 | **3348** | 1.0 | **2746** | 1.0 | **10446** | **10465** | **18939** | **15122** |
| Lower 95% CI of GM | 8.9 | 1870 | 1.0 | 106 | 1.0 | 7075 | 8102 | 13728 | 9786 |
| Upper 95% CI of GM | 84.0 | 5993 | 1.0 | 71458 | 1.0 | 15424 | 13518 | 26129 | 23369 |

*Figure 5: Antibodies to the NANP repeat of the CS protein, at baseline and at Day 84 (1 month post the 3rd vaccine). Group 1 – Adults, 10µg R21/50µg MM; Group 2a – Children, 5µg R21/25µg MM; Group 2b – Children, 10µg R21/50µg MM; Group 3 – Infants (received either 5µg R21/25µg MM (n=18, Group 3a/c), 10µg R21/50µg MM (n=18, Group 3b/d) or 5µg R21/50µg MM (n=15, Group 3e).*

#### 7.8. RH5 candidate malaria vaccine; key safety and immunogenicity data

An RH5 candidate malaria vaccine is being developed for the routine immunization of infants living in malaria-endemic areas as part of the Expanded Program of Immunization (EPI). In addition, it is being developed as an essential component of a multi-stage malaria vaccine aimed to completely block malaria transmission. The clinical development plan for RH5 involves investigation of different approaches to deploy the antigen, including protein-in-adjuvant, viral vectors and novel vaccine adjuvants. To date, the protein-in-adjuvant RH5 candidate malaria vaccine has been administered to malaria-naïve adult subjects as well as non-human primates.

*Pre-clinical data of RH5*

A pre-clinical study using viral vectors or RH5 protein produced from mammalian HEK293 cells showed that RH5-based vaccine vaccines can protect *Aotus* monkeys against a virulent vaccine-heterologous *Pf* challenge [79]. Protection was strongly correlated with anti-RH5 serum IgG antibody concentration and *in vitro* functional growth inhibition activity (GIA) using purified IgG [79], confirming the utility of this assay to predict *in vivo* protection and for future vaccine candidate down-selection. RH5 was administered with viral vector and as protein-in-adjuvant.  There were no safety concerns from animal studies.

*Clinical experience of RH5.1 protein in adjuvant*

Full length RH5.1 protein expressed using this new system was recently evaluated in 65 UK healthy adults in combination with the GSKBiological's manufactured AS01 adjuvant to assess safety, immunogenicity and efficacy (VAC063 trial, NCT02927145).  Three doses of RH5.1 (2, 10 and 50 µg) were tested, and both a normal 0,1,2-month dose schedule and fractional delayed dose of 0,1,6-month schedule were compared (**Figure 6).** The 10 µg (0, 1, 2-month) dose was then taken forward to a Phase IIa efficacy study (new group 5), where 14 vaccinees received a primary homologous blood-stage challenge in parallel with 15 control volunteers (new group 6). About 4 months later, 9 of these 14 group 5 vaccinees (group 7) went on to receive a fourth and final boost with RH5.1/AS01 $_{B.}$ This was followed 2 weeks later by a second homologous blood-stage challenge, in parallel with 8 of the previous control volunteers (group 8) and 6 new malaria-naïve controls (group 9). This was to investigate the durability of any vaccine effect (in parallel with any protective effect of a prior homologous challenge).

| | Group | Group size | Day 0 | Day 28 | Day 56 | Day 182 | | |
|---|---|---|---|---|---|---|---|---|
| *Phase Ia* | 1 | 6 – 12 | 2µg RH5.1/ 0.5mL AS01 | 2µg RH5.1/ 0.5mL AS01 | 2µg RH5.1/ 0.5mL AS01 | | | |
| | 2 | 6 – 12 | 10µg RH5.1/ 0.5mL AS01 | 10µg RH5.1/ 0.5mL AS01 | 10µg RH5.1/ 0.5mL AS01 | | | |
| | 3 | 12 | 50µg RH5.1/ 0.5mL AS01 | 50µg RH5.1/ 0.5mL AS01 | | 10µg RH5.1/ 0.5mL AS01 | | |
| | 4 | 12 | 50µg RH5.1/ 0.5mL AS01 | 50µg RH5.1/ 0.5mL AS01 | 50µg RH5.1/ 0.5mL AS01 | | | |
| | **Group** | **Group size** | **Day 0** | **Day 28** | **Day 56** | **2 weeks post-third vaccination** | **~4 months post third vaccination** | **1-2 weeks post-final vaccination** |
| *Phase IIa* | 5 | 15 (+2) | 10µg RH5.1/ 0.5mL AS01 | 10µg RH5.1/ 0.5mL AS01 | 10µg RH5.1/ 0.5mL AS01 | CHMI | | |
| | 6 (controls) | 15 (+2) | | | | CHMI | | |
| | 7 (Subset of Group 5 vaccinees) | 5-15 | 10µg RH5.1/ 0.5mL AS01 | 10µg RH5.1/ 0.5mL AS01 | 10µg RH5.1/ 0.5mL AS01 | CHMI | 10µg RH5.1/ 0.5mL AS01 | CHMI |
| | 8 (Subset of Group 6 controls) | 5-15 | | | | CHMI | | CHMI |
| | 9 (New controls) | 5-6 (+2) | | | | | | CHMI |

*Figure 6: VAC063 vaccination groups receiving RH5.1 with AS01 adjuvant*

The vaccine was well tolerated, with solicited adverse events primarily mild to moderate in severity, although occasional severe adverse events occurred. Of a total of 202 vaccine doses administered in the study, 5% of doses produced severe redness at the vaccination site, 2% produced severe swelling, and 1% of doses produced severe myalgia or malaise. All severe local adverse events had resolved by 48 hours following vaccination (except for one episode of redness which lasted for 96 hours before fully settling). Slightly more frequent reactogenicity (pain/redness/swelling/warmth) was reported in the high dose group after the second and third vaccinations compared to after the first vaccination. Systemic AEs were infrequent at all doses after the first vaccination, except for fatigue which was common, reported in 44-83% of volunteers. This and the majority of other systemic AEs after the first vaccination were mild. While the majority of systemic AEs were still mild after subsequent vaccinations, a higher proportion were reported as moderate after the second and third vaccinations, regardless of dose. All systemic AEs resolved within 24 hours. Both local and systemic solicited AEs were likely due to the adjuvant component of the vaccine (AS01B is known for its reactogenicity). Unsolicited adverse events were predominantly mild in nature in all dose groups.

There was one unexpected SAE classified as possibly related to vaccination with RH5.1/$AS01_B$ and so reported as a SUSAR. This was development of scalp psoriasis in a volunteer 4 weeks after they had received the second dose of the vaccine. This initially was not felt to fulfil the criteria for an SAE, but then reassessed as psoriasis is an autoimmune condition (and according to the study-specific protocol, all new or suspected autoimmune diseases should be defined as AEs of special interest and should therefore be reported as SAEs). Given there were no expected SARs to the vaccine and the timing of onset made a temporal relationship plausible, it was agreed that this SAE should be upgraded to a SUSAR.

There were no other vaccine-related SAEs reported during the study period.

There was no difference between 2µg, 10µg or 50µg RH5.1 dose with regard to peak RH5.1_FL antibody levels, but overall RH5.1/AS01B induced 10-fold higher antibody responses than what was induced following vaccination with ChAd63/MVA expressing RH5. Delayed fractional dosing (0,1,6 month) induced higher peak antibody responses at 4 weeks post final vaccination compared to 0,1, 2-month vaccination regimens (5). It was shown that delayed fractional dosing significantly increased the longevity and avidity of the anti-RH5_FL antibodies compared to standard 0,1,2-month regimen**(Figure 9**). For the first time vaccination withRH5.1/AS01 (10 µg at the standard 0,1,2-month regimen led to reduced parasite multiplication rate (PMR, median 17% reduction) following challenge with heterologous infected RBC compared to controls (**Figure 10**). Following a fourth vaccine dose and secondary challenge this effect was increased (median 33% reduction in PMR) (Minassian *et al.,* submitted).

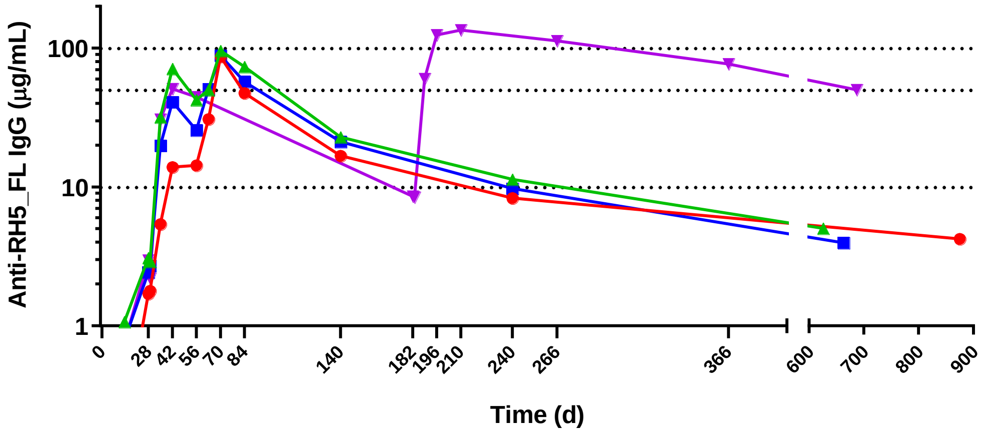

*Figure 7: Median anti-RH5_FL serum total IgG responses following immunization with RH51 in AS01B shown for Groups 1-4 over time. Group 3 (in purple) = delayed fractional dose group, showing higher peak anti-RH5 responses and superior longevity over the other groups.*

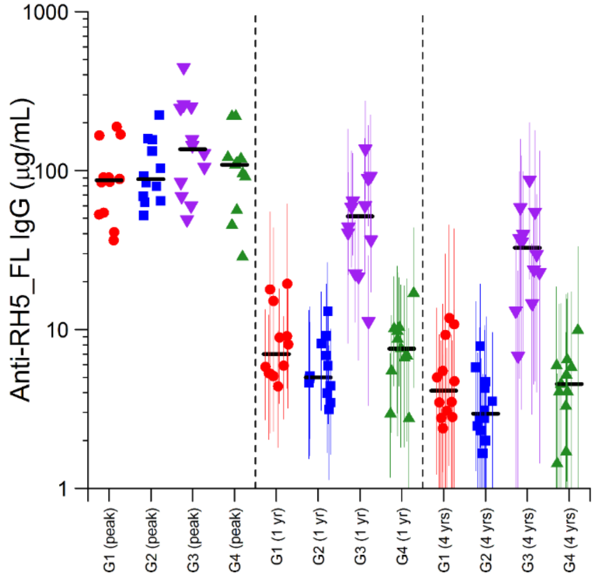

*Figure 8: Anti-RH5_FL antibody levels at peak, and one and four years following the third vaccine dose: peak antibody levels are based on the maximum measured values (d70 or d84 for Groups 1, 2 and 4; and on d196 or d210 for Group 3); antibody levels at one and four years are based on model estimates and are presented with 95% credible intervals.*

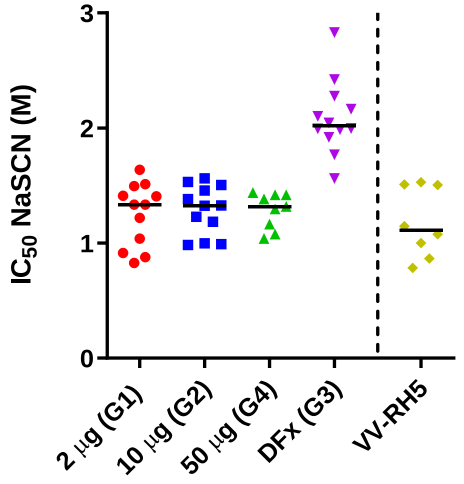

*Figure 9: Peak Anti-RH5.1_FL avidity for different dosing regimens. DFx = delayed fractional dose group. Avidity of serum total IgG responses at 14 days after three immunizations (d70 or d196) was assessed by NaSCN-displacement RH5_FL ELISA and is reported as the molar (M) concentration of NaSCN required to reduce the starting OD in the ELISA by 50% ($IC_{50}$). Historical data for the viral-vectored (VV) RH5 vaccine are shown for comparison.*

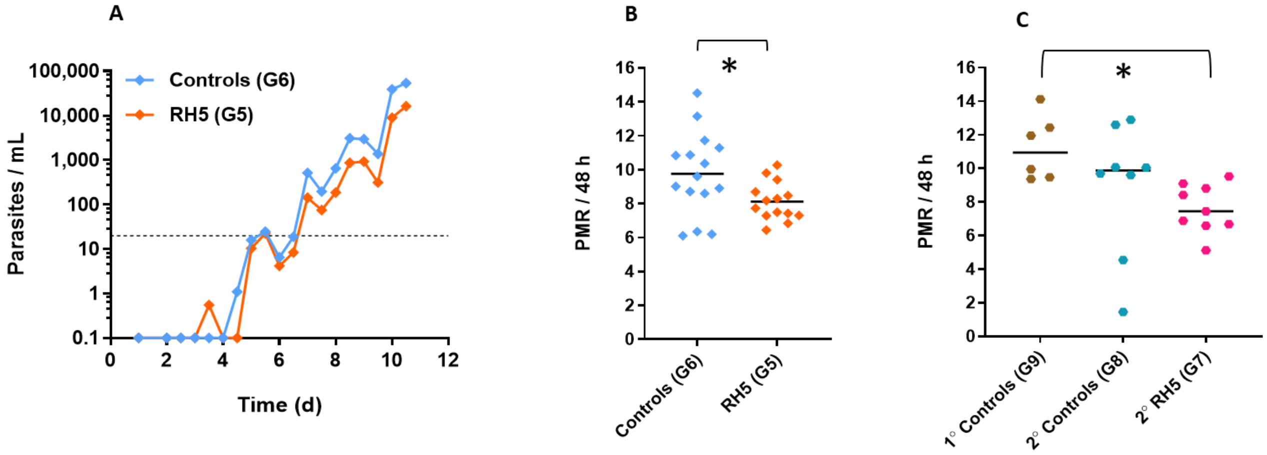

*Figure 10: (A): qPCR data for the VAC063A Phase IIa study; Group 5 vaccinees (n=14) and Group 6 (n=15). Median parasitemia is shown over time for each group. The lower limit of quantification is indicated by the dotted line at 20 p/mL. Time=days (d) post blood-stage CHMI. (B) Primary efficacy endpoint analysis of PMR, showing each individual, plus the mean. Both datasets are normally distributed (D'Agostino-Pearson test); * P=0.031 using two-tailed t-test with Welch's correction for non-equal variances (F-test; P=0.008). (C): Secondary efficacy endpoint analysis of PMR, showing each individual, plus the median for the VAC063B Phase IIa study. Group 7 (n=9), Group 8 (n=8) and Group 9 (n=6). * P=0.022; Kruskal-Wallis test with Dunn's multiple comparison test.*

#### 7.9. Rationale

Malaria vaccines are important component in the fight against malaria in Tanzania[9] and research that address this need is essential for our country. As described in section 7.8, RH5 has undergone several evaluations to test its safety, immunogenicity and efficacy in malaria-naïve adults. ChAd63/MVA expressing RH5 have been found to be safe and immunogenic in malaria naïve healthy UK adults. In an endemic setting ChAd63/MVA RH5 was evaluated in adults, children and infants in Tanzania in a study that was completed in March 2019. The vaccine was shown to be safe, and immunogenic, inducing a 5-fold increase in antibody levels in young children and infants compared to malaria-naïve adults in UK.

A trial testing RH5.1 administered in AS01B demonstrated a 10-fold improvement in immunogenicity compared to viral vectored RH5 in UK adults, and demonstrated an impact on parasite growth following homologous challenges in malaria-naïve healthy UK adults **(Figure 10)**. The duration of antibody responses was also improved in the delayed fractional dose cohort, compared to what was observed with viral vectors. The delayed fractional dose regime of RH5.1/AS01 induced a higher magnitude, increased avidity and improved longevity of RH5-specifc antibodies compared to the normal 0,1, 2-month dose schedule **(Figure 7, Figure 8, Figure 9).** Similarly, vaccination of malaria exposed infants with RH5.1 protein in adjuvant could result into 5-fold higher RH5.1_FL antibody levels compared to levels observed in malaria naïve adults. It is therefore important to investigate whether RH5.1 protein vaccine when given with potent adjuvant would lead to improved immunogenicity in target population and if delayed fractional dosing could improve the antibody level, quality and duration in target population.

Recently Matrix-M adjuvant, was used with a pre-erythrocytic malaria vaccine R21. When compared with AS01, Matrix-M with R21 had better safety outcomes and similar immunogenicity outcomes. As a result, Matrix-M was selected for the next RH5.1 trial in Tanzania.

The results from this trial will inform decisions to proceed with field studies to evaluate efficacy, or to provide more insight into the need to further improve the RH5 vaccine immunogen to focus the response on key neutralising epitopes recognised by the human B cell repertoire. This trial will address five important questions: a) Does RH5.1 protein vaccine see improved immunogenicity in target population? b) Does RH5.1 protein vaccine delayed fractional dosing improve immunogenicity in target population? d) If immunogenicity is improved in target population, how does this impact parasite growth *in vitro*? and; e) Is it the delay or the fractionation of third dose that is critical for improved immunogenicity?

*Studying bone marrow responses*

Long-lived plasma cells that reside in bone marrow are believed to be important for the maintenance of vaccine-induced antibody responses [86-88]. Therefore, understanding the phenotypes and frequency of RH5 specific B cells in the bone marrow and their correlation with the RH5 specific B cells in peripheral blood and RH5-specific antibody responses would provide better insights into the relevant immune responses. Vaccination studies have very rarely included bone marrow sampling in their protocols so there remains this missing link in malaria vaccine immunogenicity analyses. We will investigate this gap as an exploratory analysis in this protocol. The samples will be taken from 2 adults volunteers from each of Group 1a and 1b at baseline and 4 adult volunteers from each of Group 1a and 1b at 4 weeks post final vaccinations. Therefore, each adult volunteer will undergo a single bone marrow aspiration during the entire period of study.

*Studying iron status and vaccine responses*

Recent studies in Tanzania and Mozambique children who received vaccination with RTS,S showed that anaemia dampened the immune response to the malaria vaccine [89]. Further studies with B cells revealed that iron status can significantly affect the B cell differentiation which are critical for the development of effective antibodies. It is therefore important to determine if the responses to RH5 vaccination will be affected by iron status in vaccinated individuals.

#### 8. OBJECTIVES AND OUTCOME MEASURES

This is a dose-escalation, age de-escalation randomised open label Phase Ib trial to assess the safety, tolerability and immunogenicity of RH5.1/Matrix-M. Adults (18-45 years) and infants (5-17 months) will be enrolled in the study. Standard (0,1,2-month) and delayed (0,1,6-month) dosing regimens will be evaluated and compared. Safety data will be collected for each of the vaccination regimens. The humoral and cellular immune responses generated by each of these regimens will be assessed.

| 8.1. **Objectives** | 8.2. **Outcome Measures** | 8.3. **Time point(s) of evaluation** |
|---|---|---|
| **Primary Objective**<br><br>To determine safety and tolerability of RH5.1/Matrix-M given intramuscularly using different vaccination regimes adults (18-45 years) and infants (5-17 months) residing in a malaria endemic country. | • Solicited symptoms after vaccination.<br>• Unsolicited symptoms after each vaccination.<br>• Serious adverse events during the study period. | 7-day surveillance after each vaccination.<br><br>28-day surveillance after each vaccination.<br><br>Surveillance from first dose to end of study. |
| **Secondary Objectives**<br><br>To evaluate the magnitude of humoral and cellular immune responses to RH5 in adults and infants residing in a malaria endemic country.<br><br>To evaluate the quality of humoral and cellular immune responses to RH5 in adults and infants residing in a malaria endemic country.<br><br>To evaluate the longevity of humoral and cellular immune responses to RH5 in adults and infants residing in a malaria endemic country. | • Anti-RH5 antibody titres by quantitative ELISA.<br><br>• Growth inhibition activity of IgG from vaccinees on a panel of *P. falciparum* parasites.<br><br>• Avidity of anti-RH5 antibodies by ELISA and/or other assays (to be defined).<br><br>• Cellular immune responses to RH5 by ELISpot assays and/or Flow cytometry and/or other assays (to be defined). | At baseline, and various time points after vaccination as described in Appendix A |
| **Exploratory**<br><br>To determine the frequency of RH5 specific plasma cells in the bone marrow of adults vaccinated with RH5.1/Matrix-M. | • Frequency of RH5 specific plasma cells from bone marrow aspirates<br>• Statistical analysis of secondary outcome measures of G2D with other group | • at baseline and 4 weeks after 3rd vaccinations.<br><br>• At baseline, and various time points after vaccination as |

| | | |
|---|---|---|
| | | described in Appendix A |
| To assess impact of malaria exposure status on vaccine responses. | Comparisons of RH5 specific immune responses and GIA between volunteers from high and low malaria exposure areas. | At baseline, and various time points after vaccination as described in Appendix A |
| To assess the impact of iron status on the vaccine immune responses | Correlation between vaccinnee's iron status and RH5 specific immune responses and GIA. | At baseline, and various time points after vaccination as described in Appendix A |

### 9. TRIAL DESIGN

#### 9.1. Summary of trial design

- Experimental design: Phase Ib, open label, age de-escalation dose-escalation trial.
- Healthy adults (18-45 years) and infants (5-17 months) will be screened; those determined to be eligible, based on the inclusion and exclusion criteria, will be enrolled in the study.
- Infants will be classified into high and low malaria exposed based on anti-schizont IgG. Seropositivity will be defined as ELISA OD values+3SDabove the mean OD values of healthy malaria-naïve UK adults. Classification into high or low malaria exposure will be based on ELISA OD being above or below the local population mean (from previous malaria vaccine trials of infants between the age of 5 and 17 months).
- Route of administration: all vaccines will be administered by the intramuscular route to the left deltoid.
- Each participant will be observed for at least 1 hour after vaccination to evaluate and treat any acute adverse events (AEs).
- There will be 7-day follow-up period for solicited AEs post-vaccination: Day 0, 2 and 7 evaluations will be carried out by the study clinician at the study centre and day 1, 3, 4, 5 and 6 evaluations will be carried out by a trained community health worker in the participant's home, after each vaccination.
- There will be a 30-day (day of vaccination and 29 subsequent days) follow-up after each vaccine dose for reporting unsolicited symptoms.
- Serious adverse events (SAEs) will be recorded throughout the study period from administration of the first dose of RH5.1/Matrix-M to 2-2.5 years after the first vaccination.
- Immunogenicity (humoral and cellular immune responses) of RH5 as secondary outcome of interest will be evaluated at baseline and on specific timelines as shown in the outline of study procedures. As an exploratory objectives, frequency of RH5 specific plasma cells in the bone marrow of adults vaccinated with RH5.1/Matrix-M will also be evaluated.
- The duration of involvement in the study from enrolment will be approximately 2-2.5 years. The vaccination phase of the entire study takes 9 weeks and the post-vaccination follow-up lasts for up to two-two and half years after the first dose.
- The vaccine research teams will ensure that insecticide treated bed net is provided to all the study participants and training about its use are conducted.
- Data collection: conventional Case Report Form (CRF) followed by transcription into the OpenClinica Electronic Data Capture system.

#### 9.2. Trial Centre

The study will take place at the IHI Clinical Trial Facility (CTF) located in Bagamoyo town. The facility has clinical consultation rooms, a pharmacy, vaccination room, data management room and an observation area for study participants. There is a resuscitation room that is equipped with oxygen, suction, defibrillator and resuscitation kits. The facility is supported by Bagamoyo Research and Training Centre (BRTC) laboratory located within the grounds of Bagamoyo District Hospital (BDH) and Kingani compound in Bagamoyo. BRTC laboratory provides routine haematology, biochemistry parasitology and microbiology services. The laboratories can also conduct a wide range of immunology and biomedical analyses.

The clinical trial facility has experienced trained staff including pharmacist, physicians and highly experienced nurses. The facility has Quality Assurance department responsible for ensuring trials are conducted according to protocol and relevant SOP and data collected is of high quality and integrity. Study physician and nurses are ACLS and PALS certified and are involved in regular re-training activities involving simulations.

All participants who require outpatient treatment will be encourage to come to CTF during the day. All cases that require inpatient care will be seen at BDH. A clinician will be accessible 24 hours a day by phone to provide medical care to participants as needed outside the scheduled study visits. BDH is a public hospital providing secondary level health care services. The District Hospital has one paediatric ward and a general ward. There is facility for basic radiology and ultrasound. Routine paediatric surgery is carried out at the District Hospital. Referral to tertiary level facilities is rarely required, but in cases where such referrals are required participants will be sent to either Aga Khan Hospital or Muhimbili National Hospital in Dar es Salaam. The cost of referral and treatment will be covered by the study. All SAEs will be managed in the inpatient wards in BDH or other tertiary hospital if required.

#### 9.3. Study Population and Malaria Epidemiology

Participants will be recruited from Bagamoyo district. Bagamoyo district is one of the 6 districts of the Pwani region of Tanzania (area 8,462.63 km$^2$). According to the 2012 Tanzania National Census, the population of the Bagamoyo District was 343,412. The population of Bagamoyo is highly mixed due to migration and settlement of different ethnic groups. The main economic activities include agriculture, fishing, trade and commerce, and tourism. Bagamoyo has a humid tropical climate with seasonal average temperature ranging from 13˚C to 30˚C with humidity as high as 98%. Rainfall ranges between 800-

1200 mm per annum. The short rain season starts from October to December while the long rain season starts from March to May. The driest months are June to September when monthly rainfall is generally less than 50 mm per month.

Malaria remains a public health problem in the district. The district prevalence of malaria among children aged 2 months to 9 years in 2012 was 13% with higher prevalence reported in the western side [66]. The prevalence is much lower in Bagamoyo town. Transmission of malaria tends to be highest between March and May.

#### 9.4. HIV Prevalence

The prevalence of HIV in Tanzania is estimated to be about 5.9% [90]. The prevalence among healthy children in the community is thought to be below 1%. A Prevention of Mother to Child Transmission of HIV program has been operational in Bagamoyo since 2010 and the Ministry of Health anti-viral program is operational. All patients must take part in extensive counselling before treatment. Highly Active Antiretroviral Therapy (HAART) is readily available to all HIV positive people at selected government hospitals.

#### 9.5. Vaccination Services

The United Republic of Tanzania provides immunization services countrywide both on routine and non-routine basis. Immunization services are coordinated by the Immunization and Vaccine Development (IVD) services. This service is available on public and private facilities free of charge. As high as 80% of a total of 5,650 health facilities in the country provides immunization services. Tanzanian policy on immunization calls for support for routine immunization to accelerate the control of vaccine preventable diseases. The following vaccines are provided: BCG, OPV, DTP-HepB-Hib (pentavalent), pneumococcal, rotavirus, Tetanus Toxoid, vitamin A, Measles Rubella and HPV. The immunization schedule in mainland Tanzania is shown in Table 1.

Table 1 Immunization Schedule in Tanzania.

| Antigen | Time of administration |
|---|---|
| BCG, OPV 0 | At birth or first contact |
| OPV1, DTP-HepB-Hib1,PCV 1, Rota 1 | 6 Weeks of age |
| OPV2, DTP-HepB-Hib2, PCV 2, Rota 2 | 10 Weeks of age |
| OPV3, DTP-HepB-Hib3 , PCV3, IPV | 14 Weeks of age |
| MR 1 | 9 Months of age |
| MR 2 | 18 Months of age |
| HPV 1 | 9 years |
| HPV 2 | 6 months after 1st dose |

The Immunization and Vaccine Development (IVD) services are provided through four main strategies. The primary strategy is through routine immunization services at health facilities. The secondary strategies include outreach services to remote communities and mop-up campaigns aimed at selected districts to capture defaulters and reach out to children missed in routine services. Finally, there are occasional national vaccination campaigns which are implemented to reach large populations in a given period, as a supplementary activity to the routine immunization in order to increase the immunity in the community.

With the exception of the measles vaccine, the infant participants (5-17 months) would have received all immunizations by the time of enrolment. However, it is possible that a few defaulters may be encountered during screening. Vaccination with the IP will be delayed for 28 days after vaccination with measles vaccine and 14 days after non-live vaccines.

#### 9.6. Rationale for Trial Design

*Administration Schedules*

A delayed fractional dose regimen of RTS,S, in which the standard 0,1,2-month immunization schedule was changed to 0,1,6-month schedule and the third dose reduced to 20% of original dose, resulted into significant increase in vaccine efficacy[91]. Similarly, delayed fractional dosing of RH5/AS01 improved the magnitude, quality and longevity of RH5.1_FL specific antibodies compared to standard dosing (Silk, Minassian et al, submitted). These results showed that it is possible to dramatically improve the protective effect of a candidate malaria vaccine by just modifying the vaccine regimen alone without altering the vaccine formulation.

In this study we will evaluate and compare three different vaccine regimens; a) a standard 0,1,2-month dosing using 10µg RH5.1; b) a delayed fractional 0,1,6-month dosing using 50µg RH5.1 in first and second, and 10µg RH5.1 in the third vaccination and; c) a delayed non-fractional 0,1,6 month dosing using 10µg RH5.1 in all vaccinations.

The regimens have been designed to allow address the question; is the delaying of the third vaccination or fractionation of the last dose that make a difference. The aim is to select the best regimen that can be evaluated in phase IIb trials.

*Route & Dose*

RH5.1/Matrix-M will be administered intramuscularly in the left deltoid. Intramuscular administration of malaria antigens has been shown to be safe and well tolerated and this is a more practical form of administration in the field. The study will determine the ideal dose and regimen based on the safety and immunogenicity data that will be collected. Dose escalation will occur only when the safety data from previous lower dose has been reviewed by the independent safety monitors and the approval given to proceed.

RH5.1/AS01 has safely been given to 65 healthy UK adults. The doses used in the study were 2µg, 10µg and 50µg of RH5.1 antigen and 50µg of AS01B. In studies conducted in healthy UK adults, malaria antigen R21 given in combination with Matrix-M showed better reactogenicity outcomes compared to when given with AS01, but produced similar immunogenicity outcomes.

Because this is the first time RH5.1/Matrix-M is administered in infants, vaccinations will start with adults before proceeding to infants.

A dose of 10µg RH5.1 will initially be used in Tanzanian adults and infants. The dose will then be escalated to 50µg RH5.1 subject to favourable safety data from previous dose. This choice is based on the good safety and tolerability data from malaria naïve adults in the UK.

Administration of equal doses of RH5.1 in adults and infants is based on safety and immunogenicity data from previous studies. Unlike medications, use of similar doses in adults and infants for the vaccine is not an uncommon phenomenon due to differences in mechanisms of action[92]. For instance the prophylactic dose for the yellow fever vaccine is 0.5ml in both adults and children (>9 months)[93,94]. Safety and reactogenicity information is more critical in selection of the vaccine doses. We have established a robust design to carefully assess the safety and reactogenicity of lower doses before proceeding to higher doses (section 9.7).

Adjuvant dose are informed by safety and immunogenicity data. Based on the preliminary data from Burkina Faso and Kenya as well as the recommendation of the adjuvant manufacturer, we will use 50µg in both adults and infants.

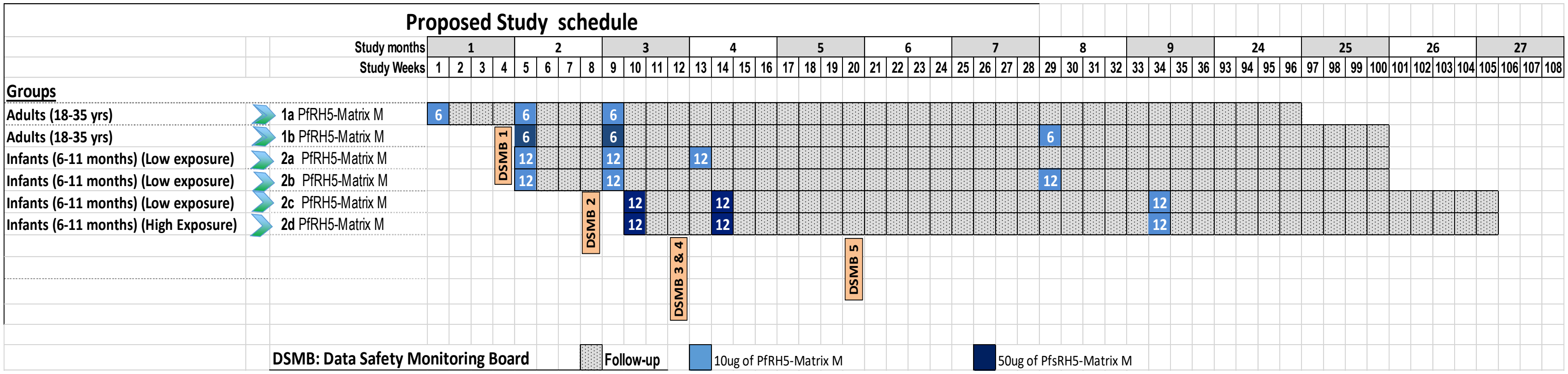

*Figure 11 : Study groups and schedule*

#### 9.7. Safety oversight during age de-escalation and dose-escalation

To assure safety, vaccinations will start in adults and will progress to infants using staggered start dates at approximately 3 weekly intervals. In infants we will start with 10µg before escalating to 50µg (Figure 11).

The approval to proceed to a higher dose within an age-group as well as age de-escalate to infants will be sought from the independent Data Safety Monitoring Board (DSMB) after the review of the safety data from the preceding dose and vaccination groups.

Progression to the higher dose or continuation of vaccination within the group may take place as long as none of the following specified criteria are met:

- Two (calculated as 33% of 6) or more participants in either Group 1a or 1b experience the same grade 3 solicited local adverse event beginning within 2 days after vaccination (day of vaccination and one subsequent day) and persisting at Grade 3 for > 72 hrs.
- Four (calculated as 33% of 12) or more participants in either Group 2a, 2b, 2c or 2d experience the same grade 3 solicited local adverse event beginning within 2 days after vaccination (day of vaccination and one subsequent day) and persisting at Grade 3 for > 72 hrs.
- Two (calculated as 33% of 6) or more participants in either Group 1a or 1b experience the same grade 3 solicited systemic adverse event beginning within 2 days after vaccination (day of vaccination and one subsequent day) and persisting at Grade 3 for > 48 hrs.
- Four (calculated as 33% of 12) or more participants in either 2a, 2b, 2c or 2d experience the same grade 3 solicited systemic adverse event beginning within 2 days after vaccination (day of vaccination and one subsequent day) and persisting at Grade 3 for > 48 hrs.
- Two (calculated as 33% of 6) or more participants in either Group 1a or 1b experience the same grade 3 unsolicited adverse event beginning within 2 days after vaccination (day of vaccination and one subsequent day) and persisting at Grade 3 for > 48 hrs that is deemed possibly, probably or definitely related to study product administration.
- Four (calculated as 33% of 12) or more participants in either Group 2a, 2b, 2c or 2d experience the same grade 3 unsolicited adverse event beginning within 2 days

after vaccination (day of vaccination and one subsequent day) and persisting at Grade 3 for > 48 hrs that is deemed possibly, probably or definitely related to study product administration.

- One participant experiences an SAE deemed possibly, probably or definitely related to study product administration.

If any of these criteria are met, all vaccinations will be put on temporary hold until guidance from the DSMB is obtained to proceed with vaccinations.

The DSMB will be convened by the study Sponsor and will be provided with a formal safety report to review, and they will provide recommendation with regard to proceeding with vaccination. The age de-escalations into infants will take place approximately 3 weeks after the adults' first vaccinations, generation of safety report and its review by the DSMB.

Details of the age de-escalation/ escalation scheme are shown in Figure 11 and also described here:

(**DSMB Review 1**) After first vaccinations have been completed in Group 1a, safety and tolerability data up until a minimum of one week will be assembled into a formal safety report and reviewed by the DSMB. If recommended by the DSMB and approved by the Sponsor's representative (CI), the trial will continue with first vaccinations of Groups 1b, 2a and 2b, and second vaccinations in Group 1a.

(**DSMB Review 2**) Safety and tolerability data up until a minimum of one week after the completion of first vaccination of Groups 1b, 2a & 2b and second vaccinations in Group 1a will be assembled into a formal safety report and reviewed by the DSMB. If recommended by the DSMB and approved by the CI, the trial will continue with first vaccinations of Groups 2c, 2d, second vaccinations of Group 1b, 2a, 2b, and third vaccinations of Group 1a.

(**DSMB Review 3**) Safety and tolerability data up until a minimum of one week after the completion of second vaccinations of Groups 1b, 2a, 2b and third vaccinations of Group 1a, will be assembled into a formal safety report and reviewed by the DSMB. If recommended by the DSMB and approved by the CI, the trial will continue with third vaccinations of Group 1b, 2a and 2b.

(**DSMB Review 4**) Safety and tolerability data up until a minimum of one week after the completion of first vaccinations of Groups 2c, 2d will be assembled into a formal safety report and reviewed by the DSMB. If recommended by the DSMB and approved by the CI, the trial will continue with second vaccinations of Groups 2c and 2d.

(**DSMB Review 5**) Safety and tolerability data up until a minimum of one week after the completion of second vaccinations of Groups 2c and 2d will be assembled into a formal safety report and reviewed by the DSMB. If recommended by the DSMB and approved by the CI, the trial will continue with the third vaccinations of Groups 2c and 2d.

DSMB Review number 3 and 4 will be done together at the same sitting.

*Sentinel Participants*

As an additional safety measure, three sentinel participants from each group will be inoculated at least 24 hours before the remaining participants are vaccinated. The three sentinel volunteers will be vaccinated in the following order; one volunteer will first be vaccinated alone and then reviewed on the day following vaccination. Providing there are no safety concerns at the day 1 review, another two volunteers may be immunised, a minimum of 2 days after the first volunteer and at least one hour apart. The remaining participants will be vaccinated if neither of the following criteria is met;

- Two participants experience at least one grade 3 solicited local adverse event beginning within 2 days after vaccination (day of vaccination and one subsequent day) and persisting at Grade 3 for > 72 hrs.
- Two participants experience at least one grade 3 solicited systemic adverse event beginning within 2 days after vaccination (day of vaccination and one subsequent day) and persisting at Grade 3 for > 48 hrs.
- Two participants experience at least one grade 3 unsolicited adverse event beginning within 2 days after vaccination (day of vaccination and one subsequent day) and persisting at Grade 3 for > 48 hrs that is deemed possibly, probably or definitely related to study product administration.
- One participant experiences an SAE deemed possibly, probably or definitely related to study product administration.

If either of these criteria are met, and before an DSMB review is called, the DSMB will review the case and advise the CI and the Investigator if they should continue with vaccinations.

### 10. PARTICIPANT IDENTIFICATION

#### 10.1. Sensitization

After approval from ethics committees, local authorities will be informed about the study in order to receive permission to approach the community. A series of meetings will then be held to explain the study to the potential participants in the community. These meetings will consist of sensitization meetings for general information and pre-screening meetings for more detailed information. The meetings will adhere to the Ministry of Health Ministry of Health, Community Development, Gender, Elderly and Children (MoHCDEC) guidelines and directives to minimize the risk of COVID-19 infection. Volunteers that previously participated in an epidemiological study conducted at IHI clinical trial facility using similar sensitization procedures and have consented to be contacted for possible participation in a study like this as per the initial informed consent, may also be invited to the sensitization meetings. The local administrative leaders will be invited to be present for these meetings. The need and the difficulties of developing a vaccine against malaria will be discussed in a culturally appropriate fashion, as well as an outline of the proposed trial, including the rationale, the background data available, the study objectives and the screening and consent procedures. Particular attention will be paid to study procedures, immunization and blood collection. To avoid misunderstandings, the purpose of blood collection and the associated risks will be explained in a culturally sensitive fashion. Opportunity will be provided for the community members to ask questions and get responses from the Investigators.

Potentially interested participants including the parents/guardians will be asked to register their names at the end of the meetings with the study staff. Potential participants will be invited to attend a pre-screening meeting at CTF.

At the pre-screening meeting the PI and trained study clinicians and nurses will provide detailed information about the study using the PIS in a group and individual sessions. A copy of the PIS will also be provided before commencing these meetings. The PI will ensure potential participants or parents/guardians fully understand the risks and benefits associated with participation in the study.

It will be stressed that RH5 is an experimental vaccine and cannot be guaranteed to provide protection and that therefore it will still be necessary to seek treatment for possible malaria even after vaccination and with the use of available preventive measures including insecticide treated bed nets. It will also be made clear that the study will require HIV testing of the participants. It will be explained that eventually the plan is to offer immunization to everyone including those testing HIV positive, but that normal practice in intervention evaluation is to begin with those who are HIV negative until the safety profile is better defined. It will be made clear that screening on enrolment will include a range of diseases (not just HIV), so exclusion from participating in the trial does not mean being HIV positive and the confidentiality of those participants infected with HIV will be protected. It will be stressed that this is the beginning of a long process of RH5 vaccine development, which will be accelerated by the conduct of similar trials in Africa.

Opportunity will be provided for the potential participants to ask questions and get responses from the PI and study physicians. Those who are still interested to take part will then be invited for a formal screening visit where an informed consent form will be signed or thumb-printed. A copy of the PIS will be given to the potential participants to take home and read it further including consulting their spouses or relatives.

##### 10.2. Trial Participants

The study will recruit healthy young adults (18-45 years) and infants (5-17 months) residing in Bagamoyo district in Tanzania. The participants will be enrolled into two groups for adults and 4 groups for infants as shown in Figure 11.

##### 10.3. Inclusion Criteria

Only participants who meet all the inclusion criteria will be enrolled into the trial;

- Group 1: Healthy male or female adults aged 18-45 years at the time of enrolment with signed consent.
- Group 1 (Female only participants): Must be non-pregnant (as demonstrated by a negative urine pregnancy test), and practice continuous effective contraception* for the duration of the study.
  * Female volunteers are required to use an effective form of contraception during the course of the study as this is a Phase I, study and there is currently no information about the effect of this vaccine on a foetus. Acceptable forms of contraception for female volunteers include:
    - Established use of oral, injected or implanted hormonal methods of contraception.
    - Placement of an intrauterine device (IUD) or intrauterine system (IUS).
    - Total abdominal hysterectomy.
- Groups 2a, 2b, 2c & 2d: Healthy male or female infants aged 5-17 months at the time of enrolment with signed consent obtained from parents or guardians.
- Planned long-term (at least 24 months from the date of recruitment) or permanent residence in the study area.
- Adults with a Body Mass Index (BMI) 18 to 30 Kg/$m^2$; or infants with Z-score of weight-for-age within ±2SD.

##### 10.4. Exclusion Criteria

The participant may not enter the trial if ANY of the following apply:

- Clinically significant congenital abnormalities as judged by the PI or other delegated individual.

- Clinically significant history of skin disorder (psoriasis, contact dermatitis etc.), allergy, cardiovascular disease, respiratory disease, endocrine disorder, liver disease, renal disease, gastrointestinal disease and neurological illness as judged by the PI or other delegated individual.
- Any confirmed or suspected immunosuppressive or immunodeficient state, including HIV infection; asplenia; recurrent, severe infections and chronic (more than 14 days) immunosuppressant medication within the past 6 months (inhaled and topical steroids are allowed).
- History of cancer (except basal cell carcinoma of the skin and cervical carcinoma in situ).
- Weight for age z-scores below 2 standard deviations of normal for age.
- History of allergic disease or reactions likely to be exacerbated by any component of the vaccines, e.g. egg products, Kathon, neomycin, betapropiolactone.
- Any history of anaphylaxis in relation to vaccination.
- Clinically significant laboratory abnormality as judged by the PI or other delegated individual.
- Blood transfusion within one month of enrolment.
- History of vaccination with previous experimental malaria vaccines.
- Administration of immunoglobulins and/or any blood products within the three months preceding the planned administration of the vaccine candidate.
- Participation in another research study involving receipt of an investigational product in the 30 days preceding enrolment, or planned use during the study period.
- Seropositive for hepatitis B surface antigen (HBsAg) or hepatitis C (HCV IgG).
- Any other finding which in the opinion of the PI or other delegated individual would increase the risk of an adverse outcome from participation in the trial.
- Likelihood of travel away from the study area.
- Positive malaria by blood smear at screening.
- Female participant who is pregnant, lactating or planning pregnancy during the course of the trial.
- Scheduled elective surgery or other procedures requiring general anaesthesia during the trial.
- Any other significant disease, disorder or situation which, in the opinion of the Investigator, may either put the participants at risk because of participation in the trial, or may influence the result of the trial, or the participant's ability to participate in the trial.

##### 10.5. **Contraindications to subsequent vaccination**

*<u>Indications for deferral of vaccination</u>*

These events constitute contraindications to administration of the IP at that point in time and will result in deferral of vaccination to another time within the protocol specified window, or withdrawal at the discretion of the Investigator. AEs should be followed-up according to the instructions in Section 13.

- Acute disease at the time of administration of the IP (acute disease is defined as the presence of a moderate or severe illness with or without fever and symptoms suggestive of possible COVID-19 disease). All vaccines can be administered to persons with a minor illness such as diarrhoea or mild upper respiratory infection without fever, i.e. axillary temperature < 37.5°C.
- Axillary temperature of ≥ 37.5°C.
- Abnormal laboratory parameters that have been determined to be clinically significant.
- Diphtheria, Tetanus, Pertussis (whole-cell), *Hemophilus influenzae* type B, Measles, or Polio vaccination within 14 days of schedule to receive the IP.
- On request of the participant.
- On advice of the safety monitor.
- On advice of the Investigators.

*Permanent Contraindications for further vaccination*

The following AEs constitute absolute contraindications to further administration of the IP. If any of these AEs occur during the study, the participant must not receive additional doses of vaccine, but will continue with the full safety monitoring procedure (if the participant has already received a vaccination) as per protocol:

- Acute allergic reaction, significant IgE-mediated event or anaphylactic shock following the administration of vaccine investigational product.
- Any confirmed or suspected immunosuppressive or immunodeficient condition, including human immunodeficiency virus (HIV) infection.
- The participant experiences an SAE that is determined to be related to the study product.
- The participant starts taking a concomitant medication for a chronic illness.
- Pregnancy.
- On request of the participant.
- On advice of the Independent DSMB.
- On advice of the Investigators.
- On advice of the EC, regulatory authority or DSMB.

*Withdrawal of study participant*

Participants have a right to withdraw from the study at any time and are not obliged to give his or her reasons for doing so. The Investigator may withdraw the participant at any time in the interests of the participant's health and well-being. In addition, the participant may withdraw/be withdrawn for any of the following reasons:

- Any serious adverse event (SAE) deemed to be related to the IP.
- Any adverse event (AE) that, according to the clinical judgment of the Investigator, is considered a contraindication to proceeding with the study procedures.
- The use of concomitant, chronic medication active on the immune system (steroids, immunosuppressive agents) or potentially affecting RH5.1/Matrix-M.
- Participant completely lost to follow-up.
- Poor compliance with study procedures, such that completeness or integrity of data are jeopardized.
- Administrative decision by the Investigator.
- Ineligibility (either arising during the study or retrospectively, having been overlooked at screening).
- Significant protocol deviation.

If a participant withdraws from the study, blood samples collected before withdrawal will be used/ stored to address the safety and immunogenicity objectives of the current protocol unless the participant specifically requests otherwise. All participants can withdraw their permission to store the samples for future studies at any time.

The reason for withdrawal will be recorded in the CRF. If the participant is withdrawn due to an adverse event, the Investigator will arrange for follow-up visits or telephone calls until the adverse event has resolved or stabilised.

*Replacement of Individual Participants after Withdrawal*

If for any reason a participant cannot receive the first vaccination, they can be replaced by a backup participant that has been screened and found to be eligible. This can only happen on the day of the first RH5 vaccination for all the groups. However, if the participant is withdrawn after receiving the firstRH5, there will be no replacement.

##### 10.6. **Pregnancy**

Should a participant become pregnant during the trial, she will not receive further vaccination but will be followed up as other participants, and in addition will be followed until pregnancy outcome, with the participant’s permission. We will not routinely perform venepuncture on such participants.

##### 10.7. **Managing volunteers during COVID-19 outbreak**

Reports from authorities indicates that hospitalized COVID-19 cases have declined significantly. However, the population is still being urged to take all necessary precautions as the transmission in still ongoing.

According to the Ministry of Health, Community Development, Gender, Elderly and Children (MoHCDEC), a suspected COVID-19 case will be defined as[95]:

- Fever of measured temperature (axillary temperature ≥38 C°) or history of fever and acute respiratory infection (sudden onset of respiratory infection with at least

one of the following: shortness of breath, cough or sore throat) with or without anosmia and ageusia.
OR

- Severe acute respiratory infection requiring admission to hospital with clinical or radiological evidence of pneumonia or acute respiratory distress syndrome (i.e. even if no evidence of fever).

All participants who present with symptoms suggestive of SARS-COV2 at the screening, during vaccinations and follow-up will be managed as shown by guidelines in *Table 2*. All volunteers who require COVID-19 testing will be referred to public testing facilities where samples will be collected for testing at National Public Health Laboratory. The study will cover the costs of treatment of volunteers who are diagnosed with COVID-19. The management of COVID-19 will follow the guidelines developed by the Ministry of Health, Community Development, Gender, Elderly and Children (MoHCDEC). Mild and moderate cases that don't require hospitalization will be managed by study doctors while cases requiring admission will be referred to the Government treatment centres.

*Table 2: Guidelines on management of COVID-19 suspect cases*

| **Situation** | **Action** |
|---|---|
| Asymptomatic infection at any time | No need for testing. Risk to staff can be mitigated by PPE. |
| Symptomatic at D0 and at subsequent vaccinations: | Advice and facilitate COVID-19 testing at Public testing facility.<br>If positive for COVID-19 and volunteer has moderate to severe symptoms<br>• Postpone vaccination for a minimum of 14 days.<br>• Vaccinate if symptoms resolve and afebrile. (Volunteer need to be afebrile for 24 hrs before vaccination).<br>If negative for COVID-19<br>• Postpone vaccination if symptoms are moderate or severe and if febrile.<br>• Vaccinate if symptoms resolve or become mild and afebrile. (Volunteer need to be afebrile for 24 hrs before vaccination). |
| Symptomatic at follow-up visits | If meets case definition for suspect COVID-19:<br>• Advise and facilitate COVID-19 testing at Public testing facility<br>• Follow-up the patient by phone and discourage visit at study clinic while awaiting test results.<br>If positive for COVID-19<br>• Advise patient to isolate for 14 days and prescribe supportive treatment.<br>• Postpone sample collection until better<br>• Continue follow up with home visit by study staff with PPE.<br>If negative for COVID-19<br>• Continue with follow-up as planned |

### 11. TRIAL PROCEDURES

#### 11.1. Informed Consent

The participant (Group 1) and parent/guardian of the participant (Groups 2) must personally sign and date the latest approved version of the Informed Consent form before any trial specific procedures are performed.

Written versions of the Participant Information Sheet (PIS) and Informed Consent Form (ICF) will be presented to the participants detailing no less than: the exact nature of the trial; what it will involve for the participant; the implications and constraints of the protocol; the known side effects and any risks involved in taking part. It will be clearly stated that the participant is free to withdraw from the trial at any time for any reason without prejudice to future care, without affecting their legal rights and with no obligation to give the reason for withdrawal.

All participants (Group 1) and parents/guardians of the participants (Groups 2) will be required to demonstrate their understanding of the study by answering a questionnaire which will consist of 10 true/false statements that will be instituted by study personnel. Participants must respond correctly to all 10 questions. They will have a maximum of two attempts after which those who fail to correctly answer all questions will be excluded. Those who correctly respond to all 10 questions will be allowed to sign the ICF.

The participant will be allowed as much time as wished to consider the information, and the opportunity to question the Investigator, or other independent parties, to decide whether they will participate in the trial. Written Informed Consent will then be obtained by means of participant-dated signature or thumb-print and dated signature of the person who presented and obtained the Informed Consent. If the participant is illiterate a literate witness will be present during the consent process and verify the process by signing and dating the ICF. Consenting will be done by trained study staff including the clinicians and nurses. A copy of the signed ICF will be given to the participant. The original signed form will be retained at the trial site.

#### 11.1. Screening and Eligibility Assessment

Screening procedures will only start once a participant has provided a written informed consent and continue until all 60 participants and at least 2 back-up participants have been enrolled into the study. Two screening visits will be conducted for each participant.

*Screening Visit 1*

Only participants who meet eligibility criteria and have signed/thumb-printed the informed consent will undergo a full screening procedure. At screening visit 1, study staff will go through the PIS for the second time and describe the study in detail. This will be done in a group session where potential participants will be encouraged to ask questions. After the group session each participant will have a private session with study staff where additional clarification will be provided and time for more questions provided.

Participants will be assigned with a study ID on a first come first serve basis. A photo will also be taken to allow preparation of Study ID cards which will be given after the first vaccination. In the interim all participants will receive temporary study ID forms.

The following procedures will then take place:

- Taking medical history of a participant.
- Complete physical examination including vital signs, height, weight and blood pressure.
- Venous blood sample collection for the following analyses:
    - Haematology: Full Blood Count (CBC) which includes: red blood cells, platelets, white blood cell count, lymphocyte count, haemoglobin, mean corpuscular volume and mean corpuscular haemoglobin concentration.
    - Biochemistry: serum creatinine and ALT and Iron status tests (including but not limited to serum ferritin, transferrin saturation, soluble transferrin receptor and erythrocyte protoporphyrin).
    - Serology: HIV, HCV and HBV tests.
    - Antibody level to malaria schizonts
    - Thick blood smear
- Collection of urine for the following analyses:
    - Dipstick for blood, glucose and protein.
    - Pregnancy test (for women in Group 1 only).
- Checking of inclusion and exclusion criteria

Once all specific visit procedures are completed, participants will be allowed to go home and asked to return for Screening Visit 2.

The screening laboratory results will be reviewed by study clinicians before participants are seen at screening visit 2. These procedures will be documented in the case report forms (CRFs) and relevant clinical notes, which will be kept in the individual study participant’s file. After collection of all screening information, the study team will review and assess the eligibility of each participant. All participants will be contacted and informed of the specific date to come to CTF for Screening Visit 2.

*Screening Visit 2*

During Screening Visit 2, participants will be informed of their screening results and receive HIV and hepatitis post-test counselling. Study clinicians will inform them if they are eligible for enrolment into the study. The principal investigator (PI) will have the final decision on the eligibility of a participant. Only those laboratory results with clinical relevance will be provided to the participant. Immunology results will be summarized and presented as part of final feedback to participants at the end of the study. Participants excluded from this trial because of clinically significant medical conditions may be managed initially by study clinicians and/or referred to the routine health care system for further evaluation and treatment as necessary. In the event that a participant tests positive for HIV, HBV, or HCV, he or she will be referred to the appropriate public health facility for further counselling and treatment. Participants (and their parents/guardians if applicable) will be provided with ITNs at this visit and will be encouraged to use them to reduce the risk of natural malaria infection.

Recruitment will continue until all the required number of participants per Group has fulfilled all inclusion criteria and none of the exclusion criteria. Should the participant or the parents of participants change their mind and decline to participate prior to immunization, additional participants will be screened the required number of participants per group is enrolled. A list of eligible participants per group will be generated and used to identify the participants eligible for immunization. Screening tests will be completed within 30 days prior to immunization.

#### 11.2. Baseline Assessments

Before vaccination all participants with undergo baseline assessment. This will include collection of safety and immunology samples. Once all final eligibility criteria are reviewed including safety laboratory results, eligible participants will be vaccinated. The window period as shown in the Appendix A will be used in case a participant cannot be available on a scheduled date. Pre-vaccination laboratory assessments will be done within 24 hours prior to vaccination. However only clinical assessments and vital signs collected on the day of vaccination will be recorded in the pre-vaccination CRF.

#### 11.3. Randomisation procedure

There will be no randomization in this study. Volunteer will be recruited into the groups as they become eligible. Study ID will be assigned to participants in the order in which they are enrolled in the trial. Once assigned, the group and the dose regimens will be known to volunteers and investigators.

#### 11.4. Blinding

This is an open label trial.

#### 11.5. Detailed scheduled study visits

**For standard schedule vaccination Groups**

**<u>Day 0: (First Vaccination with RH5.1/Matrix-M)</u>**

Participants will first be identified using their Study ID cards and vital signs and body weight will be measured. Blood sample (if not already done within the last 48 hours) will be collected for the following analyses:

- haematology (full blood count);
- Thick smear for malaria parasitemia
- biochemistry (including creatinine and ALT);
- Immunology (including RH5 specific humoral and T cell responses) and HLA typing
- Immunology (gene expression profiling) in adults only.

In addition, a bone marrow aspirate sample will be collected (as specified in study specific SOP) from 2 volunteers from Group 1a. The two volunteers will be randomly selected and will undergo only one bone marrow aspiration during the entire study period. Volunteers will have Prothrombin Time/INR and platelets tests within one week before bone marrow aspiration to rule out any bleeding disorder.

Medical history and focused physical examination (informed by complaints) will be conducted by study clinicians who will also check for inclusion/exclusion criteria, contraindications to vaccination and elimination criteria. Safety laboratory results will be reviewed by study clinician before vaccination and checked by the PI (or designee). If participant is confirmed to be eligible they will be vaccinated according to vaccination SOP.

RH5.1/Matrix-M vaccine will then be administered intramuscularly in the left deltoid by trained study staff according to the site vaccination SOP. Each participant will be assessed for at least 1 hour after vaccination to evaluate and treat any acute adverse events. During this time the following will be recorded;

- Post vaccination vital signs
- Post-vaccination solicited signs/symptoms.
- Post-vaccination unsolicited adverse events.
- Post-vaccination SAEs.
- Any concomitant medication.

Day 1; Post-Vaccination with RH5.1/Matrix-M)- Group 1a only

Participants will be seen at CTF where vital signs and temperature monitoring will be performed and recorded in the CRF. AEs will be recorded in the CRF. Blood sample will be collected for the analysis of:

- Immunology (gene expression profiling) in adults only.

Day 2 (-1/+1) days; Post-Vaccination with RH5.1/Matrix-M)

Participants will be seen at CTF where medical history, temperature monitoring and physical examination will be performed and recorded in the CRF. AEs will be recorded in the CRF.

Days 3, 4, 5 and 6 (Post-Vaccination with RH5.1/Matrix-M)

Each participant will be visited at home on 3, 4, 5 and 6 by a trained study staff for assessment and recording of any solicited and unsolicited AEs. If necessary, the participant will continue to be seen regularly until the AEs have resolved or stabilised. During the home visit, the following will be assessed and recorded in the CRF;

- Axillary body temperature.
- Local and general solicited adverse events.
- SAEs experienced by the participant since the last visit.
- Unsolicited adverse events experienced by the participant since the last visit.
- Concomitant medication taken.

Day 7 (-1/+3 days; Post-Vaccination with RH5.1/Matrix-M)

Participants will be seen at CTF where medical history, temperature monitoring and physical examination will be performed and recorded in the CRF. AEs will be recorded in the CRF.

Blood sample will be collected for the analysis of:

- haematology (full blood count);
- biochemistry (including creatinine and ALT);

Day 14 (-1/+3 days; Post-Vaccination with RH5.1/Matrix-M)

Participants will be seen at CTF where medical history, temperature monitoring and physical examination will be performed and recorded in the CRF. AEs will be recorded in the CRF.

Blood sample will be collected for the analysis of:

- haematology (full blood count);
- biochemistry (including creatinine and ALT);
- Immunology (including RH5 specific humoral and T cell responses).

**Day 28 (Second Vaccination with RH5.1/Matrix-M) (-7/+14 days);**

Participants will first be identified using their Study ID cards and vital signs and body weight will be measured. Medical history and focused physical examination (informed by complaints) will be conducted by study clinicians who will also check for contraindications to vaccination and elimination criteria.

Blood sample will be collected for the pre-vaccination assessment of (if not already done within the last 48 hours):

- haematology (full blood count);
- biochemistry (including creatinine and ALT);
- Immunology (including RH5 specific humoral and T cell responses)
- Immunology (gene expression) in adults only.

- symptomatic malaria parasitaemia (only if participant is symptomatic)
- asymptomatic malaria parasitaemia (retrospectively);

Safety laboratory results will be reviewed by the study clinician before vaccination and checked by the PI (or designee).

RH5.1/Matrix-M vaccine will then be administered intramuscularly in the left deltoid by trained study staff according to the site vaccination SOP. Each participant will be assessed for at least 1 hour after vaccination to evaluate and treat any acute adverse events. During this time the following will be recorded;

- Post vaccination vital signs
- Post-vaccination solicited signs/symptoms.
- Post-vaccination unsolicited adverse events.
- Post-vaccination SAEs.
- Any concomitant medication.

Day 29; Post-Vaccination with RH5.1/Matrix-M) –Group 1aonly

Participants will be seen at CTF where vital signs and temperature monitoring will be performed and recorded in the CRF. AEs will be recorded in the CRF. Blood sample will be collected for the analysis of:

- Immunology (gene expression profiling) in adults only.

Day 30 (-1/+1) days; Post-Vaccination with RH5.1/Matrix-M)

Participants will be seen at CTF where medical history, temperature monitoring and physical examination will be performed and recorded in the CRF. AEs will be recorded in the CRF.

Days 31, 32, 33 and 34 (Post-Vaccination with RH5.1/Matrix-M)

Each participant will be visited at home on day 1, 3, 4, 5 and 6 by a trained study staff for assessment and recording of any solicited and unsolicited AEs. If necessary, the participant will continue to be seen regularly until the AEs have resolved or stabilised. During the home visit, the following will be assessed and recorded in the CRF;

- Axillary body temperature.
- Local and general solicited adverse events.
- SAEs experienced by the participant since the last visit.
- Unsolicited adverse events experienced by the participant since the last visit.
- Concomitant medication taken.

Day 35 (-1/+3 days; Post-Vaccination with RH5.1/Matrix-M)

Participants will be seen at CTF where medical history, temperature monitoring and physical examination will be performed and recorded in the CRF. AEs will be recorded in the CRF.

Blood sample will be collected for the analysis of:

- haematology (full blood count);
- biochemistry (including creatinine and ALT);
- Immunology

Day 42 (-1/+3 days; Post-Vaccination with RH5.1/Matrix-M)

Participants will be seen at CTF where medical history, temperature monitoring and physical examination will be performed and recorded in the CRF. AEs will be recorded in the CRF.

Blood sample will be collected for the analysis of:

- haematology (full blood count);
- biochemistry (including creatinine and ALT);
- Immunology (including RH5 specific humoral and T cell responses)

**Day 56 (Third vaccination with RH5.1/Matrix-M) (-7/+14 days);**

Participants will first be identified using their Study ID cards and vital signs and body weight will be measured. Medical history and focused physical examination (informed by complaints) will be conducted by study clinicians who will also check for contraindications to vaccination and elimination criteria.

Blood sample will be collected for the pre-vaccination assessment of (if not already done within the last 48 hours):

- haematology (full blood count);
- biochemistry (including creatinine and ALT);
- Immunology (including RH5 specific humoral and T cell responses)
- Immunology (gene expression) in adults only
- symptomatic malaria parasitaemia (only if participant is symptomatic)
- asymptomatic malaria parasitaemia (retrospectively);

Safety laboratory results will be reviewed by the study clinician before vaccination and checked by the PI (or designee).

RH5.1/Matrix-M vaccine will then be administered intramuscularly in the left deltoid by trained study staff according to the site vaccination SOP. Each participant will be assessed for at least 1 hour after vaccination to evaluate and treat any acute adverse events. During this time the following will be recorded;

- Post vaccination vital signs
- Post-vaccination solicited signs/symptoms.
- Post-vaccination unsolicited adverse events.
- Post-vaccination SAEs.
- Any concomitant medication.

Day 57; Post-Vaccination with RH5.1/Matrix-M) –Group 1a only

Participants will be seen at CTF where vital signs and temperature monitoring will be performed and recorded in the CRF. AEs will be recorded in the CRF. Blood sample will be collected for the analysis of:

- Immunology (gene expression profiling) in adults only.

Day 58 (-1/+1 days; Post-Vaccination with RH5.1/Matrix-M)

Participants will be seen at CTF where medical history, temperature monitoring and physical examination will be performed and recorded in the CRF. AEs will be recorded in the CRF.

Days 59, 60, 61 and 62 (Post-Vaccination with RH5.1/Matrix-M)

Each participant will be visited at home daily on days 57, 59, 60, 61 and 62 by a community health worker (CHW) for assessment and recording of any solicited and unsolicited AEs. If necessary, the participant will continue to be seen regularly until the AEs have resolved or stabilised. CHW will be blinded to the group allocation of the subject. During the home visit, the following will be assessed and recorded in the CRF;

- Axillary body temperature.
- Local and general solicited adverse events.
- SAEs experienced by the vaccinees since the last visit.
- Unsolicited adverse events experienced by the vaccinees since the last visit.
- Concomitant medication taken.

Day 63 (-1/+3 days; Post-Vaccination with RH5.1/Matrix-M)

Participants will be seen at CTF where medical history, temperature monitoring and physical examination will be performed and recorded in the CRF. AEs will be recorded in the CRF.

Blood will be collected for analysis of:

- haematology (full blood count);
- biochemistry (including creatinine and ALT);
- symptomatic malaria parasitaemia (only if participant is symptomatic);
- asymptomatic malaria parasitaemia (retrospectively);
- Immunology:

Day 70 (-1/+3 days; Post-Vaccination with RH5.1/Matrix-M)

Participants will be seen at CTF where medical history, temperature monitoring and physical examination will be performed and recorded in the CRF. AEs will be recorded in the CRF.

Blood will be collected for analysis of:

- haematology (full blood count);
- biochemistry (including creatinine and ALT);
- symptomatic malaria parasitaemia (only if participant is symptomatic);
- asymptomatic malaria parasitaemia (retrospectively);
  Immunology (including RH5 specific humoral and T cell responses)

Day 84 (-7/+2 days; Post-Vaccination with RH5.1/Matrix-M)

Participants will be seen at CTF where medical history, temperature monitoring and physical examination will be performed and recorded in the CRF. AEs will be recorded in the CRF.

Blood sample will be collected for the analysis of:

- haematology (full blood count);
- biochemistry (including creatinine and ALT);
- Immunology (including RH5 specific humoral and T cell responses and gene expression profiles of immune cells)
- symptomatic malaria parasitaemia (only if participant is symptomatic);
- asymptomatic malaria parasitaemia (retrospectively);

In addition, a bone marrow aspirate sample will be collected (as specified in study specific SOP) in the remaining 4 volunteers from Group 1a who did not undergo bone marrow aspiration at Day 0. Volunteers will have Prothrombin Time/INR and platelets tests within one week before bone marrow aspiration to rule out any bleeding disorder. Bone marrow aspiration will not be conducted in PT/INR are abnormal.

Day 140 (±7 days; Post-Vaccination with RH5.1/Matrix-M)

Medical history, temperature monitoring and physical examination will be performed and recorded in the CRF. AEs will be recorded in the CRF.

Blood sample will be collected for the analysis of:

- haematology (full blood count);
- biochemistry (including creatinine and ALT);
- symptomatic malaria parasitaemia (only if participant is symptomatic);

- asymptomatic malaria parasitaemia;
- Immunology (including RH5 specific humoral and T cell responses)

Day 168 (± 7days; Post-Vaccination with RH5.1/Matrix-M)

Medical history, temperature monitoring and physical examination will be performed and recorded in the CRF. AEs will be recorded in the CRF.

Blood sample will be collected for the analysis of:

- haematology (full blood count);
- biochemistry (including creatinine and ALT);
- Immunology (including RH5 specific humoral and T cell responses)
- symptomatic malaria parasitaemia (only if participant is symptomatic);
- asymptomatic malaria parasitaemia;

Year 1 (± 28 days; Post-Vaccination with RH5.1/Matrix-M)

Medical history, temperature monitoring and physical examination will be performed and recorded in the CRF. AEs will be recorded in the CRF.

Blood sample will be collected for the analysis of:

- haematology (full blood count);
- biochemistry (including creatinine and ALT);
- Immunology (including RH5 specific humoral and T cell responses)
- symptomatic malaria parasitaemia (only if participant is symptomatic);
- asymptomatic malaria parasitaemia;

Year 2 (± 28 days; Post-Vaccination with RH5.1/Matrix-M)

Medical history, temperature monitoring and physical examination will be performed and recorded in the CRF. AEs will be recorded in the CRF.

Blood sample will be collected for the analysis of:

- haematology (full blood count);
- biochemistry (including creatinine and ALT);
- Immunology (including RH5 specific humoral and T cell responses)
- symptomatic malaria parasitaemia (only if participant is symptomatic);
- asymptomatic malaria parasitaemia;

**Delayed Dosing regimen**

**Day 0: (First Vaccination with RH5.1/Matrix-M)**

Participants will first be identified using their Study ID cards and vital signs and body weight will be measured. Blood sample (if not already done within the last 48 hours) will be collected for the following analyses:

- haematology (full blood count);
- biochemistry (including creatinine and ALT);
- Immunology (including RH5 specific humoral and T cell responses)
- Immunology (gene expression) in adults only.

In addition, a bone marrow aspirate sample will be collected (as specified in study specific SOP) in 2 volunteers from Group 1b. Volunteers will be randomly selected randomly and each volunteer will undergo only a single bone marrow aspiration during the entire study period. Volunteers will have Prothrombin Time/INR and platelets tests within one week before bone marrow aspiration to rule out any bleeding disorder. Bone marrow aspiration will not be conducted in PT/INR are abnormal.

Medical history and focused physical examination (informed by complaints) will be conducted by study clinicians who will also check for inclusion/exclusion criteria, contraindications to vaccination and elimination criteria. Safety laboratory results will be reviewed by study clinician before vaccination and checked by the PI (or designee). If participant is confirmed to be eligible they will be vaccinated according to vaccination SOP.

RH5.1/Matrix-M vaccine will then be administered intramuscularly in the left deltoid by trained study staff according to the site vaccination SOP. Each participant will be assessed for at least 1 hour after vaccination to evaluate and treat any acute adverse events. During this time the following will be recorded;

- Post vaccination vital signs
- Post-vaccination solicited signs/symptoms.
- Post-vaccination unsolicited adverse events.
- Post-vaccination SAEs.
- Any concomitant medication.

Day 1; Post-Vaccination with RH5.1/Matrix-M)-Group 1b only

Participants will be seen at CTF where vital signs and temperature monitoring will be performed and recorded in the CRF. AEs will be recorded in the CRF.

Blood sample will be collected for the analysis of:

- Immunology (gene expression profiling) in adults only.

Day 2 (-1/+1) days; Post-Vaccination with RH5.1/Matrix-M)

Participants will be seen at CTF where medical history, temperature monitoring and physical examination will be performed and recorded in the CRF. AEs will be recorded in the CRF.

Days 3, 4, 5 and 6 (Post-Vaccination with RH5.1/Matrix-M)

Each participant will be visited at home on day 1, 3, 4, 5 and 6 by a trained study staff for assessment and recording of any solicited and unsolicited AEs. If necessary, the participant will continue to be seen regularly until the AEs have resolved or stabilised. During the home visit, the following will be assessed and recorded in the CRF;

- Axillary body temperature.
- Local and general solicited adverse events.
- SAEs experienced by the participant since the last visit.
- Unsolicited adverse events experienced by the participant since the last visit.
- Concomitant medication taken.

Day 7 (-1/+3 days; Post-Vaccination with RH5.1/Matrix-M)

Participants will be seen at CTF where medical history, temperature monitoring and physical examination will be performed and recorded in the CRF. AEs will be recorded in the CRF.

Blood sample will be collected for the analysis of:

- haematology (full blood count);
- biochemistry (including creatinine and ALT);

Day 14 (-1/+3 days; Post-Vaccination with RH5.1/Matrix-M)

Participants will be seen at CTF where medical history, temperature monitoring and physical examination will be performed and recorded in the CRF. AEs will be recorded in the CRF.

Blood sample will be collected for the analysis of:

- haematology (full blood count);
- biochemistry (including creatinine and ALT);
- Immunology:

**Day 28 (Second Vaccination with RH5.1/Matrix-M) (-7/+14 days);**

Participants will first be identified using their Study ID cards and vital signs and body weight will be measured. Medical history and focused physical examination (informed by complaints) will be conducted by study clinicians who will also check for contraindications to vaccination and elimination criteria.

Blood sample will be collected for the pre-vaccination assessment of (if not already done within the last 48 hours):

- haematology (full blood count);
- biochemistry (including creatinine and ALT);

- Immunology (including RH5 specific humoral and T cell responses)
- Immunology (gene expression) in adults only.
- symptomatic malaria parasitaemia (only if participant is symptomatic)
- asymptomatic malaria parasitaemia (retrospectively);
- malaria parasite genotyping (retrospectively);

Safety laboratory results will be reviewed by the study clinician before vaccination and checked by the PI (or designee).

RH5.1/Matrix-M vaccine will then be administered intramuscularly in the left deltoid by trained study staff according to the site vaccination SOP. Each participant will be assessed for at least 1 hour after vaccination to evaluate and treat any acute adverse events. During this time the following will be recorded;

- Post vaccination vital signs
- Post-vaccination solicited signs/symptoms.
- Post-vaccination unsolicited adverse events.
- Post-vaccination SAEs.
- Any concomitant medication.

<u>Day 29; Post-Vaccination with RH5.1/Matrix-M)-Group 1b only</u>

Participants will be seen at CTF where vital signs and temperature monitoring will be performed and recorded in the CRF. AEs will be recorded in the CRF. Blood sample will be collected for the analysis of:

- Immunology (gene expression profiling) in adults only

<u>Day 30 (-1/+1) days; Post-Vaccination with RH5.1/Matrix-M)</u>

Participants will be seen at CTF where medical history, temperature monitoring and physical examination will be performed and recorded in the CRF. AEs will be recorded in the CRF.

<u>Days 31, 32, 33 and 34 (Post-Vaccination with RH5.1/Matrix-M)</u>

Each participant will be visited at home on day 1, 3, 4, 5 and 6 by a trained study staff for assessment and recording of any solicited and unsolicited AEs. If necessary, the participant will continue to be seen regularly until the AEs have resolved or stabilised. During the home visit, the following will be assessed and recorded in the CRF;

- Axillary body temperature.
- Local and general solicited adverse events.
- SAEs experienced by the participant since the last visit.
- Unsolicited adverse events experienced by the participant since the last visit.
- Concomitant medication taken.

Day 35 (-1/+3 days; Post-Vaccination with RH5.1/Matrix-M)

Participants will be seen at CTF where medical history, temperature monitoring and physical examination will be performed and recorded in the CRF. AEs will be recorded in the CRF.

Blood sample will be collected for the analysis of:

- haematology (full blood count);
- biochemistry (including creatinine and ALT);
- Immunology (including RH5 specific humoral and T cell responses)

Day 42 (-1/+3 days; Post-Vaccination with RH5.1/Matrix-M)

Participants will be seen at CTF where medical history, temperature monitoring and physical examination will be performed and recorded in the CRF. AEs will be recorded in the CRF.

Blood sample will be collected for the analysis of:

- haematology (full blood count);
- biochemistry (including creatinine and ALT);
- Immunology (including RH5 specific humoral and T cell responses).

Day 56 (-1/+3 days; Post-Vaccination with RH5.1/Matrix-M)

Participants will be seen at CTF where medical history, temperature monitoring and physical examination will be performed and recorded in the CRF. AEs will be recorded in the CRF.

Blood sample will be collected for the analysis of:

- Immunology (including RH5 specific humoral and T cell responses and gene expression profiles of immune cells).

**Day 182 (Third vaccination with RH5.1/Matrix-M) (-30/+14days);**

Participants will first be identified using their Study ID cards and vital signs and body weight will be measured. Medical history and focused physical examination (informed by complaints) will be conducted by study clinicians who will also check for contraindications to vaccination and elimination criteria.

Blood sample will be collected for the pre-vaccination assessment of (if not already done within the last 48 hours):

- haematology (full blood count);
- biochemistry (including creatinine and ALT);
- Immunology (including RH5 specific humoral and T cell responses)
- Immunology (gene expression profiling) in adults only.

- symptomatic malaria parasitaemia (only if participant is symptomatic)
- asymptomatic malaria parasitaemia (retrospectively);
- malaria parasite genotyping (retrospectively);

Safety laboratory results will be reviewed by the study clinician before vaccination and checked by the PI (or designee).

RH5.1/Matrix-M vaccine will then be administered intramuscularly in the left deltoid by trained study staff according to the site vaccination SOP. Each participant will be assessed for at least 1 hour after vaccination to evaluate and treat any acute adverse events. During this time the following will be recorded;

- Post vaccination vital signs
- Post-vaccination solicited signs/symptoms.
- Post-vaccination unsolicited adverse events.
- Post-vaccination SAEs.
- Any concomitant medication.

Day 183; Post-Vaccination with RH5.1/Matrix-M)-Group 1b only

Participants will be seen at CTF where vital signs and temperature monitoring will be performed and recorded in the CRF. AEs will be recorded in the CRF. Blood sample will be collected for the analysis of:

- Immunology (gene expression profiling) in adults only.

Day 184 (-1/+1 days; Post-Vaccination with RH5.1/Matrix-M)

Participants will be seen at CTF where medical history, temperature monitoring and physical examination will be performed and recorded in the CRF. AEs will be recorded in the CRF.

Days 185, 186, 187 and 188 (Post-Vaccination with RH5.1/Matrix-M)

Each participant will be visited at home daily on days 1, 3, 4, 5 and 6 post third vaccination by trained study staff for assessment and recording of any solicited and unsolicited AEs. If necessary, the participant will continue to be seen regularly until the AEs have resolved or stabilised. CHW will be blinded to the group allocation of the subject. During the home visit, the following will be assessed and recorded in the CRF;

- Axillary body temperature.
- Local and general solicited adverse events.
- SAEs experienced by the vaccinees since the last visit.
- Unsolicited adverse events experienced by the vaccinees since the last visit.
- Concomitant medication taken.

Day 189 (-1/+3 days; Post-Vaccination with RH5.1/Matrix-M)

Participants will be seen at CTF where medical history, temperature monitoring and physical examination will be performed and recorded in the CRF. AEs will be recorded in the CRF.

Blood will be collected for analysis of:

- haematology (full blood count);
- biochemistry (including creatinine and ALT);
- symptomatic malaria parasitaemia (only if participant is symptomatic);
- asymptomatic malaria parasitaemia (retrospectively);
- Immunology:

Day 196 (-1/+3 days; Post-Vaccination with RH5.1/Matrix-M)

Participants will be seen at CTF where medical history, temperature monitoring and physical examination will be performed and recorded in the CRF. AEs will be recorded in the CRF.

Blood will be collected for analysis of:

- haematology (full blood count);
- biochemistry (including creatinine and ALT);
- symptomatic malaria parasitaemia (only if participant is symptomatic);
- asymptomatic malaria parasitaemia (retrospectively);
- Immunology (including RH5 specific humoral and T cell responses)

Day 210 (-7/+2 days; Post-Vaccination with RH5.1/Matrix-M)

Participants will be seen at CTF where medical history, temperature monitoring and physical examination will be performed and recorded in the CRF. AEs will be recorded in the CRF.

Blood sample will be collected for the analysis of:

- haematology (full blood count);
- biochemistry (including creatinine and ALT);
- Immunology (including RH5 specific humoral and T cell responses).
- symptomatic malaria parasitaemia (only if participant is symptomatic);
- asymptomatic malaria parasitaemia (retrospectively);

In addition, a bone marrow aspirate sample will be collected (as specified in study specific SOP) in the remaining 4 volunteers from Group 1b who did not undergo bone marrow aspiration on Day 0. Volunteers will have Prothrombin Time/INR and platelets tests within one week before bone marrow aspiration to rule out any bleeding disorder. Bone marrow aspiration will not be conducted in PT/INR are abnormal.

Day 266 (-1/+7 days; Post-Vaccination with RH5.1/Matrix-M)

Participants will be seen at CTF where medical history, temperature monitoring and physical examination will be performed and recorded in the CRF. AEs will be recorded in the CRF.

Blood sample will be collected for the analysis of:

- Symptomatic malaria parasitaemia (only if participant is symptomatic)
- Asymptomatic malaria parasitaemia;
- Immunology(including RH5 specific humoral and T cell responses)

Day 294 (±7 days; Post-Vaccination with RH5.1/Matrix-M)

Medical history, temperature monitoring and physical examination will be performed and recorded in the CRF. AEs will be recorded in the CRF.

Blood sample will be collected for the analysis of:

- haematology (full blood count);
- biochemistry (including creatinine and ALT);
- symptomatic malaria parasitaemia (only if participant is symptomatic);
- asymptomatic malaria parasitaemia;
- malaria parasite genotyping;
- Immunology(including RH5 specific humoral and T cell responses)

Year 1.5 (± 28 days; Post-Vaccination with RH5.1/Matrix-M)

Medical history, temperature monitoring and physical examination will be performed and recorded in the CRF. AEs will be recorded in the CRF.

Blood sample will be collected for the analysis of:

- haematology (full blood count);
- biochemistry (including creatinine and ALT);
- Immunology(including RH5 specific humoral and T cell responses)
- symptomatic malaria parasitaemia (only if participant is symptomatic);
- asymptomatic malaria parasitaemia;

Year 2.5 (± 28 days; Post-Vaccination with RH5.1/Matrix-M)

Medical history, temperature monitoring and physical examination will be performed and recorded in the CRF. AEs will be recorded in the CRF.

Blood sample will be collected for the analysis of:

- haematology (full blood count);
- biochemistry (including creatinine and ALT);
- Immunology:
- symptomatic malaria parasitaemia (only if participant is symptomatic);
- asymptomatic malaria parasitaemia;

#### 11.6. **Unscheduled Visits:**

Participants will be advised to seek first line health care at CTF where they were enrolled during working hours and at BDH outside the working hours. The CTF and BDH will be staffed by clinically-qualified health workers who are on call 24 hours a day, 7 days a week. If needed participants will be referred, and if urgently needed, transported to Muhimbili National Hospital. In the event the unscheduled visit occurs in another health facility then whenever possible, clinical notes related to the management of that event will be retrieved.

Participants will also be informed that community health workers will be readily available to facilitate access to care during the course of the trial. CHW will have access to a study clinician for consultation by mobile phone 24 hours a day and 7 days a week. Reimbursements for transport to CTF or BDH will be provided when participants present for unscheduled visits.

#### 11.7. **Sample Handling**

Samples collected during the study will be used to address the study objectives and obtain a better understanding of vaccine immune responses following immunization with RH5. Samples may be used to make monoclonal antibodies for analysis related to this vaccine. It is possible that these antibodies may be developed into a product in the future. The participant may withdraw permission for future use of specimens at any time. If a participant withdraws his or her permission for future use of specimens, the Investigator or designee will destroy all known remaining specimens and report this destruction to the participant and the EC. This decision will not affect the participant's participation in this protocol or any protocols supported by Ifakara Health Institute.

All samples stored will be labelled with the participant's study identification (ID) number, which cannot identify the study participant directly but is linkable to other research databases (e.g., questionnaires, clinical assessments, logbooks) generated by the main study. The subject identification log linking the study participant ID number to the name of the participant will be maintained with access limited to authorized research team members. In the event of samples being requested in the future, only the PIs or study coordinator(s) will have access to the log linking the study participant to the samples. We will not collect duplicate samples. Only the necessary volume of blood will be collected to address primary, secondary and exploratory objectives of the study.

At the completion of the clinical study protocol and the primary and secondary immunology studies described in this protocol, remaining samples will either be destroyed or stored indefinitely (Section 17.9).

In more detail: for samples held at IHI in Tanzania, samples from participants whose parents/guardians or themselves did not provide permission to store them will be discarded after the primary and secondary analyses described in this protocol have been completed. Samples will also be destroyed if the study participants withdraw their permission to store the samples for future studies during the trial, and investigator or designee will report the destruction to the participant and EC (IHI IRB, OxTREC and NatHREC). If there is no withdrawal all remaining samples will either be destroyed or transferred to IHI biobank and stored indefinitely (only if the permission to store the samples indefinitely has been obtained from the participants) for future use. Any additional immunological analyses on these samples will be limited to malaria vaccine or malaria monoclonal antibody development unless specific permission for additional studies is obtained from the relevant ECs. Transfer to another protocol will require approval from the ECs. In the future, other investigators (both at IHI and outside) may wish to study these samples and/or data. In that case, ECs (IHI IRB and NatHREC) approval must be sought prior to any sharing of samples. Any clinical information shared about the sample with or without participant identifiers would similarly require prior EC approval (IHI IRB and NatHREC).

The samples exported to UK will be stored at the University of Oxford and used for analyses as outlined in the protocol during the study period. However, once the ethical approval on the protocol expires (1 year after the last subject last visit) the samples will be either be returned, destroyed or will be transferred to the Oxford Vaccine Centre (OVC) Biobank (only in the cases where consent to store the samples indefinitely has been provided by the participant). The OVC Biobank acts as a secure and regulatory compliant storage facility. Research scientists including Tanzanian investigators who wants to have access to the stored samples will have to submit a request through OVC Biobank who will review the application to ensure the research is in line with consented use of samples.

Blood samples (serum, plasma, whole blood and PBMC) will be stored at IHI BRTC Laboratory in Kingani, Bagamoyo, Tanzania and some may be shipped to the following collaborative laboratories;

- Jenner Institute laboratories, Centre for Clinical Vaccinology & Tropical Medicine, University of Oxford, Churchill Hospital, Old Road, Oxford, OX3 7LJ, United Kingdom.
- Department of Biochemistry laboratories, University of Oxford, 3 South Parks Road, OX1 3QU
- NIH/NIAID Laboratory of Malaria and Vector Research, Malaria Immunology Section, GIA Reference Centre, 12735 Twinbrook Parkway, Twinbrook III, Room 3W-13, Rockville, MD 20852, USA.
- KEMRI-Wellcome Trust Research Laboratories, P.O. Box 230 80108 Kilifi, Kenya.

At BRTC laboratory, freezer and refrigerator temperatures are recorded twice a day (in the morning and evening), through in-built system and thermometers. The temperature of the freezers will be recorded on temperature logs; in addition, temperature is also continuously recorded by a data logger.

#### 11.8. Laboratory Evaluations

*Safety Blood Assessments*

Blood samples will be collected at specific time-points for haematological and biochemistry tests. The time points and the specific tests are shown in Appendix A. Protocol specific tests will include the following;

- Complete Blood Count:
  Red blood count, haemoglobin, platelets, white blood count with differential (neutrophil, lymphocyte and eosinophil) counts, and Prothrombin time/International normalised ratio (INR) test to assess time for blood to clot.
- Biochemistry parameters:
- *At screening:* ALT and creatinine.
- *During subsequent scheduled visits:* ALT and creatinine.
- Iron Status tests

During unscheduled visits, laboratory parameters to be tested will depend on the relevance as determined by the study clinician.

*Urinalysis*:

Urine sample will be collected at screening for urinalysis.

*Malaria Microscopy:*

The WHO malaria microscopy method will be adopted and used to identify (thick blood smear) and quantify (thin blood smear) malaria parasites in participants. Microscopy will be done at screening (before RH5.1/Matrix-M), and monthly thereafter to monitor concurrent malaria infections during the study, and whenever participant presents with malaria symptoms.

*Serology*

Blood samples for HIV, HBV and HCV serology will be done at the screening visit 1 and all individuals infected with any one of these viruses will be excluded from the study.

*Urine Sample Pregnancy Test*

For female adult participants (18-45 years), a urine pregnancy test will be performed at first screening and before each vaccination. Pregnant women will be excluded from participation. If pregnancy is diagnosed after vaccination, the participant will be excluded from further vaccination but will be followed up for safety until the end of the study. If possible, the outcome of the pregnancy will also be determined, including the health of the new-born.

*Antibodies to blood stage antigens*

At screening blood samples will be collected and ELISA will be conducted to quantify antibodies to schizonts. To determine if volunteers have high or low previous exposure, a pooled population mean anti-schizont OD value from Bagamoyo district will be used. Volunteers with OD values above upper 25% quarrtile of population mean will be classified as having high exposure and included in the trial. Antibodies to other merozoites antigens including but not limited to AMA-1and MSP-1 may be assessed and later correlated to the anti-schizont OD.

##### 11.9. Rationale for blood volume collected

As part of the process to address the research question, blood samples will be collected to assess both the safety and immunogenicity of RH5 candidate malaria vaccine. It is however imperative to prioritise the safety and wellbeing of the study participants during the study as stipulated in the Declaration of Helsinki. In order to minimize risk to the study participants, the amount of blood volume that will be collected has been estimated using existing guidelines which recommend age-specific volume limits based on total blood volume, haemoglobin level and general health status of the donors [96]. The blood volume limits as recommended by WHO range from 1-5% of the total volume within 24 hours and up to 10% of total blood volume over 8 weeks[97]. A mean of 4% of TBV has been selected as the maximum limit in this trial. This is equivalent to around 3.2mL/Kg. A mean weight for each age group was estimated using the maximum and minimum normal weight limit at –2SD of WHO growth charts and a standard 70kg for young adults. Table 3 show the estimated maximum allowable blood volume per visit and the maximum amount of the blood volume that will be collected. The total blood volume that will be collected per participant during the whole study period of 2.5 years for each study group is shown in **Appendix A**.

Table 3: Maximum allowable blood volumes to be collected

| Age Group | Mean weight (Kg) | Maximum allowable blood volume per visit | Maximum blood volume to be collected per visit |
|---|---|---|---|
| Infants (5-17 months) | 6.5 | 21mL | 12 mL |
| Adults (18-45 years) | 70 | 140mL | 104.5 mL |

For infants, a lower volume will be taken to minimize the parents/guardian anxiety. Only health participants will be included in the study. This will ensure all participants have normal haemoglobin levels.

#### 11.10. Research Assays to Address Secondary endpoints

*Humoral and cell medicated Immunity*

The blood volumes for the immunological outcomes and time points at which samples will be collected will depend on the group of the participants as shown in Appendix A.

Serum samples collected will be assessed by ELISA for antibodies to RH5 antigen. Growth inhibition activity (GIA) against vaccine homologous *Pf* line and potentially a panel of other *Pf* isolates and lines to determine antibody function against merozoite invasion of erythrocytes will be performed. Serum isotypes, IgG subclasses and avidity will be assessed by ELISA, and linear epitopes may be mapped by peptide arrays. Single-cell analysis of RH5-specific B cells may be conducted and would allow for production of human monoclonal antibodies to investigate epitope specificities. Surface Plasmon resonance may be used to assess antibody affinity to RH5 and quantify the concentration of RH5 specific IgG.

PBMC will be stored after separation at -70ºC and then transferred into liquid nitrogen vapour phase. Fresh or stored samples will be used to address if T cells are activated by vaccination and show vaccine specific responses by IFN-γ+ ELISPOT.

A number of exploratory assays will also be conducted to further characterize the cellular immune responses following vaccination with RH5.1/Matrix-M. Some of these assay methods are currently being refined and will be described in details in the immunology analysis plan. These assays will be prioritized by the investigators to ensure optimal use of limited sera and cells. Some of these assays will include but are not limited to;

- Anti-RH5 µg/ml: ELISA with CFCA
- Antigen Secreting Cells IgG/ IgM ELISPOTs
- T-follicular helper cells: *ex vivo* and *in vitro* flow cytometry
- IgG+/ IgM+ memory B cell: ELISPOT (indirect)
- IgG+/ IgM+ memory B cell: tetramer-based detection (direct; flow cytometry)
- Bone marrow aspirate: LLPC ELISPOT and flow cytometry

The specific types of cells that the analysis will focus on will be further described in immunology analysis plan. We will also conduct transcriptional profiling of the RH5 specific responses and seek to identify key pathways upregulated in response to vaccination with different dosing regimens targeting RH5.1_FL protein. All analyses will be conducted at BRTC labs or collaborative research laboratory with the exception of GIA, gene expression studies and B cell epitope analyses. These will be conducted at The Jenner Institute laboratory/Department of Biochemistry laboratories in the UK, NIH and collaborating laboratories.

*Investigating tissue-specific responses in vivo is a critical knowledge gap in human immunology.*

Bone marrow is a reservoir for plasma cells and long-lived memory cells, and is the most likely site for immune-priming in malaria. Furthermore, the antibodies that are critical to RH5-mediated protection are produced by long-lived plasma cells, and these cells are only thought to be present in the bone marrow. These antibodies are not produced by circulating B cell populations. The long-lived plasma cells found in the bone marrow are therefore more relevant to the serum anti-RH5 IgG concentrations than RH5-specific plasmablasts detectable in peripheral blood. Characterizing the phenotype and frequency of RH5-specific B cells in the bone marrow would therefore give much better insight into the relevant immune responses. Despite this, vaccination studies very rarely include bone marrow sampling in their protocols so there remains this missing link in immunogenicity analyses.

We therefore plan to isolate bone marrow aspirate at baseline and 4 weeks after the third vaccination with RH5.1/Matrix-M in adult volunteers. Only adult volunteers in Group 1A and 1B will have bone marrow aspirate collected. To ensure each volunteer only provides one bone marrow aspirate sample, two participants from each group will have bone marrow aspirates at baseline, and four volunteers from each group will have bone marrow aspirates at 4 weeks after the last vaccination. We will for the first time in malaria vaccine trials assess the vaccine specific plasma cells, long lived memory cells and how they correlate with vaccine specific B cells and antibody levels.

Diagnostic Laboratory Department of Muhimbili National Hospital has experience in bone marrow procedures (aspirates and trephine biopsies). The hospital receive referred patients from all over Tanzania and conducts 10-20 bone marrow sampling per week. We will work with the specialist from the hospital to collect bone marrow aspirates from the adult volunteers participating in the trial. The procedure is safe and well tolerable and it takes about 10 minutes only. As with any procedure where a sample of tissue is taken, there is a small risk of bleeding from the puncture site and a small risk of infection at the site, but extreme care is taken to keep the procedure sterile in order to minimise this risk. Mild pain/discomfort at the time of and after tissue aspiration is short-lived and easily relieved with paracetamol. The clinical trial facility in Kingani has an ICU which is well equipped to allow this procedure to take place. The department in collaboration with our study clinicians will provide personnel to perform this procedure safely and with minimal side effects.

It is also important to note that the risk of bleeding from a bone marrow procedure is very small in a healthy volunteer and considerably less than that from a liver biopsy. A bone marrow aspirate is also much less invasive than a lymph node biopsy, a procedure which has been done previously in a clinical trial conducted by IHI among HIV infected adults, and approved by IHI IRB and NatHREC.

### 12. INVESTIGATIONAL PRODUCT (IP)

#### 12.1. IP Description

##### The RH5.1 Protein (also see Investigator's Brochure [IB]

RH5.1 was manufactured under Good Manufacturing Practice (GMP) by the Clinical Biomanufacturing Facility (CBF) in Oxford in 2015. The doses to be used in this study are 10 µg and 50 µg. These are nominal doses as the actual dose may vary slightly dependent on dilution, mixing and administration.

The Quality Control Standards and Requirements for the vaccine are described in separate Quality Assurance documents (e.g. certificate of analysis) and the required approvals have been obtained. The vaccines are QP certified, labelled and packed according to applicable regulatory requirements at the CBF.

In summary the RH5.1 protein consists of the entire full-length ectodomain of the RH5 antigen (amino acids E26 – Q526) with the sequence based on the 3D7 clone of *P. falciparum*. The vaccine was produced from a stable *Drosophila* S2 cell line, and also contained an N-terminal 18 αα BiP insect signal peptide (MKLCILLAVVAFVGLSLG) which is cleaved off as the protein is secreted from the cell, and a C-terminal four amino acid (E-P-E-A) "C-tag" used for affinity purification. The cell line system called ExpreS2 was provided by ExpreS2ion Biotechnologies in Denmark.

##### Matrix-M

The Matrix-M adjuvant is manufactured by Novavax AB (Uppsala, Sweden), a subsidiary of Novavax, Inc. The adjuvant contains purified saponin components derived from an extract of the bark of the Quillaja saponaria tree; a phospholipid, egg-derived phosphatidylcholine (PC); and semi-synthetic cholesterol of non-animal origin.

*Matrix-M1 supply*

Matrix-M1 will be supplied to Xerimis by Novovax AB (Uppsala, Sweden), a subsidiary of Novavax, Inc, where the adjuvant is formulated and vialed. All adjuvants will be certified for release by a qualified person (QP) at Xerimis Ltd, Reading and labelled for investigational use only. Matrix M will then be shipped to IHI clinical trial facility by Xerimis Ltd.

*Matrix-M1 formulation and packaging*

Matrix-M1 is formulated at a concentration of 0.375mg/ml in PBS. The drug product is filled into sterile 2mL glass vials. Matrix-M1 (85 parts Matrix A and 15 parts of Matrix C) is obtained by simply mixing Matrix A and C, followed by dilution in PBS, filtration though filter 0.22 µm and filling into vials in a volume of 0.75 mL (nominal 0.6 mL for extraction). Matrix-M1 is a colourless slightly-opalescent non-viscous liquid.

#### 12.2. Storage of IP

Any temperature deviation outside the ranges specified above must be reported to the Sponsor as soon as detected. Following an exposure to such a temperature deviation, vaccines will not be used until Sponsor approval has been given.

The RH5.1 vaccine will be shipped from Oxford on dry ice, and then stored in between –70°C and -90°C in a freezer that will be located in a secure pharmacy until required. Matrix-M adjuvant will be shipped to the CBF by the manufacturer and will be stored between +2 and +8°C in a fridge that is located in a secure pharmacy. The storage conditions will be under the responsibility of the study pharmacist. Vaccine accountability, storage, shipment and handling will be in accordance with relevant local standard operating procedures (SOPs) and forms. All movement of vaccines will be documented in vaccine accountability logs according to local site SOPs.

During vaccinations the vaccine and adjuvants will be removed from the freezer and fridge and they will be formulated and used within 1 hour of thawing. The vaccine and adjuvant will be handled according to the relevant SOPs.

#### 12.3. Packaging and Labelling

The Jenner Institute will supply RH5.1 and Novavax AB will supply Matrix-M adjuvant to the study site. An example of labels for RH5.1 and Matrix-M is shown in Figure 12. This is only an example and general format, but the final labels are subject to change.

| | |
|---|---|
| **CLINICAL TRIAL: VAC080**<br>RH5.1 VACCINE 0.8 mL (nominal)<br>Solution for Injection<br>Vial contains 0.174 mg/mL<br>Vial no: _____ for Intramuscular injection<br>Lot No: 02M15-01 Store at -80°C<br>Volunteer number: _ Expiry date: xxxxxx | **FOR CLINICAL TRIAL USE ONLY**<br>PI: Dr. Ally Olotu<br>Sponsor: University of Oxford<br>CCVTM, Old Road, Oxford OX3, 7LE<br>Tel: 01865 611422<br>Fax: 01865 289694 |
| **CLINICAL TRIAL: VAC080**<br>Matrix-M1 Adjuvant 0.6 mL (nominal)<br>Solution for Injection<br>Vial contains 0.375 mg/mL<br>Vial no: _____ for Intramuscular injection<br>Lot No: M1-108 store at +2-8°C<br>Volunteer number: _ Expiry date: xxxxxx | **FOR CLINICAL TRIAL USE ONLY**<br>PI: Dr. Ally Olotu<br>Sponsor: University of Oxford<br>CCVTM, Old Road, Oxford OX3, 7LE<br>Tel: 01865 611422<br>Fax: 01865 289694 |

*Figure 12: Example of Vaccine labels*

#### 12.4. Accountability of the Investigational Product

At all times the figures on supplied, used and remaining vaccine doses should match. At the end of the study, it must be possible to reconcile delivery records with those of used and unused stocks. An explanation must be given of any discrepancies. The study pharmacist will ensure accurate records are maintained with regard to the date the vaccine is received, manufacture date, lot numbers, quantity received and disposition of the vaccine according to the relevant SOP. The Sponsor (Oxford University) will also maintain a copy of the records held at the site including Study ID numbers, date and time the IP has been administered, lot number and signature of the person administering the IP. All information will be kept in a confidential place by the pharmacist. The clinical team will not have access to the vaccine preparation room during vaccination. All unused vaccines will be returned to Jenner Institute at the end of the trial, or destroyed if requested.

##### 12.5. Administration of the IP

RH5.1/Matrix-M will all be administered intramuscularly into the left deltoid area according to study specific SOP. Each participant will be monitored for one hour (or longer if necessary) after each vaccination. Resuscitation equipment and medication will be available in the clinic site. Staff trained in Advanced Cardiac Life Support and Paediatric Advanced Life Support will be present at all times.

##### 12.6. Concomitant Medication

Collection of concomitant medications will be done at each visit throughout the immunization period until 30 days after the RH5.1/Matrix-M vaccination. Participants will be assessed by the study clinician and medication provided in the event of adverse events. The treatment will follow the Tanzania Ministry of Health, World Health Organization and/or international guidelines. Information on the use of antimalarial drugs will specifically be sought during scheduled clinic visits.

### 13. SAFETY REPORTING

#### 13.1. Definitions

*Adverse Event*

Any untoward medical occurrence in a patient or clinical investigation subject administered a pharmaceutical product and which does not necessarily have a causal relationship with this treatment. An adverse event (AE) can therefore be any unfavourable and unintended sign (including an abnormal laboratory finding), symptom, or disease temporally associated with the use of a medicinal (investigational) product, whether or not related to the medicinal (investigational) product.

Anticipated day-to-day fluctuations of pre-existing conditions that do not represent a clinically significant exacerbation will not be considered adverse events.

Discrete episodes of chronic conditions occurring during a study period will be reported as adverse events in order to assess changes in frequency or severity.

Adverse events will be documented in terms of a medical diagnosis. When this is not possible, the adverse event will be documented in terms of signs and symptoms observed by the Investigator at each study visit.

Pre-existing conditions or signs and/or symptoms (including any which are not recognised at study entry but are recognised during the study period) present in a subject prior to the start of the study will be recorded on the Medical History form within the subject's CRF.

*Adverse Reaction (AR)*

An AR is any untoward or unintended response to an IP. This means that a causal relationship between the IP and an AE is at least a reasonable possibility, i.e., the relationship cannot be ruled out. All cases judged by the reporting medical Investigator as having a reasonable suspected causal relationship to an IP (i.e. possibly, probably or definitely related to an IP) will qualify as adverse reactions.

*Unexpected Adverse Reaction*

An adverse reaction, the nature or severity of which is not consistent with the applicable product information (e.g., IB for an unapproved IP).

*Serious Adverse Event (SAE)*

A serious adverse event (experience) or reaction is any untoward medical occurrence that at any dose:

- results in death,
- is life-threatening, note: the term "life-threatening" in the definition of "serious" refers to an event in which the patient was at risk of death at the time of the event;

it does not refer to an event which hypothetically might have caused death if it were more severe.

- requires inpatient hospitalisation or prolongation of existing hospitalisation,
- results in persistent or significant disability/incapacity, or
- results in a congenital anomaly/birth defect;

*Serious Adverse Reaction (SAR)*

An adverse event (expected or unexpected) that is both serious and, in the opinion of the reporting Investigator or Sponsors, believed to be possibly, probably or definitely due to an IP or any other study treatments, based on the information provided.

*Suspected Unexpected Serious Adverse Reaction (SUSAR)*

A serious adverse reaction, the nature and severity of which is not consistent with the information about the medicinal product in question set out in the IB or Summary of Product Characteristics (SmPC).

Medical and scientific judgment should be exercised in deciding whether expedited reporting is appropriate in other situations, such as important medical events that may not be immediately life-threatening or result in death or hospitalisation but may jeopardise the patient or may require intervention to prevent one of the other outcomes listed in the definition above. These should also usually be considered serious.

##### 13.2. **Procedures for Recording Adverse Events**

*Recording adverse events*

Investigators will evaluate all adverse events observed or reported by the participant or their parents/guardians at each scheduled or unscheduled visit. New adverse events will be recorded in the Adverse Event form within the participant's CRF. Solicited adverse events will be recorded on separate pages of the CRF. The nature of each event, date of onset, outcome, intensity and relationship to vaccination will be established.

Any corrective treatment will be recorded in the CRF as concomitant medication. For solicited adverse events participants or their parents/guardians will be asked direct questions related to the pre-listed symptoms. For unsolicited adverse events, participants or their parents/guardians will be asked an open-ended question such as: "Have you felt different in any way since receiving the vaccine or since the last visit?" The investigator will record only those adverse events having occurred within the time frames defined above.

Adverse events already documented in the CRF, i.e. at a previous assessment and designated as 'ongoing' will be reviewed at subsequent visits, as necessary. If these events have resolved, the documentation in the CRF will be completed, including the date that the adverse event resolved. If an adverse event changes in frequency or intensity during a study period, the record will be updated to reflect the maximum intensity or describe the frequency.

*Solicited Systemic AEs*

For all participants, a prescribed list of systemic AEs will be solicited on the day of vaccination through to 7 days post vaccination. During these periods all participants will be examined for elevated body temperature and allergic reaction (rash, urticaria, pruritus, and/or oedema). Additional AEs occurring in adults (Group 1) will be solicited by history from a list of systemic AEs (nausea, subjective fever, arthralgia, chills, malaise, myalgia, and headache) while the parents/guardians of infants (Group 2) will be asked about observed history of fever, vomiting, diarrhoea, reduced oral intake, and reduced activities (see **Table 4**).

*Solicited Local AEs:*

Pain, limitation of arm movement, pruritus, bruising/ extravasated blood, erythema, swelling, tenderness and induration will be solicited from all participants after each injection. For infants answers to pain, tenderness and pruritus may not be able to be obtained. Local AEs will be solicited on the day of vaccination and 7 days following each vaccination (see **Table 4**).

*Unsolicited AEs*

Unsolicited AEs will be conducted up until day 28 after each vaccination. A syndromic classification will be used for unsolicited AEs, e.g., cough, nasal congestion, sore throat should be combined into upper respiratory tract infection. Thus the term for the unifying diagnosis is recorded as the AE, not each individual sign or symptom whenever applicable.

***Table 4: Solicited Adverse Events***

| | | | | |
|---|---|---|---|---|
| Local Solicited AEs *(at injection site)* | • Pain<br>• Limitation of arm movement | | 0 | No pain |
| | | | 1 | Painful on touch, no restriction on movement of limb (baby cries when limb is touched but moves the limb around without restriction) |
| | | | 2 | Painful when arm is moved (baby cries when limb moved) |
| | | | 3 | Unable to use the limb due to pain |
| | • Pruritus | | 0 | No symptom/sign |
| | | | 1 | Awareness of a symptom, but the symptom is easily tolerated and causes no or minimal interference with usual activity. |
| | | | 2 | Discomfort enough to cause greater than minimal interference with usual activity. |
| | | | 3 | Incapacitating; symptoms causing inability to perform usual activities; requires medical intervention |
| | Infants | • Erythema<br>• Swelling<br>• Induration | 0 | 0 |
| | | | 1 | <20 mm |
| | | | 2 | 20-50 mm |
| | | | 3 | >50 mm and/or necrosis or exfoliative dermatitis |
| | Adults | • Erythema<br>• Swelling<br>• Induration | 0 | 0 |
| | | | 1 | <50 mm |
| | | | 2 | 50 – 100 mm |
| | | | 3 | >100 mm and/or necrosis or exfoliative dermatitis |
| Systemic Solicited AEs | • Fever (objective) | | | |
| | | | 1 | 37.6°C – 38.4°C |
| | | | 2 | 38.5°C – 39°C |
| | | | 3 | >39.0°C |
| | Adults | • Allergic reaction *(rash, urticaria, pruritis, oedema)*<br>• Headache<br>• Nausea<br>• Subjective Fever<br>• Fatigue/Malaise<br>• Chills<br>• Myalgia<br>• Arthralgia | 0 | No symptom/sign |
| | | | 1 | Awareness of a symptom, but the symptom is easily tolerated and causes no or minimal interference with usual activity. |
| | Infants | • Allergic reaction *(rash, urticaria, pruritis, oedema)*<br>• Subjective fever<br>• Vomiting<br>• Diarrhoea<br>• Reduced activities<br>• Reduced oral intake | 2 | Discomfort enough to cause greater than minimal interference with usual activity. |
| | | | 3 | Incapacitating; symptoms causing inability to perform usual activities; requires absenteeism or bed rest |

#### *Follow-up of Adverse Events*

All AEs will be followed until resolution of the signs or symptoms or laboratory changes occurs, or until a non-study related causality is assigned.

At the study end, if participants have moderate or severe on-going adverse events not considered to be related to the study vaccine, they will be advised to consult a government health facility. If considered to be related to the study vaccine, a follow-up visit will be arranged to manage the problem and to determine the severity and duration of the event. If appropriate, specialist review will be arranged by Investigators.

Any serious adverse event possibly related to the vaccine and occurring after termination of the clinical trial should be reported by the Investigator according to the procedure described below.

*Grading the severity of adverse events*

For adverse events other than local swelling, redness/discoloration and pain/limitation of limb movement, for which the severity scales are detailed above, AEs will be graded according to the relevant grading tables[98,99]. These internationally recognized grading tables classify adverse events into one of four grades, ranging from mild to potentially life-threatening. The grading tables have indications for each of over sixty clinical parameters and forty laboratory parameters for grading adult and paediatric AEs. The table also includes general guidelines for estimating the grade of parameters not explicitly listed. Each grade is described broadly below:

- Grade 1 (mild): awareness of a symptom, but the symptom is easily tolerated and causes no or minimal interference with usual activity.
- Grade 2 (moderate): discomfort enough to cause greater than minimal interference with usual activity.
- Grade 3 (severe): incapacitating; symptoms causing inability to perform usual activities; requires absenteeism or bed rest.
- Grade 4 (potentially life-threatening): symptoms causing inability to perform basic self-care functions OR medical or operative intervention is indicated to prevent permanent impairment, persistent disability or death.

Laboratory tests will also be graded based on the relevant toxicity grading scales relevant to healthy HIV negative volunteers with modification to suit local or regional normal values. In addition to the referenced toxicity grading scales above, any other relevant toxicity grading scale may be used to supplement and accomplish proper assessment. Toxicity grades and reference scales which will be used to grade severity of laboratory solicited AEs for this trial will be described in a relevant study SOPs. The abnormal values will be categorized as either clinically significant or non-clinically significant based on the study physician's medical judgement. Abnormal laboratory assessments that are judged by the Investigator to be serious will be recorded as SAEs.

##### 13.3. Causality

The causal relationship between the AE and the product will be evaluated by the PI. This interpretation will be based on the type of event, the relationship of the event to the time of vaccine administration, and the known biology of vaccine therapy. The following are guidelines for assessing the causal relationship:

No relationship:
- No temporal relationship to study product; and
- Alternate aetiology (clinical state, environmental or other interventions); and
- Does not follow known pattern of response to study product

Unlikely relationship
- Unlikely temporal relationship to study product; and
- Alternate aetiology likely (clinical state, environmental or other interventions); and
- Does not follow known typical or plausible pattern of response to study product

Possible relationship:
- Reasonable temporal relationship to study product; or
- Event not readily produced by clinical state, environmental or other interventions; or
- Similar pattern of response to that seen with other vaccines

Probable relationship:
- Reasonable temporal relationship to study product; and
- Event not readily produced by clinical state, environment, or other interventions or
- Known pattern of response seen with other vaccines

Definite relationship:
- Reasonable temporal relationship to study product; and
- Event not readily produced by clinical state, environment, or other interventions; and
- Known pattern of response seen with other vaccines

When a regulatory authority requests distinct classification of AEs into either related or unrelated, without intermediate categories, only “not related” will be regarded as “unrelated” and “unlikely related”, “possibly related,” “probably related,” and “definitely related” will be combined as “related”.

##### 13.4. Reporting Procedures for Serious Adverse Events

In order to comply with current regulations on serious adverse event reporting to regulatory authorities, the event will be documented accurately and notification deadlines respected. SAEs will be reported on the standard SAE forms to members of the study team immediately when the investigators become aware of their occurrence. Copies of all reports will be forwarded by email for review to the Chief Investigator (as the Sponsor's representative) within 24 hours of the Investigator being aware of the suspected SAE. The ISMs forming DSMB will be notified immediately by the PI if SAEs are deemed possibly, probably or definitely related to study interventions; in such a situation, PI will notify the DSMB immediately within 24 hours of the PI being aware of their occurrence.

SAEs will not normally be reported immediately to the OxTREC unless there is a clinically important increase in occurrence rate, an unexpected outcome, or a new event that is likely to affect safety of trial volunteers, at the discretion of the Chief Investigator and/or DSMB. Sponsor will notify The Oxford Tropical Research Ethics Committee (OxTREC) of all SAE that are deemed possibly, probably or definitely related to study interventions and SUSARs within specified time-frame as required by the regulations.

All SAEs will be summarized and reported to IHI IRB and The National Health Research Ethics Sub-Committee (NatHREC) in the required specified time-frames. The PI, on behalf of the Sponsor, will also report all SAEs to the Tanzanian Food and Drug Administration within specified time-frame as required by the regulations.

Additional or follow-up information (outcome, precise description of medical history, results of the investigation, copy of hospitalisation report, etc.) relating to the initial SAE report will also be reported to the Sponsor and DSMB within 24 hours of receipt of such information.

*Reporting Procedures for SUSARS*

The Chief Investigator (on behalf of The Sponsor) will report all SUSARs to the ethical committee(s) and Tanzanian regulatory authorities within required timelines. The Chief Investigator will also inform all Investigators concerned of relevant information about SUSARs that could adversely affect the safety of participants.

All SUSARs and deaths occurring during the study will be reported to the Sponsor. For all deaths, available autopsy reports and relevant medical reports will be made available for reporting to the relevant authorities.

##### 13.5. Data Safety Monitoring Board (DSMB)

For this study, an independent DSMB will be appointed by the UOXF (Sponsor) to follow up the safety of participants as described above and in section 9.7. The DSMB will be composed of 3 members one of whom one will be the Local Safety Monitor (LSM). A local Safety Monitor will be a qualified Tanzanian Paediatrician. All the members will be experienced clinicians qualified to evaluate safety data from clinical studies. The DSMB will be responsible for reviewing the safety results during age de-escalation/dose escalation and to provide clearance to progress to the next vaccinations They will also take part in the assessment of adverse events (if requested) and any recommendation regarding halting further vaccination. The DSMB will liaise closely with the PI throughout the course, mutually relaying relevant safety information. Safety monitoring charter will be developed that will state their responsibility and provide guidance on the procedures for reviewing the safety data.

In addition to the scheduled reviews, DSMB may be contacted for advice and independent review by the Sponsor, CI or PI, as in the following situations:

- AEs in the sentinel participants trigger criteria for an ad hoc DSMB review
- Following any SAE deemed to be possibly, probably, or definitely related to the study vaccine.
- Any other situation where the PI, CI or Sponsor thinks independent advice or review is important.

DSMB will support the review of safety during scheduled and ad hoc reviews. Only SAE deemed possibly, probably or definitely related to study interventions will be reported to the DSMB by the PI. If necessary, Sponsor may notify DSMB of unrelated SAE in case they need their opinion of the SAE. If required, the DSMB may provide a separate written report to document his/her assessment of the SAE or other concerning AE. The occurrence of a SAE possibly related to RH5 will lead to suspension of the trial pending further review of the event by the DSMB, as well as the IHI IRB, Oxford Tropical Research Ethics Committee (OxTREC), National Health Research Ethics Sub-Committee (NatHREC) and TFDA.

### 14. STATISTICAL ANALYSIS CONSIDERATION

Data entry will be performed on site at the IHI clinical trial facility. All final analyses will be performed by the PI with the support from the Sponsor and IHI statisticians.

#### 14.1. Safety Cohort

The 'Safety Cohort' will consist of all participants who have received at least single dose Of RH5.1/Matrix-M for whom any data on safety are available. The presentation of safety data will explore separately the adverse experiences among participants who have received less than three doses of RH5.1/Matrix-M.

#### 14.2. Immunogenicity Cohort

The 'Immunogenicity Cohort' will include all evaluable participants (i.e., those meeting all eligibility criteria, and who have received at least a single dose of RH5.1/Matrix-M vaccination for whom data concerning immunogenicity endpoint measures are available). This will include participants for whom assay results are available for anti-RH5 antibodies after vaccination.

#### 14.3. Sample Size Considerations

The trial is designed to only detect very large differences in the incidence of local and generalized adverse events between the vaccination groups. This is done to balance the chance to detect any possible untoward reactions against the desire to limit the number of volunteers involved for safety purposes. The sample size of 6-12 per group is reasonable given that early phase Ib trials use between 6 and 30 participants per group.

#### 14.4. Final Analysis Plan

A final research analysis plan will be prepared by investigator in advance to guide the statistical analysis and reporting of the study. The analysis plan will be agreed upon by the PI and Sponsor prior to locking of the database for the final analysis. The analysis plan will provide information of the populations that will be analysed and the safety and immunogenicity parameters that will be evaluated as well as specific statistical methods that will be applied. The plan will be applied to the data and sample collected until 2.5 years post first vaccination for each for delayed dosing regimen and 2years post first vaccination for standard dosing regimen.

##### 14.4.1. Analysis of Demographics

Demographic characteristics (age, gender) and baseline covariates of each vaccination group and entire study cohort will be tabulated. The mean age (plus range and standard deviation) and gender proportion of the enrolled participants, as a whole, and per vaccination groups, will be calculated.

##### 14.4.2. Analysis of Safety

Data analysis will consist primarily of descriptive summaries for vaccination groups. The overall percentage of volunteers with at least one local adverse event (solicited or unsolicited) and the percentage with at least one general adverse event (solicited and unsolicited) during the surveillance period after each vaccination will be tabulated. Comparisons between study groups will be made based on a two-sided Fisher's Exact Tests.

##### 14.4.3. Analysis of Clinical Laboratory Parameters

Haematological and biochemical laboratory parameters will be measured at specific time-points. Clinically relevant abnormal values will be tabulated and a trend analysis could be performed if deemed necessary. Where appropriate highly skewed data will be log-transformed and presented as geometric means with 95% confidence intervals.

##### 14.4.4. Analysis of immunological endpoints

The secondary analysis will be based on the immunogenicity cohort. The primary immunogenicity endpoint for humoral immune responses will be the concentration of anti-RH5.1 serum antibodies and their percentage GIA *in vitro* using purified IgG. For cellular immune responses frequency of cells specific to RH5.1 will be identified as T-cells expressing cytokines (e.g. IFN-γ) upon *in vitro* stimulation with RH5.1 antigen and/or RH5-specific ASC or memory B cell responses.

Descriptive summaries and plot distributions of cellular and humoral immune responses over time for each participant and vaccination group will be presented. The results will be presented both as raw data and as log-transformed data. Between and within groups comparisons will be performed by non-parametric Wilcoxon Rank Sum tests or Kruskal–Wallis test and Wilcoxon Signed Rank test respectively.

### 15. DATA HANDLING AND RECORD KEEPING

#### 15.1. Data handling and record keeping

The PI will be the data manager with responsibility for delegating the receiving, entering, cleaning, querying, analysing and storing all data that accrues from the study. Trained study staff will enter the data into the participants' CRFs, which will be in a paper format. These data will include clinical and laboratory safety data. All source documents and laboratory reports will be reviewed by the clinical team and data entry staff, who will ensure that they are accurate and complete. Adverse events must be graded, assessed for severity and causality, and reviewed by the PI or designee.

The Investigators will maintain appropriate medical and research records for this trial in compliance with ICH E6 GCP and regulatory and institutional requirements for the protection of confidentiality of participants. The Investigators will permit authorized representatives of the Sponsor, regulatory agencies and the monitors to examine clinical records for the purposes of quality assurance reviews, audits and evaluation of the study safety and progress.

Data and samples collected will be provided to the Sponsor to allow study related documentation and immunological analyses where these analyses cannot be done in Tanzania. Data and sample shipment agreements will be established between the Sponsor and the IHI before samples and data are transferred to the Sponsor and collaborators.

Study documents will be retained for a minimum of 20 years after the end of the clinical trial. These documents will be retained for a longer period, however, if required by local regulations. No records will be destroyed without the written consent of Sponsor and it is the responsibility of Sponsor to inform the investigator when these documents no longer need to be retained.

#### 15.2. Access to Data

Direct access will be granted to authorised representatives from the Sponsor, host institution and the ethical and regulatory authorities to permit trial-related monitoring, audits and inspections and evaluation of the study safety, progress, and data validity. All documents will be stored safely in a locked filing cabinet where only study Investigators will have access to the keys. Data and sample shipment agreements will be established between the Sponsor and the IHI before samples and data are transferred to the Sponsor. Only anonymized study data including safety data will be shared with the third parties including the sponsors.

#### 15.3. Source document and Case Report Form

All protocol-required information will be collected in CRFs designed by the investigator. All source documents will be filed in the CRF. Source documents are original documents, data, and records from which the volunteer's CRF data are obtained. For this study these will include, but are not limited to; volunteer consent form, blood results, clinician's medical notes, laboratory records, and relevant correspondences. In the majority of cases, CRF entries will be considered source data as the CRF is the site of the original recording (i.e. there is no other written or electronic record of data). In this study collected information in the CRF will include, but is not limited to medical history, medication records, vital signs, physical examination records, urine assessments, blood, urine analysis results, adverse event data and details of vaccinations. If participants fell ill and receive medical treatment, medical notes and investigational results will also be considered as source documents. All source data and participant CRFs will be stored in a secure place accessible to only authorized study staff.

Data entry into the 21 CFR Part 11-compliant Electronic Data Capture system will be done by study staff. The data system will include password protection and internal quality checks, such as automatic range checks, to identify data that appear inconsistent, incomplete, or inaccurate. Clinical data will be entered directly from the source documents. Immunology data will be generated from analyses of blood samples and will be stored in Excel format for sharing and archiving, and analysed using statistical packages (Prism, and/or STATA). The name and any other identifying detail will NOT be included in any trial data electronic file. On all trial-specific documents, other than the signed consent, the participant will be referred to by the trial Study ID number, not by name.

### 16. QUALITY ASSURANCE PROCEDURES

The trial will be conducted in accordance with the current approved protocol, GCP, relevant regulations and standard operating procedures.

#### 16.1. Monitoring

Clinical trial monitoring will be conducted by monitors from KEMRI-Wellcome Trust Research Program Clinical Trial Facility in Kenya and IHI QA team to ensure that GCP standards and regulatory guidelines are being followed. This arrangement will ensure impartiality during monitoring and increase the credibility of the process. The monitoring reports will be sent to Sponsor (University of Oxford) and local PI. Pre-trial monitoring visits will be made to the site, including the clinical laboratory. All records will be made available to monitors, including regulatory files, CRFs and other source documents, QA/QC documentation, SOPs, etc.

A detailed monitoring plan, subject to approval by the sponsor, will be developed by KEMRI-Wellcome Trust Research Program Clinical Trial Facility and IHI QA team. The monitoring plan will include the number of participant charts to be reviewed, which/what proportion of data fields and what will be monitored, and who will be responsible for conducting the monitoring visits, and who will be responsible for ensuring that monitoring findings are addressed.

#### 16.2. Investigator procedures

Approved site-specific SOPs will be used at all clinical and laboratory sites.

#### 16.3. Modification to protocol

No amendments to this protocol will be made without consultation with, and agreement of, the Sponsor. Any amendments to the trial that appear necessary during the course of the trial must be discussed by the Investigator and Sponsor concurrently. If agreement is reached concerning the need for an amendment, it will be produced in writing by the PI and will be made a formal part of the protocol following ethical and regulatory approval.

An administrative change to the protocol is one that modifies administrative and logistical aspects of a protocol but does not affect the subjects' safety, the objectives of the trial and its progress. An administrative change does not require EC or regulatory approval.

The Investigator is responsible for ensuring that changes to an approved trial, during the period for which regulatory and EC approval has already been given, are not initiated without regulatory and EC review and approval except to eliminate apparent immediate hazards to the subject.

#### 16.4. Protocol deviation

A protocol deviation is any noncompliance with the clinical trial protocol, GCP, or SOP requirements. The noncompliance may be either on the part of the participant, the investigator, or the study site staff. As a result of deviations, corrective actions are to be developed by the site and implemented promptly.

It is the responsibility of the site to use continuous vigilance to identify and report deviations within 5 working days of identification of the protocol deviation, or within 5 working days of the planned protocol-required activity. All deviations must be promptly reported to the sponsor.

Any deviations that impact subject safety, or that alter the risk: benefit analysis or the scientific integrity of the study is regarded as major deviation and will be reported to the Sponsor within 24 hours of the PI or study personnel becoming aware of the deviation. Major deviation will be reported to the Tanzanian regulatory authorities by the PI within 7 days of the Sponsor becoming aware of the deviation.

All deviations from the protocol must be addressed in study participant source documents. A completed copy of the Protocol Deviation Form must be maintained in the trial master file, as well as in the participant's source document. Protocol deviations will be reported to the EC's per their guidelines.

#### 16.5. **Audit & inspection**

The QA designated site staff will conduct internal audits to check that the trial is being conducted, data recorded, analysed and accurately reported according to the protocol, sponsor's SOPs and in compliance with ICH GCP. The audits will also include laboratory activities according to an agreed audit schedule.

The Sponsor may carry out audit to ensure compliance with the protocol, GCP and appropriate regulations. GCP inspections may also be undertaken by the regulatory authority to ensure compliance with protocol and national regulations. The sponsor will assist in any inspections.

#### 16.6. **Trial Progress**

The progress of the trial will be overseen by the Principal Investigator

### 17. ETHICAL AND REGULATORY CONSIDERATIONS

#### 17.1. Declaration of Helsinki

The Investigator will ensure that this trial is conducted in accordance with the principles of the Declaration of Helsinki 2008.

#### 17.2. Guidelines for Good Clinical Practice

The Investigator will ensure that this trial is conducted in accordance with Medicine for Human use (clinical trials) Regulations 2004 and its amendments and with the ICH guidelines for GCP (CPMP/ICH/135/95) July 1996. The trial will also comply with the European Communities (Clinical Trials on Medicinal Products for Human Use) Regulations, 2004 [S.I. 190 of 2004].

#### 17.3. Approvals

The protocol, informed consent form, participant information sheet and any proposed advertising material will be submitted to IHI IRB, National Health Research Ethics Sub-Committee (NatHREC) and Oxford Tropical Research Ethics Committee (OxTREC) for written approval. In addition, approval from Tanzania Medicines and Medical Devices Authority (TMDA), a regulatory authority, will be sought before the study starts. No data collection will start without approval from ethics committees and regulatory authority.

The Investigator will submit and, where necessary, obtain approval from the above parties for all substantial amendments to the original approved documents.

#### 17.4. Reporting

The PI shall submit once a year throughout the clinical trial, or on request, an Annual Progress Report to the IHI IRB, National Health Research Ethics Sub-Committee (NatHREC), Oxford Tropical Research Ethics Committee (OxTREC) and Sponsor. In addition, an End of Trial notification and final report will be submitted to the IHI IRB, National Health Research Ethics Sub-Committee (NatHREC), Oxford Tropical Research Ethics Committee (OxTREC) and Sponsor.

#### 17.5. Participant Confidentiality

The trial staff will ensure that the participants' anonymity is maintained. The participants will be identified only by a participant study ID on all trial documents and any electronic database, with the exception of the CRF, where participant initials may be added. All documents will be stored securely and only accessible by trial staff and authorised personnel, including representatives of the REC and regulatory authorities. The trial will comply with the Data Protection Act, 1998, which requires data to be anonymised as soon as it is practical to do so.

Photographs taken of vaccination sites (if required, with the volunteer's written, informed consent) will not include the volunteer's face and will be identified by the date, trial code and subject's unique identifier. Once developed, photographs will be stored as confidential records, as above. This material may be shown to other professional staff, used for educational purposes, or included in a scientific publication.

#### 17.6. Potential Benefits to participants

Participants will benefit from receiving a free consultation and information about their general health status at screening. This information will help participants get medical attention as soon as possible to avoid potential complications. During the trial, all participants will receive information about their health status. Medical costs for acute illnesses whether or not related to the IP or study procedure during the study period will be covered by the study to the limit of the allocated funds. When experiencing illness, enrolled participants will receive care at the clinical trial facility during working hours and at Bagamoyo District Hospital at night or during holidays. If the situation arises in which medical care for a complication unrelated to the study procedure/product exceeds the limits of the budget allocated for clinical care, the participant or guardian will be responsible for funding this care through the normal channels provided by the Ministry of Health and Social Welfare of United Republic of Tanzania.

#### 17.7. Potential Risks and Burden to Participants

*Vaccination:*

Potential expected risks from vaccination, which include local and systemic reactions are described in section 13. It is important to note that RH5.1/Matrix-M has not previously been administered to infants but have been safely administered in UK adults. RH5 expressed by viral vectors ChAd63 and MVA have been safety administered in infants and young children in Africa [100], and recently including Tanzania. Therefore, although the AE profile can be estimated from previous vaccination with viral vectors, the reactogenicity may vary from that seen previously with ChAd63 and MVA. For this reason, vaccinees will be enrolled in a staggered format to allow early identification of any concerning reactogenicity before the majority of individuals have been vaccinated. Any vaccine can cause allergic reactions including anaphylaxis. Therefore, vaccination will take place in the presence Advanced Life Support trained physicians and where equipment and drugs for managing anaphylaxis and any other severe adverse reaction are available.

*Phlebotomy:*

It is expected that participants will experience pain of a needle during vaccination and blood drawing at different time-points. The maximum volume of blood drawn over the study period will be based on age and weight and is not expected to compromise the health of participants. There may be minor bruising, local tenderness or pre-syncopal symptoms associated with venepuncture.

##### 17.8. Incentives

The study will cover all the costs related to participant's participation in the trial including clinic visits or hospitalization. A reasonable time reimbursement of 15,000 Tanzanian shillings will be provided during scheduled visits. In addition, participants will receive a meal during the scheduled visits. In case of expected or unexpected medical complications, medical treatment at CTF and BDH will be available at no cost to the participant. Participants who come for unscheduled visits will be reimbursed for their transport fare. In the event it is required, additional insurance coverage for the costs of medical treatment for acute illnesses or adverse events related to investigational product will be covered by the local clinical trial insurance and liability insurance held by the Oxford University.

##### 17.9. Future use of stored samples

If residual sera and cells are available following the serological and CMI assays described in this protocol, additional immunological and *in vitro* studies related to malaria vaccine development may be performed on those samples for which permission was expressly granted for storing the samples for future studies at the time of informed consent. This may include but not restricted to malaria exposure characterization, age-related immunology, and RH5 epitope mapping. These assays may include ability of participant sera to interfere with *in vitro* parasite growth, volunteer's factors that can influence quantity and quality the RH5 specific immune response (such as iron status) and specificity of RH5 specific immune response. Additional research questions to be asked for cells include antigen-specific cytokine induction as measured by ELISPOT, flow cytometry, or both. Samples from participants whose parents/guardians or themselves did not provide permission to store samples will be discarded after the analyses described in the current protocol have been completed. Study participants will have the right to withdraw their permission for further use of their samples at any time during and after the study.

The samples that are sent to UK will be stored at the Jenner Institute/Department of Biochemistry at the University of Oxford, and used for analyses as outlined in the protocol. At the end of the study (defined as one year after the last subject last visit), samples will be either returned, destroyed, or transferred to the Oxford Vaccine Centre (OVC) Biobank if the consent to store the sample indefinitely has been granted by the participant. Additional analyses related to malaria vaccine development that the volunteers have consented to, can be done on these samples after requests from the Biobank who will be the custodian of the samples while in the UK. Tanzanian PI and the UK investigators will have access to these samples.

Maximizing the research use of collected samples from participants to benefit science and society is an important ethical consideration. It is important to make efficient use of participant samples in an ethical manner with respect and transparency rather than collecting new samples [101-103].

Development of a malaria vaccine is an evolving science and it is expected that the samples collected from this trial and stored for future use will be used to analyse the immune mechanisms of RH5 and other candidate malaria vaccines with potential benefit to science and global society. Development of some advanced candidate malaria vaccines such as RTS,S and ME-TRAP has been ongoing for over 40 years and some of the important insights on these antigens were made from the stored samples collected from different clinical trials and immune-epidemiological studies through collaborative research[32,104,105].

### 18. FINANCE AND INSURANCE

#### 18.1. Funding

The study will be funded primarily by a grant (RIA2016V-1649) from the European and Developing Countries Clinical Trials Partnership (EDCTP).

#### 18.2. Insurance

*Negligent Harm:* Indemnity and/or compensation for negligent harm arising specifically from an accidental injury for which Ifakara Health Institute is legally liable will be covered by the IHI.

*Non-Negligent Harm:* Indemnity and/or compensation for harm arising specifically from an accidental injury, and occurring as a consequence of the Research Subjects' participation in the trial for which the University is the Research Sponsor will be covered by the Ifakara Health Institute and University of Oxford (the University has a specialist insurance policy in place with Newline Underwriting Management Ltd, at Lloyd’s of London, UK). In addition, compensation for injury will be guided by the respective insurance policies that adhere to the national guidelines provided by the TMDA. Clinical Insurance from Strategy Company will be purchased by the study team at IHI to cover study participants.

#### 19. DISSEMINATION AND PUBLICATION POLICY

Study outcomes will be shared with study participants after the final safety report is completed. In addition, local authorities will be informed of the end of the study and the summary of the outcomes. Final safety report will be submitted to all ethics committees and regulatory authority.

The Final Safety Report will be prepared by the PI. The protocol and data derived from the trial are the shared property of the IHI, Oxford University and Novavax. Any publication or presentation, abstracts and press releases related to the trial must be approved by all parties' representatives before submission of the manuscript. Investigator will also seek permission from NatHREC before publishing any manuscript. After publication of the results of the trial, any participating partner may publish or otherwise use its own data provided that any publication of data from the trial gives recognition to the trial group. Either partner must have the opportunity to review the proposed abstract, manuscript or presentation before submission for publication/presentation. A request for delay in publication shall only be allowed to enable the requesting party to secure its proprietary and confidential information contained in the draft publication and/or to prevent the loss of patenting opportunities. Such delay shall not be more than 3 months starting from the day of submission for approval. Any information identified as confidential must be deleted prior to submission. The authorized persons as an author of the publication(s) are those who have contributed to the protocol and/or to the analysis of the data, drafting and reviewing the communication. According to the main topic of the publication, the first author will be the greatest contributing Investigator. Data from the study may also be used as part of a thesis for a PhD, MD or Masters. Publications arising from this study will be made open access.

### 20. REFERENCES

1. World Health Organization (2015) World Malaria Report 2015. World Health Organization. 280 p.
2. Snow RW, Guerra CA, Noor AM, Myint HY, Hay SI (2005) The global distribution of clinical episodes of Plasmodium falciparum malaria. Nature 434: 214-217.
3. Birbeck GL, Molyneux ME, Kaplan PW, Seydel KB, Chimalizeni YF, et al. (2010) Blantyre Malaria Project Epilepsy Study (BMPES) of neurological outcomes in retinopathy-positive paediatric cerebral malaria survivors: a prospective cohort study. Lancet Neurol 9: 1173-1181.
4. Carter JA, Neville BG, White S, Ross AJ, Otieno G, et al. (2004) Increased prevalence of epilepsy associated with severe falciparum malaria in children. Epilepsia 45: 978-981.
5. Idro R, Carter JA, Fegan G, Neville BG, Newton CR (2006) Risk factors for persisting neurological and cognitive impairments following cerebral malaria. Arch Dis Child 91: 142-148.
6. Chuma JM, Thiede M, Molyneux CS (2006) Rethinking the economic costs of malaria at the household level: evidence from applying a new analytical framework in rural Kenya. Malar J 5: 76.
7. Orem JN, Kirigia JM, Azairwe R, Kasirye I, Walker O (2012) Impact of malaria morbidity on gross domestic product in Uganda. Int Arch Med 5: 12.
8. Breman JG, Egan A, Keusch GT (2001) The intolerable burden of malaria: a new look at the numbers. Am J Trop Med Hyg 64: iv-vii.
9. PROGRAMME NMC (2014) National Malaria Strategic Plan 2014–2020: Abridged Version. In: Programme NMC, editor. Dar es salaam The Government of Tanzania.
10. Farnert A, Yman V, Homann MV, Wandell G, Mhoja L, et al. (2014) Epidemiology of malaria in a village in the Rufiji River Delta, Tanzania: declining transmission over 25 years revealed by different parasitological metrics. Malar J 13: 459.
11. Aregawi MW, Ali AS, Al-mafazy AW, Molteni F, Katikiti S, et al. (2011) Reductions in malaria and anaemia case and death burden at hospitals following scale-up of malaria control in Zanzibar, 1999-2008. Malar J 10: 46.
12. Ceesay SJ, Casals-Pascual C, Nwakanma DC, Walther M, Gomez-Escobar N, et al. (2010) Continued decline of malaria in The Gambia with implications for elimination. PLoS One 5: e12242.
13. Okiro EA, Hay SI, Gikandi PW, Sharif SK, Noor AM, et al. (2007) The decline in paediatric malaria admissions on the coast of Kenya. Malar J 6: 151.
14. Assele V, Ndoh GE, Nkoghe D, Fandeur T (2015) No evidence of decline in malaria burden from 2006 to 2013 in a rural Province of Gabon: implications for public health policy. BMC Public Health 15: 81.
15. Okiro EA, Bitira D, Mbabazi G, Mpimbaza A, Alegana VA, et al. (2011) Increasing malaria hospital admissions in Uganda between 1999 and 2009. BMC Med 9: 37.
16. Smith DL, McKenzie FE, Snow RW, Hay SI (2007) Revisiting the basic reproductive number for malaria and its implications for malaria control. PLoS Biol 5: e42.
17. Robertson RL, Foster SO, Hull HF, Williams PJ (1985) Cost-effectiveness of immunization in The Gambia. J Trop Med Hyg 88: 343-351.
18. Holden JD (1987) Benefits and risks of childhood immunisations in developing countries. Br Med J (Clin Res Ed) 294: 1329-1331.
19. Fenner F (1988) Smallpox and its eradication: World Health Organization.
20. de Quadros CA, Hersh BS, Nogueira AC, Carrasco PA, da Silveira CM (1998) Measles eradication: experience in the Americas. Bull World Health Organ 76 Suppl 2: 47-52.
21. Biellik R, Madema S, Taole A, Kutsulukuta A, Allies E, et al. (2002) First 5 years of measles elimination in southern Africa: 1996-2000. Lancet 359: 1564-1568.
22. Rashid H, Khandaker G, Booy R (2012) Vaccination and herd immunity: what more do we know? Curr Opin Infect Dis 25: 243-249.
23. Kim TH, Johnstone J, Loeb M (2011) Vaccine herd effect. Scand J Infect Dis 43: 683-689.
24. Lopez MC, Silva Y, Thomas MC, Garcia A, Faus MJ, et al. (1994) Characterization of SPf(66)n: a chimeric molecule used as a malaria vaccine. Vaccine 12: 585-591.

25. Alonso PL, Smith T, Schellenberg JR, Masanja H, Mwankusye S, et al. (1994) Randomised trial of efficacy of SPf66 vaccine against Plasmodium falciparum malaria in children in southern Tanzania. Lancet 344: 1175-1181.
26. Stoute JA, Slaoui M, Heppner DG, Momin P, Kester KE, et al. (1997) A preliminary evaluation of a recombinant circumsporozoite protein vaccine against Plasmodium falciparum malaria. RTS,S Malaria Vaccine Evaluation Group. N Engl J Med 336: 86-91.
27. Kester KE, McKinney DA, Tornieporth N, Ockenhouse CF, Heppner DG, et al. (2001) Efficacy of recombinant circumsporozoite protein vaccine regimens against experimental Plasmodium falciparum malaria. J Infect Dis 183: 640-647.
28. Thera MA, Doumbo OK, Coulibaly D, Laurens MB, Ouattara A, et al. (2011) A field trial to assess a blood-stage malaria vaccine. N Engl J Med 365: 1004-1013.
29. Ogutu BR, Apollo OJ, McKinney D, Okoth W, Siangla J, et al. (2009) Blood stage malaria vaccine eliciting high antigen-specific antibody concentrations confers no protection to young children in Western Kenya. PLoS One 4: e4708.
30. Seder RA, Chang LJ, Enama ME, Zephir KL, Sarwar UN, et al. (2013) Protection against malaria by intravenous immunization with a nonreplicating sporozoite vaccine. Science 341: 1359-1365.
31. Doolan DL, Hoffman SL (2001) DNA-based vaccines against malaria: status and promise of the Multi-Stage Malaria DNA Vaccine Operation. Int J Parasitol 31: 753-762.
32. Hill AV, Reyes-Sandoval A, O'Hara G, Ewer K, Lawrie A, et al. (2010) Prime-boost vectored malaria vaccines: progress and prospects. Hum Vaccin 6: 78-83.
33. Rts SCTP (2015) Efficacy and safety of RTS,S/AS01 malaria vaccine with or without a booster dose in infants and children in Africa: final results of a phase 3, individually randomised, controlled trial. Lancet 386: 31-45.
34. Olotu A, Fegan G, Wambua J, Nyangweso G, Awuondo KO, et al. (2013) Four-year efficacy of RTS,S/AS01E and its interaction with malaria exposure. N Engl J Med 368: 1111-1120.
35. Olotu A, Fegan G, Wambua J, Nyangweso G, Leach A, et al. (2016) Seven-Year Efficacy of RTS,S/AS01 Malaria Vaccine among Young African Children. N Engl J Med 374: 2519-2529.
36. Gilbert SC, Plebanski M, Harris SJ, Allsopp CE, Thomas R, et al. (1997) A protein particle vaccine containing multiple malaria epitopes. Nat Biotechnol 15: 1280-1284.
37. Webster DP, Dunachie S, Vuola JM, Berthoud T, Keating S, et al. (2005) Enhanced T cell-mediated protection against malaria in human challenges by using the recombinant poxviruses FP9 and modified vaccinia virus Ankara. Proc Natl Acad Sci U S A 102: 4836-4841.
38. Bejon P, Mwacharo J, Kai O, Mwangi T, Milligan P, et al. (2006) A phase 2b randomised trial of the candidate malaria vaccines FP9 ME-TRAP and MVA ME-TRAP among children in Kenya. PLoS Clin Trials 1: e29.
39. Ogwang C, Afolabi M, Kimani D, Jagne YJ, Sheehy SH, et al. (2013) Safety and immunogenicity of heterologous prime-boost immunisation with Plasmodium falciparum malaria candidate vaccines, ChAd63 ME-TRAP and MVA ME-TRAP, in healthy Gambian and Kenyan adults. PLoS One 8: e57726.
40. Ogwang C, Kimani D, Edwards NJ, Roberts R, Mwacharo J, et al. (2015) Prime-boost vaccination with chimpanzee adenovirus and modified vaccinia Ankara encoding TRAP provides partial protection against Plasmodium falciparum infection in Kenyan adults. Sci Transl Med 7: 286re285.
41. O'Hara GA, Duncan CJ, Ewer KJ, Collins KA, Elias SC, et al. (2012) Clinical assessment of a recombinant simian adenovirus ChAd63: a potent new vaccine vector. J Infect Dis 205: 772-781.
42. Ishizuka AS, Lyke KE, DeZure A, Berry AA, Richie TL, et al. (2016) Protection against malaria at 1 year and immune correlates following PfSPZ vaccination. Nat Med 22: 614-623.
43. Sissoko MS, Healy SA, Katile A, Omaswa F, Zaidi I, et al. (2017) Safety and efficacy of PfSPZ Vaccine against Plasmodium falciparum via direct venous inoculation in healthy malaria-exposed adults in Mali: a randomised, double-blind phase 1 trial. Lancet Infect Dis 17: 498-509.
44. Singh SK, Roeffen W, Andersen G, Bousema T, Christiansen M, et al. (2015) A Plasmodium falciparum 48/45 single epitope R0.6C subunit protein elicits high levels of transmission blocking antibodies. Vaccine 33: 1981-1986.

45. Nikolaeva D, Draper SJ, Biswas S (2015) Toward the development of effective transmission-blocking vaccines for malaria. Expert Rev Vaccines 14: 653-680.
46. Sabchareon A, Burnouf T, Ouattara D, Attanath P, Bouharoun-Tayoun H, et al. (1991) Parasitologic and clinical human response to immunoglobulin administration in falciparum malaria. Am J Trop Med Hyg 45: 297-308.
47. Cohen S, Mc GI, Carrington S (1961) Gamma-globulin and acquired immunity to human malaria. Nature 192: 733-737.
48. Doolan DL, Dobano C, Baird JK (2009) Acquired immunity to malaria. Clin Microbiol Rev 22: 13-36, Table of Contents.
49. Pombo DJ, Lawrence G, Hirunpetcharat C, Rzepczyk C, Bryden M, et al. (2002) Immunity to malaria after administration of ultra-low doses of red cells infected with Plasmodium falciparum. Lancet 360: 610-617.
50. Genton B, Betuela I, Felger I, Al-Yaman F, Anders RF, et al. (2002) A recombinant blood-stage malaria vaccine reduces Plasmodium falciparum density and exerts selective pressure on parasite populations in a phase 1-2b trial in Papua New Guinea. J Infect Dis 185: 820-827.
51. Takala SL, Coulibaly D, Thera MA, Batchelor AH, Cummings MP, et al. (2009) Extreme polymorphism in a vaccine antigen and risk of clinical malaria: implications for vaccine development. Sci Transl Med 1: 2ra5.
52. Saul A (1987) Kinetic constraints on the development of a malaria vaccine. Parasite Immunol 9: 1-9.
53. Mouchet J, Laventure S, Blanchy S, Fioramonti R, Rakotonjanabelo A, et al. (1997) [The reconquest of the Madagascar highlands by malaria]. Bull Soc Pathol Exot 90: 162-168.
54. Drew DR, Beeson JG (2015) PfRH5 as a candidate vaccine for Plasmodium falciparum malaria. Trends Parasitol 31: 87-88.
55. Baum J, Chen L, Healer J, Lopaticki S, Boyle M, et al. (2009) Reticulocyte-binding protein homologue 5 - an essential adhesin involved in invasion of human erythrocytes by Plasmodium falciparum. Int J Parasitol 39: 371-380.
56. Chen L, Lopaticki S, Riglar DT, Dekiwadia C, Uboldi AD, et al. (2011) An EGF-like protein forms a complex with PfRh5 and is required for invasion of human erythrocytes by Plasmodium falciparum. PLoS Pathog 7: e1002199.
57. Reddy KS, Amlabu E, Pandey AK, Mitra P, Chauhan VS, et al. (2015) Multiprotein complex between the GPI-anchored CyRPA with PfRH5 and PfRipr is crucial for Plasmodium falciparum erythrocyte invasion. Proc Natl Acad Sci U S A 112: 1179-1184.
58. Galaway F, Drought LG, Fala M, Cross N, Kemp AC, et al. (2017) P113 is a merozoite surface protein that binds the N terminus of Plasmodium falciparum RH5. Nat Commun 8: 14333.
59. Crosnier C, Bustamante LY, Bartholdson SJ, Bei AK, Theron M, et al. (2011) Basigin is a receptor essential for erythrocyte invasion by Plasmodium falciparum. Nature 480: 534-537.
60. Rodriguez M, Lustigman S, Montero E, Oksov Y, Lobo CA (2008) PfRH5: a novel reticulocyte-binding family homolog of plasmodium falciparum that binds to the erythrocyte, and an investigation of its receptor. PLoS One 3: e3300.
61. Douglas AD, Williams AR, Illingworth JJ, Kamuyu G, Biswas S, et al. (2011) The blood-stage malaria antigen PfRH5 is susceptible to vaccine-inducible cross-strain neutralizing antibody. Nat Commun 2: 601.
62. de Cassan SC, Draper SJ (2013) Recent advances in antibody-inducing poxviral and adenoviral vectored vaccine delivery platforms for difficult disease targets. Expert Rev Vaccines 12: 365-378.
63. Crosnier C, Wanaguru M, McDade B, Osier FH, Marsh K, et al. (2013) A library of functional recombinant cell-surface and secreted P. falciparum merozoite proteins. Mol Cell Proteomics 12: 3976-3986.
64. Reddy KS, Pandey AK, Singh H, Sahar T, Emmanuel A, et al. (2014) Bacterially expressed full-length recombinant Plasmodium falciparum RH5 protein binds erythrocytes and elicits potent strain-transcending parasite-neutralizing antibodies. Infect Immun 82: 152-164.
65. Chiu CY, Healer J, Thompson JK, Chen L, Kaul A, et al. (2014) Association of antibodies to Plasmodium falciparum reticulocyte binding protein homolog 5 with protection from clinical malaria. Front Microbiol 5: 314.

66. Patel SD, Ahouidi AD, Bei AK, Dieye TN, Mboup S, et al. (2013) Plasmodium falciparum merozoite surface antigen, PfRH5, elicits detectable levels of invasion-inhibiting antibodies in humans. J Infect Dis 208: 1679-1687.
67. Chen L, Xu Y, Healer J, Thompson JK, Smith BJ, et al. (2014) Crystal structure of PfRh5, an essential P. falciparum ligand for invasion of human erythrocytes. Elife 3.
68. Ord RL, Rodriguez M, Yamasaki T, Takeo S, Tsuboi T, et al. (2012) Targeting sialic acid dependent and independent pathways of invasion in Plasmodium falciparum. PLoS One 7: e30251.
69. Wright KE, Hjerrild KA, Bartlett J, Douglas AD, Jin J, et al. (2014) Structure of malaria invasion protein RH5 with erythrocyte basigin and blocking antibodies. Nature 515: 427-430.
70. Bustamante LY, Bartholdson SJ, Crosnier C, Campos MG, Wanaguru M, et al. (2013) A full-length recombinant Plasmodium falciparum PfRH5 protein induces inhibitory antibodies that are effective across common PfRH5 genetic variants. Vaccine 31: 373-379.
71. Wanaguru M, Liu W, Hahn BH, Rayner JC, Wright GJ (2013) RH5-Basigin interaction plays a major role in the host tropism of Plasmodium falciparum. Proc Natl Acad Sci U S A 110: 20735-20740.
72. Hayton K, Dumoulin P, Henschen B, Liu A, Papakrivos J, et al. (2013) Various PfRH5 polymorphisms can support Plasmodium falciparum invasion into the erythrocytes of owl monkeys and rats. Mol Biochem Parasitol 187: 103-110.
73. Hayton K, Gaur D, Liu A, Takahashi J, Henschen B, et al. (2008) Erythrocyte binding protein PfRH5 polymorphisms determine species-specific pathways of Plasmodium falciparum invasion. Cell Host Microbe 4: 40-51.
74. Williams AR, Douglas AD, Miura K, Illingworth JJ, Choudhary P, et al. (2012) Enhancing blockade of Plasmodium falciparum erythrocyte invasion: assessing combinations of antibodies against PfRH5 and other merozoite antigens. PLoS Pathog 8: e1002991.
75. Richards JS, Arumugam TU, Reiling L, Healer J, Hodder AN, et al. (2013) Identification and prioritization of merozoite antigens as targets of protective human immunity to Plasmodium falciparum malaria for vaccine and biomarker development. J Immunol 191: 795-809.
76. Tran TM, Li S, Doumbo S, Doumtabe D, Huang CY, et al. (2013) An intensive longitudinal cohort study of Malian children and adults reveals no evidence of acquired immunity to Plasmodium falciparum infection. Clin Infect Dis 57: 40-47.
77. Ewer KJ, Lambe T, Rollier CS, Spencer AJ, Hill AV, et al. (2016) Viral vectors as vaccine platforms: from immunogenicity to impact. Curr Opin Immunol 41: 47-54.
78. Draper SJ, Moore AC, Goodman AL, Long CA, Holder AA, et al. (2008) Effective induction of high-titer antibodies by viral vector vaccines. Nat Med 14: 819-821.
79. Douglas AD, Baldeviano GC, Lucas CM, Lugo-Roman LA, Crosnier C, et al. (2015) A PfRH5-based vaccine is efficacious against heterologous strain blood-stage Plasmodium falciparum infection in aotus monkeys. Cell Host Microbe 17: 130-139.
80. Draper SJ, Biswas S, Spencer AJ, Remarque EJ, Capone S, et al. (2010) Enhancing blood-stage malaria subunit vaccine immunogenicity in rhesus macaques by combining adenovirus, poxvirus, and protein-in-adjuvant vaccines. J Immunol 185: 7583-7595.
81. Payne RO, Silk SE, Elias SC, Milne KH, Rawlinson TA, et al. (2017) Human vaccination against Plasmodium vivax Duffy-binding protein induces strain-transcending antibodies. JCI Insight: In press.
82. Payne RO, Milne KH, Elias SC, Edwards NJ, Douglas AD, et al. (2016) Demonstration of the Blood-Stage Controlled Human Malaria Infection Model to Assess Efficacy of the Plasmodium falciparum AMA1 Vaccine FMP2.1/AS01. J Infect Dis 213: 1743-1751.
83. Duncan CJ, Sheehy SH, Ewer KJ, Douglas AD, Collins KA, et al. (2011) Impact on malaria parasite multiplication rates in infected volunteers of the protein-in-adjuvant vaccine AMA1-C1/Alhydrogel+CPG 7909. PLoS One 6: e22271.
84. Kester KE, Cummings JF, Ofori-Anyinam O, Ockenhouse CF, Krzych U, et al. (2009) Randomized, double-blind, phase 2a trial of falciparum malaria vaccines RTS,S/AS01B and RTS,S/AS02A in malaria-naive adults: safety, efficacy, and immunologic associates of protection. J Infect Dis 200: 337-346.

85. Hjerrild KA, Jin J, Wright KE, Brown RE, Marshall JM, et al. (2016) Production of full-length soluble Plasmodium falciparum RH5 protein vaccine using a Drosophila melanogaster Schneider 2 stable cell line system. Sci Rep 6: 30357.
86. McMillan R, Longmire RL, Yelenosky R, Lang JE, Heath V, et al. (1972) Immunoglobulin synthesis by human lymphoid tissues: normal bone marrow as a major site of IgG production. J Immunol 109: 1386-1394.
87. Amanna IJ, Carlson NE, Slifka MK (2007) Duration of humoral immunity to common viral and vaccine antigens. N Engl J Med 357: 1903-1915.
88. Halliley JL, Tipton CM, Liesveld J, Rosenberg AF, Darce J, et al. (2015) Long-Lived Plasma Cells Are Contained within the CD19(-)CD38(hi)CD138(+) Subset in Human Bone Marrow. Immunity 43: 132-145.
89. Hill DL, Carr EJ, Rutishauser T, Moncunill G, Campo JJ, et al. (2020) Immune system development varies according to age, location, and anemia in African children. Sci Transl Med 12.
90. Tanzania Commission for AIDS (TACAIDS) ZACZ, National Bureau of, Statistics (NBS) OotCGSO, and ICF International, (2013) Tanzania HIV/AIDS and Malaria Indicator Survey 2011-12. Dar es Salaam, Tanzania: TACAIDS, ZAC, NBS, OCGS, and ICF International.
91. Regules JA, Cicatelli SB, Bennett JW, Paolino KM, Twomey PS, et al. (2016) Fractional Third and Fourth Dose of RTS,S/AS01 Malaria Candidate Vaccine: A Phase 2a Controlled Human Malaria Parasite Infection and Immunogenicity Study. J Infect Dis 214: 762-771.
92. Clem AS (2011) Fundamentals of vaccine immunology. J Glob Infect Dis 3: 73-78.
93. Medscape (2017) Yellow fever vaccine.
94. WHO. (2017) International travel and health.
95. `Ministry of Health CD, Gender, Elderly and Children (MoHCDEC) (2020) NATIONAL GUIDELINE OF CLINICAL MANAGEMENT AND INFECTION PREVENTION AND CONTROL OF NOVEL CORONAVIRUS (COVID-19). Tanzania: Government of Tanzania.
96. Gibson BE, Todd A, Roberts I, Pamphilon D, Rodeck C, et al. (2004) Transfusion guidelines for neonates and older children. Br J Haematol 124: 433-453.
97. Howie SR (2011) Blood sample volumes in child health research: review of safe limits. Bull World Health Organ 89: 46-53.
98. Services. DoANIoAaIDNIoHUDoHaH (2017) Division of AIDS (DAIDS) Table for Grading the Severity of Adult and Pediatric Adverse Events. Division of AIDS National Institute of Allergy and Infectious Diseases National Institutes of Health US Department of Health and Human Services.
99. US-FDA. (2014) Guidance for Industry: Toxicity Grading Scale for Healthy Adult and Adolescent Volunteers Enrolled in Preventive Vaccine Clinical Trials.
100. Afolabi MO, Tiono AB, Adetifa UJ, Yaro JB, Drammeh A, et al. (2016) Safety and Immunogenicity of ChAd63 and MVA ME-TRAP in West African children and infants. Mol Ther.
101. WHO. (1998) PROPOSED INTERNATIONAL GUIDELINES ON ETHICAL ISSUES IN MEDICAL GENETICS AND GENETIC SERVICES: Report of a WHO meeting on Ethical Issues in Medical Genetics. 15 p.
102. (2004) Human Tissue Act 2004.
103. Petrini C (2010) "Broad" consent, exceptions to consent and the question of using biological samples for research purposes different from the initial collection purpose. Soc Sci Med 70: 217-220.
104. Ballou WR, Cahill CP (2007) Two decades of commitment to malaria vaccine development: GlaxoSmithKline Biologicals. Am J Trop Med Hyg 77: 289-295.
105. Hill AV (2006) Pre-erythrocytic malaria vaccines: towards greater efficacy. Nat Rev Immunol 6: 21-32.

### 21. APPENDIX A: SCHEDULE OF PROCEDURES

| Group 1a (18 to 45 years) | | Screening | Vaccination and Post Vacciation Follow-up Period | | | | | | | | | | | | | | | | | | | | | | | | | | | Long Term Follow-up | | | | |
|---|---|---|---|---|---|---|---|---|---|---|---|---|---|---|---|---|---|---|---|---|---|---|---|---|---|---|---|---|---|---|---|---|---|---|
| Study Days Relative to First Vaccination | | `-30- to 0 | 0 | 1 | 2 | 3 | 4 | 5 | 6 | 7 | 14 | 28 | 29 | 30 | 31 | 32 | 33 | 34 | 35 | 42 | 56 | 57 | 58 | 59 | 60 | 61 | 62 | 63 | 70 | 84 | 140 | 168 | 365 | 730 |
| Study Visit Code | | SC** | V1* | V1+1 | V1+2 | V1+3 | V1+4 | V1+5 | V1+6 | V1+7 | V1+14 | V2* | V2+1 | V2+2 | V2+3 | V2+4 | V2+5 | V2+6 | V2+7 | V2+14 | V3* | V3+1 | V3+2 | V3+3 | V3+4 | V3+5 | V3+6 | V3+7 | V3+14 | V3+28 | V3+84 | V3+112 | V3+309 | V3+674 |
| Window Relative to each Study Visit | | | 0 | - | ±1 | - | - | - | - | -1/+3 | -1/+3 | -7d/+14d | - | ±1 | - | - | - | - | -1/+3 | -1/+3 | -7d/+14d | - | ±1 | - | - | - | - | -1/+3 | -1/+3 | -7/+2 | ±7 | ±28 | ±28 | ±28 |
| Home Visits | | | | | | X | X | X | X | | | | | | X | X | X | X | | | | | | | | | | | | | | | | |
| Clinic Visits | | X | X | X | X | | | | | X | X | X | X | X | | | | | X | X | X | X | X | | | | | X | X | X | X | X | X | X |
| Vaccination (PfRH5/Matrix M ) | | | X | | | | | | | | | X | | | | | | | | | X | | | | | | | | | | | | | |
| **Clinical Assessment** | | | | | | | | | | | | | | | | | | | | | | | | | | | | | | | | | | |
| Informed Consent and/or Eligibility | | X | X | | | | | | | | | | | | | | | | | | | | | | | | | | | | | | | |
| Medical History with demographic data | | X | | | | | | | | | | | | | | | | | | | | | | | | | | | | | | | | |
| Enrolment | | | X | | | | | | | | | | | | | | | | | | | | | | | | | | | | | | | |
| Photo taken and Temporary ID Card | | X | | | | | | | | | | | | | | | | | | | | | | | | | | | | | | | | |
| Permanent ID Card | | | X | | | | | | | | | | | | | | | | | | | | | | | | | | | | | | | |
| Physical Examination | •Detailed | X | | | | | | | | | | | | | | | | | | | | | | | | | | | | | | | | |
| | •Focused | | X | | X | | | | | X | X | X | | X | | | | | X | X | X | | X | | | | | X | X | X | X | | | X |
| Physical Assessments | •Vital Signs | X | X | | X | | | | | X | X | X | | X | | | | | X | X | X | | X | | | | | X | X | X | X | | | X |
| | •Weight and Height | X | X# | | | | | | | | | X# | | | | | | | | | X# | | | | | | | | | | | | | X# |
| Adverse Events Data Collection | •Solicited | | X | X | X | X | X | X | X | X | | X | X | X | X | X | X | X | X | | X | X | X | X | X | X | X | X | | | | | | |
| | •Unsolicited | | X | X | X | X | X | X | X | X | X | X | X | X | X | X | X | X | X | X | X | X | X | X | X | X | X | X | X | X | | | | |
| | •SAE & New Medical Conditions | X | X | X | X | X | X | X | X | X | X | X | X | X | X | X | X | X | X | X | X | X | X | X | X | X | X | X | X | X | X | X | | X |
| Supply Insecticide Treated Nets (ITN) | | X | | | | | | | | | | | | | | | | | | | | | | | | | | | | | | | | |
| **Laboratory Assessment:** Numbers indicate volume of blood to be collected at each timepoint | | | | | | | | | | | | | | | | | | | | | | | | | | | | | | | | | | |
| Hematology | •RBC, HGB, platelets, WBC with differential (neutrophil, lymphocyte and eosinophil) counts | 0.5 | 0.5 | | | | | | | 0.5 | 0.5 | 0.5 | | | | | | | 0.5 | 0.5 | 0.5 | | | | | | | 0.5 | 0.5 | 0.5 | 0.5 | 0.5 | 0.5 | 0.5 |
| Biochemistry | •ALT & Creatinine, Iron Status& Bleeding indices€ | 1 | 1 | | | | | | | 1 | 1 | 1 | | | | | | | 1 | 1 | 1 | | | | | | | 1 | 1 | 1 | 1 | 1 | | 1 |
| Parasitology | •Thick blood smear | 0.5 | | | | | | | | | | 0.5 | | | | | | | | | 0.5 | | | | | | | | | 0.5 | 0.5 | 0.5 | 0.5 | 0.5 |
| Serology [HIV, HBV and HCV] | | 1 | | | | | | | | | | | | | | | | | | | | | | | | | | | | | | | | |
| Serology (Anti-Schizont) | | 3 | | | | | | | | | | | | | | | | | | | | | | | | | | | | | | | | |
| Immunology | | | 103 | 3 | | | | | | | 70 | 73 | 3 | | | | | | 30 | 70 | 73 | 3 | | | | | | 30 | 100 | 70 | 70 | 70 | 70 | 70 |
| Serum (Pregnancy test)*** | | X | X | | | | | | | | | X | | | | | | | | | X | | | | | | | | | | | | | |
| Bone Marrow Aspiration$ | | | X | | | | | | | | | | | | | | | | | | | | | | | | | | | X | | | | |
| Urine Dipstick [Blood , Glucose, and Protein] | | X | | | | | | | | | | | | | | | | | | | | | | | | | | | | | | | | |
| Total Blood Volume withdrawn/Visit (mL▶ | | 6.0 | 104.5 | 3.0 | 0.0 | 0.0 | 0.0 | 0.0 | 0.0 | 1.5 | 71.5 | 75.0 | 3.0 | 0.0 | 0.0 | 0.0 | 0.0 | 0.0 | 31.5 | 71.5 | 75.0 | 3.0 | 0.0 | 0.0 | 0.0 | 0.0 | 0.0 | 31.5 | 101.5 | 72.0 | 72.0 | 72.0 | 71.0 | 72.0 |
| Cumulative Blood Volume withdrawn (mL▶ | | 6.5 | 111.0 | 114.0 | 114.0 | 114.0 | 114.0 | 114.0 | 114.0 | 115.5 | 187.0 | 262.0 | 265.0 | 265.0 | 265.0 | 265.0 | 265.0 | 265.0 | 296.5 | 368.0 | 443.0 | 446.0 | 446.0 | 446.0 | 446.0 | 446.0 | 446.0 | 477.5 | 579.0 | 651.0 | 723.0 | 795.0 | 866.0 | 938.0 |

* Laboratory tests for this visit must be done within 24 hours before vaccination.

** This includes a screening visit 2 when participant will receive their results

*** Only for women

### Only Weight will be measured

&: Iron status will be conducted retrospectively on screening samples

€: Bleeding indices (Prothrombin Time/INR and platelets tests) will be done once at baseline and 4 weeks post last vaccination before bone marrow aspiration

$: Bone marrow will be collected once either at baseline or 4 weeks post last vaccination for each volunteer

| Group 1b (18-45 years) | | Screening | Vaccination and Post Vacciation Follow-up Period | | | | | | | | | | | | | | | | | | | | | | | | | | | | Long Term Follow-up | | | | |
|---|---|---|---|---|---|---|---|---|---|---|---|---|---|---|---|---|---|---|---|---|---|---|---|---|---|---|---|---|---|---|---|---|---|---|---|
| Study Days Relative to First Vaccination | | `-30- to 0 | 0 | 1 | 2 | 3 | 4 | 5 | 6 | 7 | 14 | 28 | 29 | 30 | 31 | 32 | 33 | 34 | 35 | 42 | 56 | 182 | 183 | 184 | 185 | 186 | 187 | 188 | 189 | 196 | 210 | 266 | 294 | 491 | 856 |
| Study Visit Code | | SC** | V1* | V1+1 | V1+2 | V1+3 | V1+4 | V1+5 | V1+6 | V1+7 | V1+14 | V2* | V2+1 | V2+2 | V2+3 | V2+4 | V2+5 | V2+6 | V2+7 | V2+14 | V2+28 | V3* | V3+1 | V3+2 | V3+3 | V3+4 | V3+5 | V3+6 | V3+7 | V3+14 | V3+28 | V3+84 | V3+112 | V3+309 | V3+674 |
| Window Relative to each Study Visit | | | 0 | - | ±1 | - | - | - | - | -1/+3 | -1/+3 | -7d/+14d | - | ±1 | - | - | - | - | -1/+3 | -1/+3 | ±7 | -30/+14d | - | ±1 | - | - | - | - | -1/+3 | -1/+3 | -7/+2 | ±7 | ±7 | ±7 | ±7 |
| Home Visits | | | | | | X | X | X | X | | | | | | X | X | X | X | | | | | | | X | X | X | X | | | | | | | |
| Clinic Visits | | X | X | X | X | | | | | X | X | X | X | X | | | | | X | X | X | X | X | X | | | | | X | X | X | X | X | X | X |
| Vaccination (PfRH5 Matrix M) | | | X | | | | | | | | | X | | | | | | | | | | X | | | | | | | | | | | | | |
| **Clinical Assessment** | | | | | | | | | | | | | | | | | | | | | | | | | | | | | | | | | | | |
| Informed Consent and/or Eligibility | | X | X | | | | | | | | | | | | | | | | | | | | | | | | | | | | | | | | |
| Medical History with demographic data | | X | | | | | | | | | | | | | | | | | | | | | | | | | | | | | | | | | |
| Enrolment | | | X | | | | | | | | | | | | | | | | | | | | | | | | | | | | | | | | |
| Photo taken and Temporary ID Card | | X | | | | | | | | | | | | | | | | | | | | | | | | | | | | | | | | | |
| Permanent ID Card | | | X | | | | | | | | | | | | | | | | | | | | | | | | | | | | | | | | |
| Physical Examination | •Detailed | X | | | | | | | | | | | | | | | | | | | | | | | | | | | | | | | | | |
| | •Focused | | X | | X | | | | | X | X | X | | X | | | | | X | X | | X | | X | | | | | X | X | X | X | | | X |
| Physical Assessments | •Vital Signs | X | X | | X | | | | | X | X | X | | X | | | | | X | X | | X | | X | | | | | X | X | X | X | | | X |
| | •Weight and Height | X | X[#] | | | | | | | | | X[#] | | | | | | | | | | X[#] | | | | | | | | | | | | | X[#] |
| Adverse Events Data Collection | •Solicited | | X | X | X | X | X | X | X | X | | X | X | X | X | X | X | X | X | | | X | X | X | X | X | X | X | X | | | | | | |
| | •Unsolicited | | X | X | X | X | X | X | X | X | X | X | X | X | X | X | X | X | X | X | | X | X | X | X | X | X | X | X | X | X | | | | |
| | •SAE & New Medical Conditions | | X | X | X | X | X | X | X | X | X | X | X | X | X | X | X | X | X | X | | X | X | X | X | X | X | X | X | X | X | X | X | X | X |
| Supply Insecticide Treated Nets (ITN) | | X | | | | | | | | | | | | | | | | | | | | | | | | | | | | | | | | | |
| **Laboratory Assessment:** Numbers indicate volume of blood to be collected at each timepoint | | | | | | | | | | | | | | | | | | | | | | | | | | | | | | | | | | | |
| Hematology | •RBC, HGB, platelets, WBC with differential (neutrophil, lymphocyte and eosinophil) counts | 0.5 | 0.5 | | | | | | | 0.5 | 0.5 | 0.5 | | | | | | | 0.5 | 0.5 | | 0.5 | | | | | | | 0.5 | 0.5 | 0.5 | 0.5 | 0.5 | 0.5 | 0.5 |
| Biochemistry | •ALT & Creatinine, Iron Status[&], Bleeding indices[€] | 1 | 1 | | | | | | | 1 | 1 | 1 | | | | | | | 1 | 1 | | 1 | | | | | | | 1 | 1 | 1 | 1 | 1 | 1 | 1 |
| Parasitology | •Thick blood smear | 0.5 | | | | | | | | | | 0.5 | | | | | | | | | | 0.5 | | | | | | | | | 0.5 | 0.5 | 0.5 | 0.5 | 0.5 |
| Serology [HIV, HBV and HCV] | | 1 | | | | | | | | | | | | | | | | | | | | | | | | | | | | | | | | | |
| Serology (Anti-Schizont) | | 3 | | | | | | | | | | | | | | | | | | | | | | | | | | | | | | | | | |
| Immunology | | | 103 | 3 | | | | | | | 70 | 73 | 3 | | | | | | 30 | 70 | 70 | 73 | 3 | | | | | | 30 | 100 | 100 | 70 | 70 | 70 | 70 |
| Bone Marrow Aspiration[$] | | | X | | | | | | | | | | | | | | | | | | | | | | | | | | | | X | | | | |
| Urine Dipstick [Blood, Glucose and Protein] | | X | | | | | | | | | | | | | | | | | | | | | | | | | | | | | | | | | |
| Total Blood Volume withdrawn/Visit (mL▶ | | 6.0 | 104.5 | 3.0 | 0.0 | 0.0 | 0.0 | 0.0 | 0.0 | 1.5 | 71.5 | 75.0 | 3.0 | 0.0 | 0.0 | 0.0 | 0.0 | 0.0 | 31.5 | 71.5 | 70.0 | 75.0 | 3.0 | 0.0 | 0.0 | 0.0 | 0.0 | 0.0 | 31.5 | 101.5 | 102.0 | 72.0 | 72.0 | 72.0 | 72.0 |
| Cumulative Blood Volume withdrawn (mL▶ | | 6.5 | 111.0 | 114.0 | 114.0 | 114.0 | 114.0 | 114.0 | 114.0 | 115.5 | 187.0 | 262.0 | 265.0 | 265.0 | 265.0 | 265.0 | 265.0 | 265.0 | 296.5 | 368.0 | 438.0 | 513.0 | 516.0 | 516.0 | 516.0 | 516.0 | 516.0 | 516.0 | 547.5 | 649.0 | 751.0 | 823.0 | 895.0 | 967.0 | 1039.0 |

* Laboratory tests for this visit must be done within 24 hours before vaccination.

** This includes a screening visit 2 when participant will receive their results

### Only Weight will be measured

&: Iron status will be conducted retrospectively on screening samples

€: Bleeding indices (Prothrombin Time/INR and platelets tests) will be done once at baseline and 4 weeks post last vaccination before bone marrow aspiration

$: Bone marrow will be collected once either at baseline or 4 weeks post last vaccination for each volunteer

| Group 2a (5 to 17 months) | | Screening | Vaccination and Post Vacciation Follow-up Period | | | | | | | | | | | | | | | | | | | | | | | | | | Long Term Follow-up | | | | |
|---|---|---|---|---|---|---|---|---|---|---|---|---|---|---|---|---|---|---|---|---|---|---|---|---|---|---|---|---|---|---|---|---|---|
| Study Days Relative to First Vaccination | | `-30- to 0 | 0 | 1 | 2 | 3 | 4 | 5 | 6 | 7 | 14 | 28 | 29 | 30 | 31 | 32 | 33 | 34 | 35 | 42 | 56 | 57 | 58 | 59 | 60 | 61 | 62 | 63 | 70 | 84 | 140 | 168 | 365 | 730 |
| Study Visit Code | | SC** | V1* | V1+1 | V1+2 | V1+3 | V1+4 | V1+5 | V1+6 | V1+7 | V1+14 | V2* | V2+1 | V2+2 | V2+3 | V2+4 | V2+5 | V2+6 | V2+7 | V2+14 | V3* | V3+1 | V3+2 | V3+3 | V3+4 | V3+5 | V3+6 | V3+7 | V3+14 | V3+28 | V3+84 | V3+112 | V3+309 | V3+674 |
| Window Relative to each Study Visit | | | 0 | - | ±1 | - | - | - | - | -1/+3 | -1/+3 | -7d/+14d | - | ±1 | - | - | - | - | -1/+3 | -1/+3 | -7d/+14d | - | ±1 | - | - | - | - | -1/+3 | -1/+3 | -7/+2 | ±7 | ±7 | ±28 | ±28 |
| Home Visits | | | | X | | X | X | X | X | | | | X | | X | X | X | X | | | | | | | | | | | | | | | | |
| Clinic Visits | | X | X | | X | | | | | X | X | X | | X | | | | | X | X | X | | X | | | | | X | X | X | X | X | X | X |
| Vaccination (PfRH5/Matrix M ) | | | X | | | | | | | | | X | | | | | | | | | X | | | | | | | | | | | | | |
| **Clinical Assessment** | | | | | | | | | | | | | | | | | | | | | | | | | | | | | | | | | | |
| Informed Consent and/or Eligibility | | X | X | | | | | | | | | | | | | | | | | | | | | | | | | | | | | | | |
| Medical History with demographic data | | X | | | | | | | | | | | | | | | | | | | | | | | | | | | | | | | | |
| Enrolment | | | X | | | | | | | | | | | | | | | | | | | | | | | | | | | | | | | |
| Photo taken and Temporary ID Card | | X | | | | | | | | | | | | | | | | | | | | | | | | | | | | | | | | |
| Permanent ID Card | | | X | | | | | | | | | | | | | | | | | | | | | | | | | | | | | | | |
| Physical Examination | •Detailed | X | | | | | | | | | | | | | | | | | | | | | | | | | | | | | | | | |
| | •Focused | | X | | X | | | | | X | X | X | | X | | | | | X | X | X | | X | | | | | X | X | X | X | | | X |
| Physical Assessments | •Vital Signs | X | X | | X | | | | | X | X | X | | X | | | | | X | X | X | | X | | | | | X | X | X | X | | | X |
| | •Weight and Height | X | X# | | | | | | | | | X# | | | | | | | | | X# | | | | | | | | | | | | | X# |
| Adverse Events Data Collection | •Solicited | | X | X | X | X | X | X | X | X | | X | X | X | X | X | X | X | X | | X | X | X | X | X | X | X | X | | | | | | |
| | •Unsolicited | | X | X | X | X | X | X | X | X | X | X | X | X | X | X | X | X | X | X | X | X | X | X | X | X | X | X | X | X | | | | |
| | •SAE & New Medical Conditions | X | X | X | X | X | X | X | X | X | X | X | X | X | X | X | X | X | X | X | X | X | X | X | X | X | X | X | X | X | X | X | X | X |
| Supply Insecticide Treated Nets (ITN) | | X | | | | | | | | | | | | | | | | | | | | | | | | | | | | | | | | |
| **Laboratory Assessment:** Numbers indicate volume of blood to be collected at each timepoint | | | | | | | | | | | | | | | | | | | | | | | | | | | | | | | | | | |
| Hematology | •RBC, HGB, platelets, WBC with differential (neutrophil, lymphocyte and eosinophil) counts | 0.5 | 0.5 | | | | | | | 0.5 | 0.5 | 0.5 | | | | | | | 0.5 | 0.5 | 0.5 | | | | | | | 0.5 | 0.5 | 0.5 | 0.5 | 0.5 | 0.5 | 0.5 |
| Biochemistry | •ALT & Creatinine | 1 | 1 | | | | | | | 1 | 1 | 1 | | | | | | | 1 | 1 | 1 | | | | | | | 1 | 1 | 1 | 1 | 1 | 1 | 1 |
| Parasitology | •Thick blood smear | 0.5 | | | | | | | | | | 0.5 | | | | | | | | | 0.5 | | | | | | | | | 0.5 | 0.5 | 0.5 | 0.5 | 0.5 |
| Serology [HIV, HBV and HCV] | | 1 | | | | | | | | | | | | | | | | | | | | | | | | | | | | | | | | |
| Serology (Anti-Schizont) | | 3 | | | | | | | | | | | | | | | | | | | | | | | | | | | | | | | | |
| Immunology | | | 10 | | | | | | | | 10 | 10 | | | | | | | 5 | 10 | 10 | | | | | | | 5 | 10 | 10 | 10 | 10 | 10 | 10 |
| Urine Dipstick [Blood, Glucose and Protein] | | X | | | | | | | | | | | | | | | | | | | | | | | | | | | | | | | | |
| Total Blood Volume withdrawn/Visit (mL▶ | | 6.0 | 11.5 | 0.0 | 0.0 | 0.0 | 0.0 | 0.0 | 0.0 | 1.5 | 11.5 | 12.0 | 0.0 | 0.0 | 0.0 | 0.0 | 0.0 | 0.0 | 6.5 | 11.5 | 12.0 | 0.0 | 0.0 | 0.0 | 0.0 | 0.0 | 0.0 | 6.5 | 11.5 | 12.0 | 12.0 | 12.0 | 12.0 | 12.0 |
| Cumulative Blood Volume withdrawn (mL▶ | | 6.5 | 18.0 | 18.0 | 18.0 | 18.0 | 18.0 | 18.0 | 18.0 | 19.5 | 31.0 | 43.0 | 43.0 | 43.0 | 43.0 | 43.0 | 43.0 | 43.0 | 49.5 | 61.0 | 73.0 | 73.0 | 73.0 | 73.0 | 73.0 | 73.0 | 73.0 | 79.5 | 91.0 | 103.0 | 115.0 | 127.0 | 139.0 | 151.0 |

* Laboratory tests for this visit must be done within 24 hours before vaccination.

** This includes a screening visit 2 when participant will receive their results

### Only Weight will be measured

| Group 2b, 2c, 2d (5 to 17 months) | | Screening | Vaccination and Post Vacciation Follow-up Period | | | | | | | | | | | | | | | | | | | | | | | | | | | | Long Term Follow-up | | | | |
|---|---|---|---|---|---|---|---|---|---|---|---|---|---|---|---|---|---|---|---|---|---|---|---|---|---|---|---|---|---|---|---|---|---|---|---|
| Study Days Relative to First Vaccination | | `-30- to 0 | 0 | 1 | 2 | 3 | 4 | 5 | 6 | 7 | 14 | 28 | 29 | 30 | 31 | 32 | 33 | 34 | 35 | 42 | 56 | 182 | 183 | 184 | 185 | 186 | 187 | 188 | 189 | 196 | 210 | 266 | 294 | 491 | 856 |
| Study Visit Code | | SC** | V1* | V1+1 | V1+2 | V1+3 | V1+4 | V1+5 | V1+6 | V1+7 | V1+14 | V2* | V2+1 | V2+2 | V2+3 | V2+4 | V2+5 | V2+6 | V2+7 | V2+14 | V2+28 | V3* | V3+1 | V3+2 | V3+3 | V3+4 | V3+5 | V3+6 | V3+7 | V3+14 | V3+28 | V3+84 | V3+112 | V3+309 | V3+674 |
| Window Relative to each Study Visit | | | 0 | - | ±1 | - | - | - | - | -1/+3 | -1/+3 | -7d/+14d | - | ±1 | - | - | - | - | -1/+3 | -1/+3 | ±7 | -30d/+14d | - | ±1 | - | - | - | - | -1/+3 | -1/+3 | -7/+2 | ±7 | ±7 | ±7 | ±7 |
| Home Visits | | | | X | | X | X | X | X | | | | X | | X | X | X | X | | | | | X | | X | X | X | X | | | | | | | |
| Clinic Visits | | X | X | | X | | | | | X | X | X | | X | | | | | X | X | X | X | | X | | | | | X | X | X | X | X | X | X |
| Vaccination (PfRH5 Matrix M) | | | X | | | | | | | | | X | | | | | | | | | | X | | | | | | | | | | | | | |
| **Clinical Assessment** | | | | | | | | | | | | | | | | | | | | | | | | | | | | | | | | | | | |
| Informed Consent and/or Eligibility | | X | X | | | | | | | | | | | | | | | | | | | | | | | | | | | | | | | | |
| Medical History with demographic data | | X | | | | | | | | | | | | | | | | | | | | | | | | | | | | | | | | | |
| Enrolment | | | X | | | | | | | | | | | | | | | | | | | | | | | | | | | | | | | | |
| Photo taken and Temporary ID Card | | X | | | | | | | | | | | | | | | | | | | | | | | | | | | | | | | | | |
| Permanent ID Card | | | X | | | | | | | | | | | | | | | | | | | | | | | | | | | | | | | | |
| Physical Examination | •Detailed | X | | | | | | | | | | | | | | | | | | | | | | | | | | | | | | | | | |
| | •Focused | | X | | X | | | | | X | X | X | | X | | | | | X | X | | X | | X | | | | | X | X | X | X | | | X |
| Physical Assessments | •Vital Signs | X | X | | X | | | | | X | X | X | | X | | | | | X | X | | X | | X | | | | | X | X | X | X | | | X |
| | •Weight and Height | X | X# | | | | | | | | | X# | | | | | | | | | | X# | | | | | | | | | | | | | X# |
| Adverse Events Data Collection | •Solicited | | X | X | X | X | X | X | X | X | | X | X | X | X | X | X | X | X | | | X | X | X | X | X | X | X | X | | | | | | |
| | •Unsolicited | | X | X | X | X | X | X | X | X | X | X | X | X | X | X | X | X | X | X | | X | X | X | X | X | X | X | X | X | X | | | | |
| | •SAE & New Medical Conditions | X | X | X | X | X | X | X | X | X | X | X | X | X | X | X | X | X | X | X | | X | X | X | X | X | X | X | X | X | X | X | X | X | X |
| Supply Insecticide Treated Nets (ITN) | | X | | | | | | | | | | | | | | | | | | | | | | | | | | | | | | | | | |
| **Laboratory Assessment:** Numbers indicate volume of blood to be collected at each timepoint | | | | | | | | | | | | | | | | | | | | | | | | | | | | | | | | | | | |
| Hematology | •RBC, HGB, platelets, WBC with differential (neutrophil, lymphocyte and eosinophil) counts | 0.5 | 0.5 | | | | | | | 0.5 | 0.5 | 0.5 | | | | | | | 0.5 | 0.5 | | 0.5 | | | | | | | 0.5 | 0.5 | 0.5 | 0.5 | 0.5 | 0.5 | 0.5 |
| Biochemistry | •ALT & Creatinine | 1 | 1 | | | | | | | 1 | 1 | 1 | | | | | | | 1 | 1 | | 1 | | | | | | | 1 | 1 | 1 | 1 | 1 | 1 | 1 |
| Parasitology | •Thick blood smear | 0.5 | | | | | | | | | | 0.5 | | | | | | | | | | 0.5 | | | | | | | | | 0.5 | 0.5 | 0.5 | 0.5 | 0.5 |
| Serology [HIV, HBV and HCV] | | 1 | | | | | | | | | | | | | | | | | | | | | | | | | | | | | | | | | |
| Serology (Anti-Schizont) | | 3 | | | | | | | | | | | | | | | | | | | | | | | | | | | | | | | | | |
| Immunology | | | 10 | | | | | | | | 10 | 10 | | | | | | | 5 | 10 | 10 | 10 | | | | | | | 5 | 10 | 10 | 10 | 10 | 10 | 10 |
| Urine Dipstick [Blood, Gllucose and Protein] | | X | | | | | | | | | | | | | | | | | | | | | | | | | | | | | | | | | |
| Total Blood Volume withdrawn/Visit (mL▶ | | 6.0 | 11.5 | 0.0 | 0.0 | 0.0 | 0.0 | 0.0 | 0.0 | 1.5 | 11.5 | 12.0 | 0.0 | 0.0 | 0.0 | 0.0 | 0.0 | 0.0 | 6.5 | 11.5 | 10.0 | 12.0 | 0.0 | 0.0 | 0.0 | 0.0 | 0.0 | 0.0 | 6.5 | 11.5 | 12.0 | 12.0 | 0.0 | 12.0 | 12.0 |
| Cumulative Blood Volume withdrawn (mL▶ | | 6.5 | 18.0 | 18.0 | 18.0 | 18.0 | 18.0 | 18.0 | 18.0 | 19.5 | 31.0 | 43.0 | 43.0 | 43.0 | 43.0 | 43.0 | 43.0 | 43.0 | 49.5 | 61.0 | 71.0 | 83.0 | 83.0 | 83.0 | 83.0 | 83.0 | 83.0 | 83.0 | 89.5 | 101.0 | 113.0 | 125.0 | 125.0 | 137.0 | 149.0 |

* Laboratory tests for this visit must be done within 24 hours before vaccination.

** This includes a screening visit 2 when participant will receive their results

### Only Weight will be measured

### 22. APPENDIX B: SAE REPORTING FLOW CHART

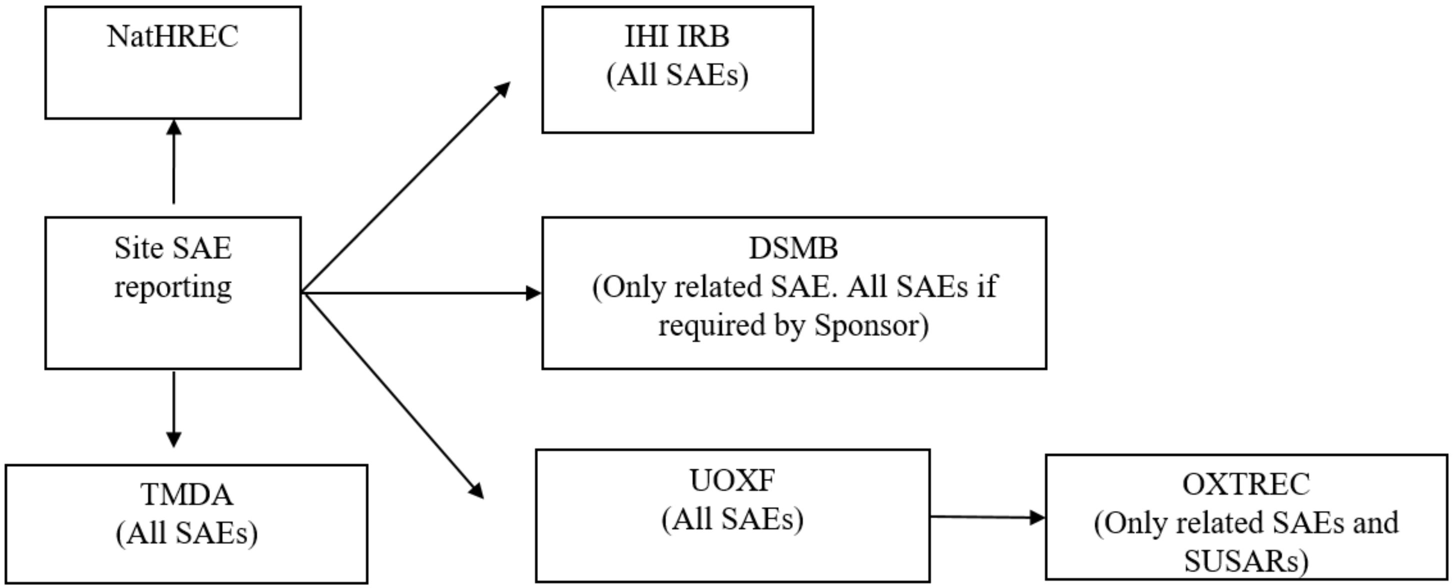

### 23. APPENDIX C: AMENDMENT HISTORY

| Amendment No. | Protocol Version No. | Date issued | Author(s) of changes | Details of Changes made |
|---|---|---|---|---|
| 1.0 | 2.0 | 9th September 2020 | Ally Olotu | • Typos in several sections has been corrected.<br>• Management of potential COVID-19 cases during the study added.<br>• Time window before first vaccination and before each dose has been updated.<br>• Clinical Safety preliminary results of Matrix M adjuvant in infants from Kenya and Burkina Faso added.<br>• Sample storage and future use clarified further.<br>• Sample handling has been clarified further.<br>• The dose of Matrix-M has been changed from 25µg to 50µg.<br>• Age of the infants changed from 6-11 months to 5-17 months.<br>• The title “Publication policy” changed to “Dissemination and Publication Policy”<br>• Clinical trial monitoring has been further clarified in the protocol.<br>• Constitution of the DSMB has been updated to include a Tanzanian paediatrician. |
| 2.0 | 3.0 | 14th April 2021 | Ally Olotu | • The allowed window for the third dose in the delayed vaccination schedule has been updated to -30d/+14d.<br>• For consistency we have replaced the use of SMC with DSMB.<br>• We have combined DSMB meetings 3 and 4 because of their close proximity.<br>• We have corrected table of outlines of study procedures to be consistent with the detail of scheduled visits in the protocol text and remove inconsistencies. |
